# Mutations in the *GBA1*, *LRRK2*, *TMEM175*, *PARK2*, *PINK1*, and *PARK7* genes lead to sex-specific serum metabolic changes in patients with Parkinson’s disease

**DOI:** 10.64898/2026.01.13.26344011

**Authors:** Carmen Marino, Federica Carrillo, Marcello Serra, Tommaso Nuzzo, Matteo Vidali, Sara Pietracupa, Nicola Modugno, Manuela Grimaldi, Francesco Errico, Anna Maria D’Ursi, Teresa Esposito, Alessandro Usiello

**Affiliations:** Department of Pharmacy, University of Salerno, 84084, Fisciano, Salerno, Italy; Institute of Genetics and Biophysics, Italian National Research Council CNR, 80131, Naples, Italy; Department of Biomedical Sciences, University of Cagliari, Monserrato, Italy; Department of Environmental, Biological and Pharmaceutical Sciences and Technologies, Università degli Studi della Campania “Luigi Vanvitelli”, 81100, Caserta, Italy; Laboratory of Translational Neuroscience, CEINGE Biotecnologie Avanzate Franco Salvatore, Naples, Italy; Clinical Pathology Unit, Fondazione IRCCS Ca’ Granda Ospedale Maggiore Policlinico, Milano, Italy; IRCCS INM Neuromed, 86077, Pozzilli, Italy; Department of Human Neuroscience, Sapienza University of Rome, Italy; Department of Agricultural Sciences, University of Naples “Federico II”, 80055, Portici, Italy

**Keywords:** Parkinson’s disease, genetic, sex, NMR spectroscopy, metabolism

## Abstract

Sex differences significantly influence the pathophysiology of Parkinson’s disease (PD). However, it remains unclear whether sex differences affect blood metabolome profiles in patients with PD-related gene mutations compared to healthy controls (HCs).

Here, we conducted a ^1^H-Nuclear Magnetic Resonance (NMR) metabolomic analysis of serum samples from a defined cohort of patients carrying variants in *LRRK2* (n=17), *GBA1* (n=30), *PARK2/PINK1/PARK7* (n=40), and *TMEM175* (n=32) genes. Their serum metabolomic profiles were compared with those of 48 age- and sex-matched HCs. Additionally, 48 patients with idiopathic PD were included to assess the extent to which metabolomic alterations overlapped between genetically defined and idiopathic PD.

Notably, Partial Least Squares Discriminant Analysis indicated significant sexual dimorphism in serum metabolomic profiles between cases and controls across all PD-linked genetic groups. Conversely, multivariate analysis of NMR data did not show clear separation of metabolomic features among patients carrying different pathogenic mutations, nor between genetically defined and idiopathic PD patients.

These findings suggest that, despite genetic heterogeneity, PD patients share a common pattern of systemic metabolic alterations. Furthermore, our findings indicate that sex differences significantly influence the blood metabolic profiles of distinct PD-related gene mutations, resulting in unique serum metabolic profiles in mutation carriers compared with matched controls.

## Introduction

Parkinson’s disease (PD) is a progressive neurodegenerative disorder characterised by the gradual loss of dopaminergic neurons in the substantia nigra pars compacta^1^. PD incidence is higher in males than in females ^2^. However, the molecular causes for this sex bias remain poorly understood. Proposed mechanisms include differential environmental exposure, hormonal influence, and sex-dependent genetic susceptibility ^3–5^.

Additional factors contributing to PD onset include ageing and genetic background. Age represents the most significant risk factor for PD ^6,7^, whereas the influence of genetic status remains an area of ongoing research. The genetics of PD is complex and heterogeneous. Approximately 10-15% of patients have highly penetrant mutations causing familial PD, while many additional mutations contribute to sporadic forms. To date, approximately twenty causative genes have been identified, including *PARK2, PINK1, LRRK2, SNCA, PARK7, VPS35, VPS13C, CHCHD2, TMEM230, TMEM175, ATP13A2, GIGYF2, HTRA2, PLA2G6, FBXO7, EIF4G1, DNAJC6, SYNJ1, UCHL1* and *DNAJC13* ^8–11^. These genes participate in cellular pathways implicated in PD pathogenesis, including intracellular trafficking, oxidative stress responses, mitochondrial metabolism, phospholipid turnover, and the ubiquitin–proteasome system ^12–20^.

In this context, *LRRK2, TMEM175, PARK2, PARK7* and *PINK1* mutations are among the most recurrent monogenic forms of PD. Pathogenic LRRK2 variants, particularly the common p.G2019S substitution, enhance kinase activity and represent the most frequent cause of autosomal dominant PD ^21^.

More recently, the *TMEM175* gene encoding a lysosomal potassium channel has emerged as a relevant PD gene in both familial and sporadic forms ^9,10,22–24^. Our recent work showed that rare *TMEM175* mutations with autosomal dominant transmission, as well as the p.M393T risk variant, are associated with an earlier age at onset ^9–11,24^.

PTEN-induced kinase 1 (*PINK1*) and Parkin RBR E3 ubiquitin-protein ligase (*PARK2*) genes encode proteins essential for mitophagy, mitochondrial dynamics, and are the primary causes of autosomal recessive PD ^25^. *PARK7* (also known as DJ1), encoding a redox-sensitive chaperone that protects neurons from oxidative stress ^26,27^, is also implicated in autosomal recessive PD.

In addition to these causative genes, variants in Glucosylceramidase beta 1 (*GBA1)* represent the most frequent genetic risk factor for PD across populations ^28^. Susceptibility variants such as p.E326K, p.T369M, p.N370S and p.L444P reduce glucocerebrosidase activity, impair lysosomal function and promote α-synuclein accumulation ^29^.

Over the past decade, metabolomic and HPLC studies of serum, plasma, and cerebrospinal fluid have revealed multiple metabolic alterations in PD patients, including disruptions in the metabolism of sugars, amino acids, lipids, and purine metabolism, as well as mitochondrial-related energy homeostasis and antioxidant defence, ultimately reflecting the complex nature of the disorder ^30–41^. However, despite the recognized roles of sex and genetic factors in influencing PD pathophysiology and clinical presentation, only very few metabolomic studies have stratified patients by specific pathogenic mutations across both sexes. This limitation hinders the understanding of how sex and genetic background interact to shape systemic metabolic alterations in PD, particularly in genetic forms, where limited sample sizes often necessitate exploratory analyses.

To address this gap, we conducted an ¹H-Nuclear Magnetic Resonance (NMR)-based serum metabolomics analysis on a genetically and clinically defined cohort of PD patients, carrying at least a pathogenic mutation in the *LRRK2*, *TMEM175*, *PARK2*, *PINK1*, *PARK7*, or *GBA1* genes (n = 119) compared to a sex- and age-matched healthy controls (HCs) (n = 48). Furthermore, to provide a broader clinical reference, we included 48 patients with idiopathic PD in the study.

## Results

### Demographic and clinical characteristics of the study cohort

The study included 167 participants: 119 patients with genetic Parkinson’s disease (gPD), 48 patients with idiopathic Parkinson’s disease (iPD), and 48 healthy controls (HCs) matched for age and sex. The demographic and clinical characteristics, stratified by sex, are summarized **in Table 1 and Table S1.**

**Table 1.** Demographic and clinical characteristics of patients with Parkinson’s disease stratified by specific pathogenic mutations, compared with idiopathic Parkinson’s disease and healthy control subjects.

|  |  | Healthy controls |  |  |  | iPD |  |  |  | gPD |  |  |  |  |  |  |  |  |  |  |  |  |  |  |  | p value |
| --- | --- | --- | --- | --- | --- | --- | --- | --- | --- | --- | --- | --- | --- | --- | --- | --- | --- | --- | --- | --- | --- | --- | --- | --- | --- | --- |
|  |  |  |  |  |  |  |  |  |  | GBA1 |  |  |  | LRRK2 |  |  |  | PARK2/PINK1/PARK7 |  |  |  | TMEM175 |  |  |  |  |
|  |  | N | Median | IQR |  | N | Median | IQR |  | N | Median | IQR |  | N | Median | IQR |  | N | Median | IQR |  | N | Median | IQR |  |  |
| Male | Age (years) | 23 | 64.0 | 58.0 | 77.0 | 23 | 68.0 | 62.0 | 73.0 | 13 | 66.0 | 63.0 | 69.0 | 6 | 60.5 | 55.0 | 68.0 | 22 | 69.0 | 61.0 | 75.0 | 20 | 69.5 | 64.0 | 71.5 | 0.270 (HC vs iPD)<br>>0.999 (HC vs GBA1)<br>0.515 (HC vs LRRK2)<br>0.345 (HC vs PARK2/PINK1/PARK7)<br>0.316 (HC vs TMEM175) |
|  | Age at disease onset (years) |  |  |  |  | 23 | 64.0 | 59.0 | 67.0 | 13 | 61.0 | 57.0 | 66.0 | 6 | 55.5 | 48.0 | 62.0 | 22 | 59.5 | 55.0 | 68.0 | 20 | 61.0 | 57.0 | 65.0 | 0.473 |
|  | Duration of disease (years) |  |  |  |  | 23 | 4.0 | 2.0 | 8.0 | 13 | 5.0 | 2.0 | 8.0 | 6 | 6.0 | 4.0 | 8.0 | 22 | 8.0 | 3.0 | 14.0 | 20 | 6.0 | 2.5 | 9.5 | 0.929 |
|  | LEDD (mg/die) |  |  |  |  | 23 | 400.0 | 205.0 | 705.0 | 13 | 500.0 | 320.0 | 600.0 | 6 | 525.0 | 286.0 | 700.0 | 22 | 402.5 | 360.0 | 779.0 | 20 | 462.5 | 305.0 | 760.0 | 0.292 |
|  | MDS-UPDRS III |  |  |  |  | 23 | 20.0 | 13.0 | 33.0 | 13 | 24.0 | 16.0 | 34.0 | 6 | 14.5 | 10.0 | 17.0 | 22 | 24.5 | 14.0 | 29.0 | 20 | 20.5 | 15.0 | 37.5 | 0.585 |
| Female | Age (years) | 25 | 69.0 | 51.0 | 71.0 | 25 | 68.0 | 63.0 | 71.0 | 17 | 69.0 | 65.0 | 74.0 | 11 | 68.0 | 59.0 | 78.0 | 18 | 67.5 | 59.0 | 71.0 | 12 | 67.5 | 66.0 | 74.5 | 0.861 (HC vs iPD)<br>0.572 (HC vs GBA1)<br>0.705 (HC vs LRRK2)<br>0.512 (HC vs PARK2/PINK1/PARK7)<br>0.757 (HC vs TMEM175) |
|  | Age at disease onset (years) |  |  |  |  | 25 | 63.0 | 60.0 | 66.0 | 17 | 62.0 | 57.0 | 67.0 | 11 | 58.0 | 50.0 | 67.0 | 18 | 56.5 | 47.0 | 64.0 | 12 | 64.0 | 59.0 | 65.0 | 0.242 |
|  | Duration of disease (years) |  |  |  |  | 25 | 5.0 | 4.0 | 7.0 | 17 | 6.0 | 4.0 | 8.0 | 11 | 11.0 | 8.0 | 17.0 | 18 | 7.5 | 3.0 | 15.0 | 12 | 6.5 | 3.0 | 12.0 | 0.191 |
|  | LEDD (mg/die) |  |  |  |  | 25 | 500.0 | 400.0 | 600.0 | 17 | 460.0 | 200.0 | 518.0 | 11 | 780.0 | 352.0 | 850.0 | 18 | 439.5 | 336.0 | 620.0 | 12 | 510.0 | 260.0 | 625.0 | 0.107 |
|  | MDS-UPDRS III |  |  |  |  | 25 | 21.0 | 14.0 | 28.0 | 17 | 16.0 | 10.0 | 23.0 | 11 | 14.0 | 12.0 | 31.0 | 18 | 17.5 | 13.0 | 38.0 | 12 | 22.0 | 16.0 | 34.5 | 0.582 |

Abbreviations: N, number of subjects; IQR, interquartile range; iPD, idiopathic Parkinson's disease; gPD, genetic Parkinson's disease; LEDD, Levodopa equivalent daily dose; MDS-UPDRS III, Movement Disorders Society Unified Parkinson’s Disease Rating Scale part III.
Age comparisons between each PD genetic group and HCs were performed using the Mann–Whitney U test, while clinical parameters across all PD genetic groups were compared using the Kruskal–Wallis test.

Among male gPD patients, 13 had a *GBA1* mutation, 22 carried at least one pathogenic variant in the recessive PD genes (*PARK2*, *PINK1*, or *PARK7*), 20 had a *TMEM175* mutation, and 6 had a *LRRK2* mutation. Among female gPD patients, 17 had a *GBA1* mutation, 18 carried a heterozygous mutation in *PARK2*, *PINK1*, or *PARK7*, 12 had a *TMEM175* mutation, and 11 had a *LRRK2* mutation (**Table 1**). A complete list of mutations is provided in **Table S2**.

Heterozygous carriers of variants in recessive PD-associated genes (*PARK2*, *PINK1*, or *PARK7*) were analyzed as a single subgroup to avoid excessive fragmentation of the cohort, which could compromise statistical power. Similarly, *GBA1* variant carriers were grouped together, despite the known functional heterogeneity of GBA1 variants, which includes their differing effects on glucocerebrosidase activity, lysosomal function, and disease severity ^42^

The iPD group included 23 males and 25 females, and the HC group included 23 males and 25 females (**Table 1**). Both iPD patients and HCs underwent Whole-Exome Sequencing (WES) and were negative for pathogenic mutations or variants in PD-associated genes ^9,11^. Age was comparable across all PD subgroups and HCs in both sexes (**Table 1**). Sex-stratified analyses showed no significant differences in age, age at onset, disease duration, Levodopa equivalent daily dose (LEDD), or Movement Disorder Society revised version of the Unified Parkinson’s Disease Rating Scale Part III (MDS-UPDRS III) scores among the different genetic PD subgroups and the iPD group in either males or females (**Table 1**).

### GBA1 mutations are associated with sex-specific serum metabolomic profiles in Parkinson’s disease patients compared to healthy controls

GBA1 mutations reduce lysosomal β-glucocerebrosidase activity, disrupting sphingolipid metabolism, protein quality control, ER–Golgi trafficking, and α-synuclein clearance ^29,43^. GBA1 variants exhibit biological and clinical heterogeneity, with variations in glucocerebrosidase activity and disease severity spanning from risk variants to those associated with mild, moderate, or severe phenotypes. This variability may underpin diverse lysosomal and metabolic effects. Following previously established classification criteria ^42^, we stratified *GBA1* carriers according to variant severity, distinguishing four groups: mild (N = 8), risk factor (RF; N = 11), likely pathogenic/variants of uncertain significance (LP/VUS; N = 8), and severe (N = 3). The severe subgroup was excluded from the initial comparative analyses due to its limited sample size.

Serum metabolomic profiles were acquired using ¹H-NMR spectroscopy, with one-dimensional CPMG sequences to enhance the detection of low-molecular-weight metabolites (**Figure S1**). PLS-DA of the stratified *GBA1* cohort did not reveal any meaningful separation among the severity-based subgroups (PC1: Q² = –0.65; R² = 0.64), indicating the absence of distinct metabolic fingerprints associated with disease severity (**Figure S2a,b**). Given the absence of detectable differences among distinct *GBA1* carriers and the limited sample size within each subgroup, we pooled all *GBA1* carriers into a single group for metabolomic comparison with HCs. Accordingly, we analyzed serum metabolomic profiles from male (N=13) and female (N=17) PD patients with *GBA1* mutations, compared with age- and sex-matched HCs (male, N=23; female, N=25).

Notably, unsupervised principal component analysis (PCA) (**Figure S3a,b**) and supervised PLS-DA revealed a clear metabolic distinction between PD-*GBA1* patients and HCs in both sexes (**Figure 1a, e**). For the first principal component, Q² and R² values were 0.58 and 0.76 in males, and 0.49 and 0.63 in females, respectively, supporting the robustness of the observed separation. To further validate these findings and mitigate the risk of overfitting, we implemented a Random Forest classification framework, which confirmed the models’ discriminative performance (**Table S3**).

**Figure 1.**
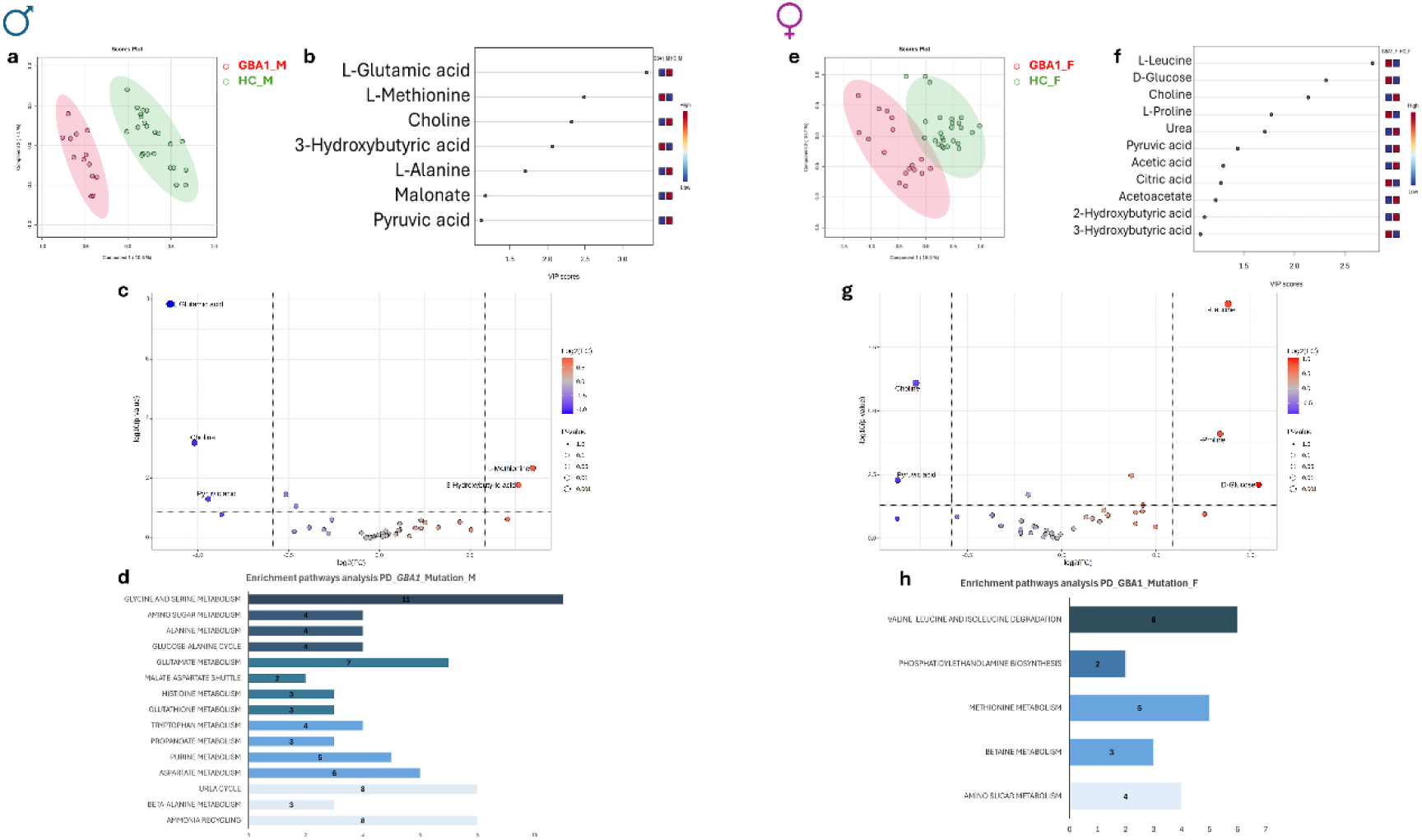
GBA1 mutations in PD patients impact serum metabolome profile in a sex-dependent manner. **a, e.** PLS-DA score plots of serum metabolite levels from genetic Parkinson’s disease (gPD) patients carrying GBA1 mutations (males: 13 patients, red; 23 healthy controls, green; **panel a**; females: 17 patients, red; 25 healthy controls, green; **panel e**). Variance explained: PC1 = 10.6%, PC2 = 7.1% (males); PC1 = 16.3%, PC2 = 10.7% (females). Models validated by 10-fold CV (males: accuracy = 0.86, 0.94; Q² = 0.58, 0.65; R² = 0.76, 0.60; females: accuracy = 0.88, 0.85; Q² = 0.49, 0.61; R² = 0.63, 0.51). **b,f.** Variable Importance in Projection (VIP) plots of metabolites driving group separation in males (**b**) and females (**f**). Only metabolites with VIP > 1 were considered significant. **c,g.** Robust volcano plot displaying the upregulated (red) and downregulated (blue) metabolites in the serum of male **(c)** and female **(g)** PD patients with GBA1 mutation. The absolute fold change value was set at 1.5, while the p-value threshold was defined as <0.05. **d,h**. Pathway enrichment analysis based on ^1^H-NMR serum metabolomics from male **(d)** and female **(h)** gPD patients with GBA1 mutations compared to sex-matched controls. Bars indicate the number of detected metabolites (hits) per pathway. Pathways were considered significant with Hits > 1, p < 0.05, Holm-adjusted p < 0.05, and FDR < 1. Darker colours denote lower p-values. Pathway analysis was conducted using SMPDB (organism: *Homo sapiens*).

In males, the Variable Importance Projection (VIP) score analysis identified L-glutamic acid as the most discriminative metabolite between PD-*GBA1* patients and HCs (**Figure 1b**). This was followed by L-alanine, L-methionine along with energy-related metabolites such as 3-hydroxybutyric acid and pyruvic acid (**Figure 1b**). Choline, which serves as a precursor for membrane phospholipids and acetylcholine biogenesis, also ranked among the top discriminative metabolites between the cases and controls (**Figure 1b**). Robust volcano plot analysis confirmed significantly lower concentrations of L-glutamic acid, choline and pyruvic acid in male PD-*GBA1* patients, whereas L-methionine and 3-hydroxybutyric acid were increased compared with HCs (**Figure 1c, Table S4**).

In female subjects, VIP score analysis identified L-leucine, L-proline, D-glucose, choline, urea, pyruvic acid, and citric acid as discriminative metabolites between cases and controls. Furthermore, ketone bodies further contributed to the blood metabolomic discrimination between female PD-*GBA1* patients and control subjects (**Figure 1f**).

Robust volcano plot analysis confirmed downregulation of choline and pyruvic acid, as well as upregulation of L-leucine, L-proline, and D-glucose compared to healthy controls (**Figure 1g, Table S4**). Heatmap analysis further emphasized sex-based differences in metabolite alterations (**Figure S4**).

Consistent with the NMR data, pathway enrichment analysis revealed dysregulation of metabolism related to glycine-serine, alanine, glutamic acid, tryptophan, and aspartate in male patients when compared to sex-matched HCs (**Figure 1d and Table S5**). Furthermore, alterations in the glucose-alanine cycle, urea cycle, ammonia recycling, and glutathione biosynthesis were also reported in male patients with PD-*GBA1* compared to the control group (**Figure 1d and Table S5**).

In female PD-*GBA1* patients, pathway enrichment analysis showed dysregulated branched-chain amino acid (BCAA) degradation, alterations in methionine and betaine metabolism, and changes in phosphatidylethanolamine biosynthesis compared to sex-matched controls **(Figure 1h; Table S6).**

Overall, our findings demonstrate distinct serum metabolomic profiles when male and female PD-*GBA1* patients were compared to sex-matched HCs. This sex-dependent patterns are consistent with a previously described role of β-glucocerebrosidase in modulating several biochemical pathways ^44^ and with prior metabolomic studies reporting perturbations in amino-acid pathways and the urea cycle in the context of GBA1 dysfunction ^45,46^.

### LRRK2 mutation is associated with sex-specific serum metabolomic profiles in PD patients compared to healthy controls

*LRRK2* encodes a multidomain kinase/GTPase involved in vesicular, lysosomal, autophagic, mitochondrial, and immune regulation, and pathogenic variants that increase its kinase activity are the leading cause of autosomal dominant PD ^21,47^.

To explore how these mutations impact systemic metabolism, we investigated serum metabolomic profiles of male (N=6) and female (N=11) PD patients carrying *LRRK2* mutations, compared with age- and sex-matched HCs (male, N=23; female, N=25).

Unsupervised PCA (**Figure S3c,d**) and PLS-DA revealed a clear serum metabolome profiles separation between PD-*LRRK2* patients and HCs in both sexes, with Q^2^ values for PC1 of 0.58 (males) and 0.59 (females) and with R^2^ values for PC1 of 0.76 for both males and females (**Figure 2a,e**).

**Figure 2.**
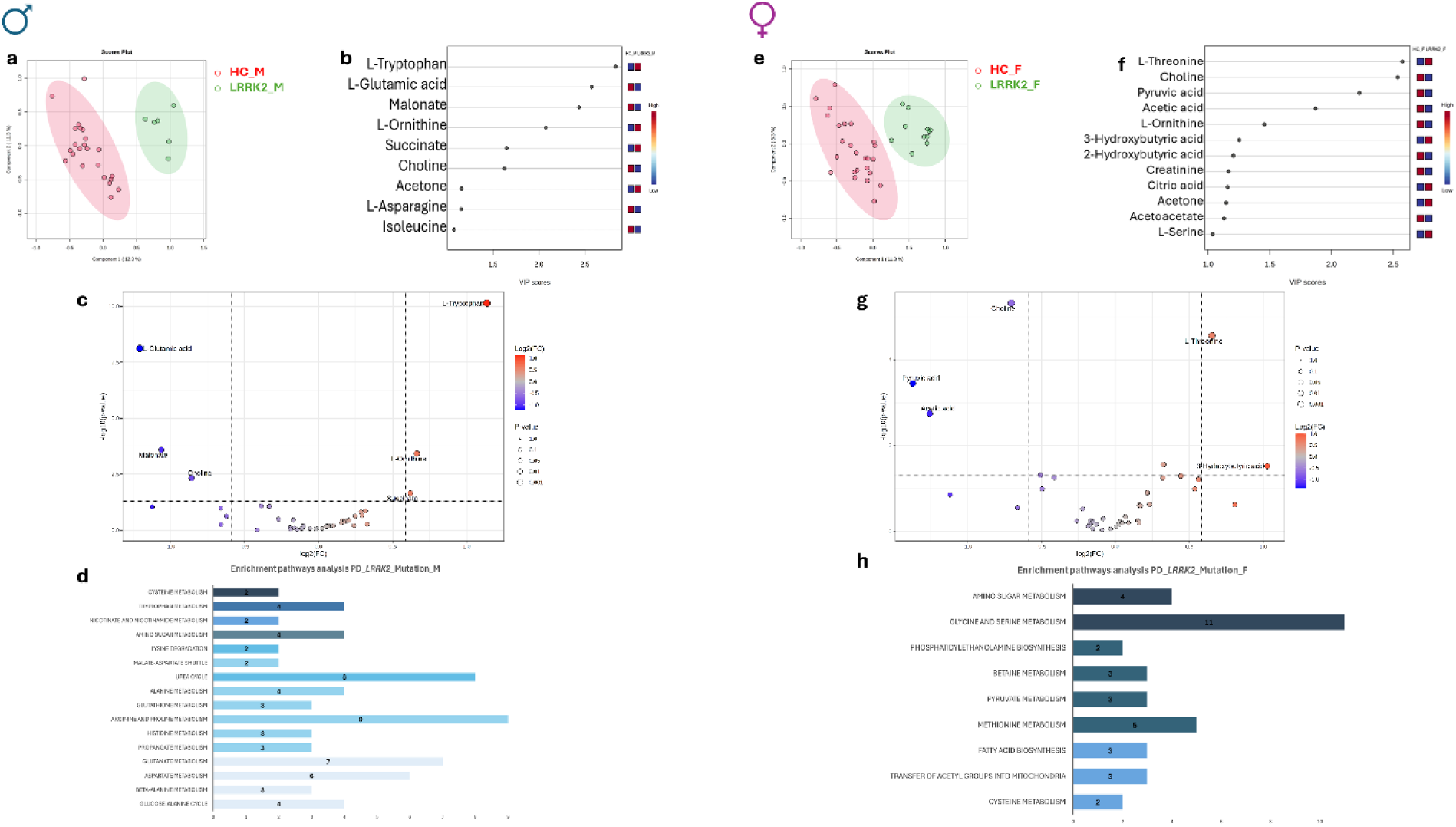
LRRK2 mutations in PD patients impacts serum metabolome profile in a sex-dependent manner. **a, e.** PLS-DA score plots of serum metabolite levels from genetic Parkinson’s disease (gPD) patients carrying LRRK2 mutations (males: 6 patients, red; 23 healthy controls, green; **panel a**; females: 11 patients, red; 25 healthy controls, green; **panel e**). Variance explained: PC1 = 12.3%, PC2 = 11.3% (males); PC1 = 11.3%, PC2 = 8.3% (females). Models validated by 10-fold CV (males: accuracy = 0.93 for both component; Q² = 0.58, 0.62; R² = 0.76, 0.42; females: accuracy = 0.91, 0.88; Q² = 0.59, 0.57, R² = 0.76, 0.40). **b,f.** Variable Importance in Projection (VIP) plots of metabolites driving group separation in males (**b**) and females (**f**). Only metabolites with VIP > 1 were considered significant. **c,g.** Robust volcano plot displaying the upregulated (red) and downregulated (blue) metabolites in the serum of male **(c)** and female **(g)** PD patients with LRRK2 mutation. The absolute fold change value was set at 1.5, while the p-value threshold was defined as <0.05. **d,h**. Pathway enrichment analysis based on ^1^H-NMR serum metabolomics from male **(d)** and female **(h)** gPD patients with LRRK2 mutations compared to sex-matched controls. Bars indicate the number of detected metabolites (hits) per pathway. Pathways were considered significant with Hits > 1, p < 0.05, Holm-adjusted p < 0.05, and FDR < 1. Darker colours denote lower p-values. Pathway analysis was conducted using SMPDB (organism: *Homo sapiens*).

In males, VIP analysis showed that group separation was primarily influenced by amino acids like L-tryptophan, L-glutamic acid, L-asparagine, isoleucine and L-ornithine, as well as metabolites affecting mitochondrial energy homeostasis and lipid metabolism, such as malonate, succinate, and choline (**Figure 2b**). A robust volcano plot in male cases confirmed significant downregulation of L-glutamic acid, malonate, and choline, along with upregulation of L-tryptophan, L-ornithine, and succinate concentrations compared to matched HCs (**Figure 2c, Table S7**).

In females, VIP analysis highlighted alterations in amino acids—such as L-threonine, L-ornithine, and L-serine—and in energy-related metabolites, including pyruvic acid, acetic acid, 3-hydroxybutyric acid, and citric acid, which choline also emerging as a key discriminant (**Figure 2f**). A robust volcano plot substantiated a significant reduction of choline, pyruvic acid and acetic acid, along with an upregulation of L-threonine and 3-hydroxybutyric acid levels in cases compared to HCs (**Figure 2g, Table S7**). Consistent with these results, heatmap analysis further highlighted sex-based differences in metabolite alterations (**Figure S5**).

Consistent with the NMR data, pathway enrichment analysis indicated that male PD-*LRRK2* patients exhibited dysregulation of multiple cellular pathways compared with sex- and age-matched HCs. Specifically, in the patients’ data analysis, significant dysmetabolism of L-tryptophan, L-alanine, L-arginine, L-proline, L-glutamic acid, L-aspartate, glutathione biosynthesis, the urea cycle, and energy homeostasis was observed compared to the control group **(Figure 2d; Table S8).**

In contrast, female patients with PD-*LRRK2* exhibited biochemical disruptions in the metabolism of serine, glycine, methionine, and betaine. Additionally, there were alterations in the metabolism of amino sugars and pyruvate, as well as the biosynthesis of phosphatidylethanolamine and fatty acids. Moreover, mitochondrial dysfunction related to acetyl group transfer pathways was observed when compared to sex-matched HCs **(Figure 2h; Table S9).**

Together, these findings indicate that LRRK2-associated metabolic alterations differ between males and females when each PD group is compared with its respective controls. These findings are consistent with previous metabolomic studies that highlight systemic changes in PD-*LRRK2* patients^48–51^. Importantly, our results also support earlier NMR-based evidence showing altered levels of amino acids and glucose metabolism intermediates in male PD-*LRRK2* patients compared to HCs ^52^.

### PARK2, PINK1, and PARK7 mutations are associated with sex-specific serum metabolomic profiles in PD patients compared to healthy controls

The *PARK2*, *PINK1*, and *PARK7* genes play crucial and complementary roles in maintaining mitochondrial quality by coordinating the identification and removal of damaged mitochondria through the process of mitophagy ^53^. Although these genes are implicated in the development of PD, serum metabolomic data from *PARK2*, *PINK1*, and *PARK7* mutation carriers remain scarce, especially in studies stratified by sex.

Given the partially distinct biological functions of these genes, where PARK2 and PINK1 mainly regulate mitochondrial quality control and mitophagy, whereas PARK7 is more strongly associated with oxidative stress response pathways ^54^, we conducted multivariate analyses (MVA) by grouping the carriers accordingly. For this analysis, we included 14 patients with *PARK2* mutations, 7 with *PINK1* mutations, and 16 with *PARK7* mutations. Three male participants—one with mutations in both *PINK1* and *PARK7*, and two with mutations in both *PARK2* and *PARK7*—were excluded from the analysis, as their combined genotypes complicated meaningful comparisons.

MVA did not show clear differences in the metabolic outcomes among the three genetic subgroups investigated (**Figure S6**). Given the absence of distinct subgroup-specific patterns, heterozygous carriers of variants in *PARK2*, *PINK1*, and *PARK7* were combined into a single category for the main analyses. We compared serum metabolomic profiles from male patients with PD (N=22) and female patients (N=18) carrying mutations in *PARK2*, *PINK1*, or *PARK7* with age- and sex-matched healthy controls (HCs) comprising 23 males and 25 females.

Unsupervised PCA (**Figure S3e,f**) and PLS-DA, stratified by sex, demonstrated a distinct separation between patients and controls. The PLS-DA models yielded Q² values of 0.64 for males and 0.62 for females on PC1, and R² values of 0.80 for males and 0.65 for females on PC1 (**Figure 3a,e**). The models’ performance was further corroborated through validation using Random Forests (**Table S3**).

**Figure 3.**
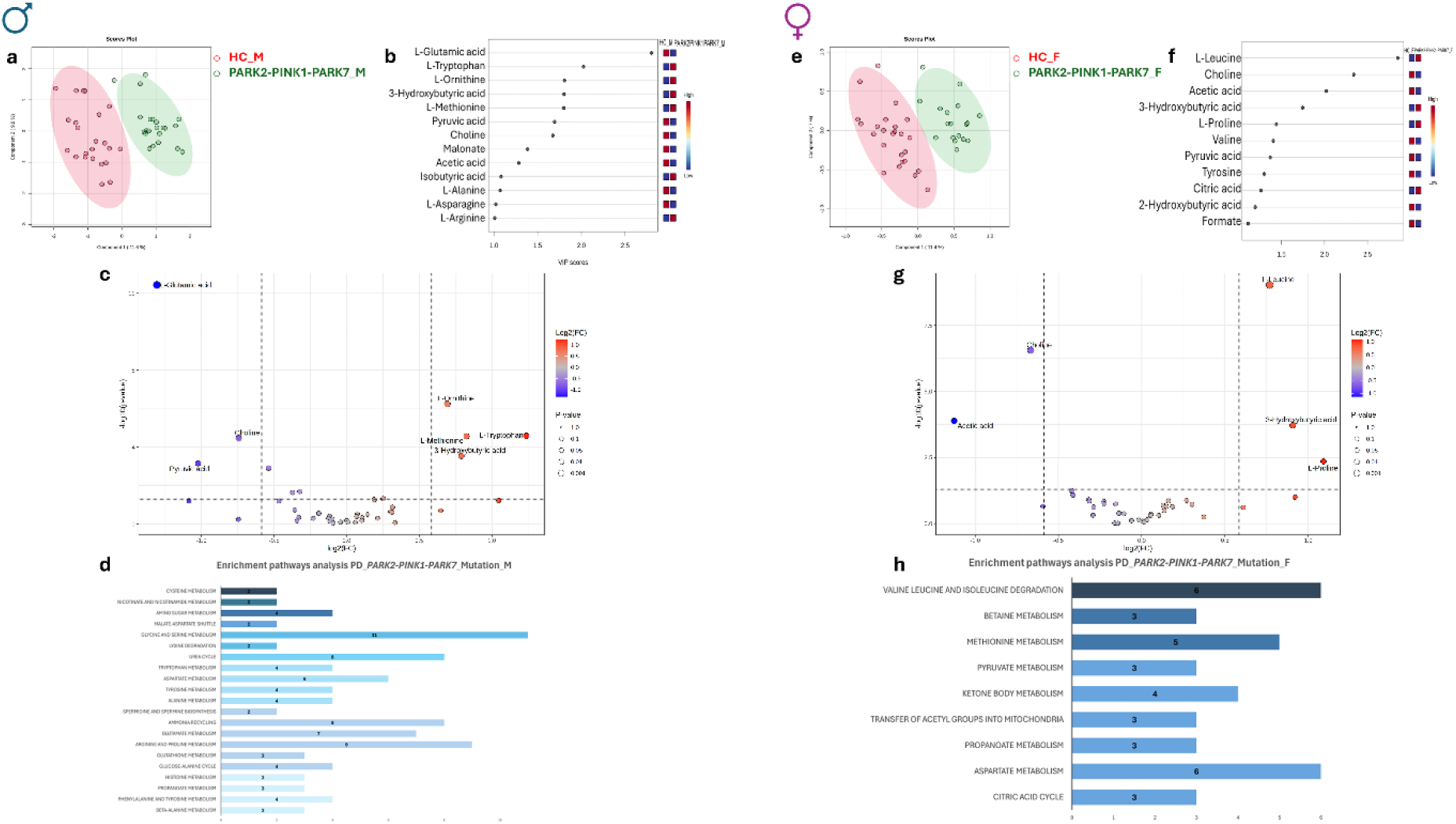
PARK2-PINK1-PARK7 mutations in PD patients impacts serum metabolome profile in a sex-dependent manner. **a, e.** PLS-DA score plots of serum metabolite levels from genetic Parkinson’s disease (gPD) patients carrying PARK2-PINK1-PARK7 mutations (males: 22 patients, red; 23 healthy controls, green; **panel a**; females: 18 patients, red; 25 healthy controls, green; **panel e**). Variance explained: PC1 = 11.6%, PC2 = 9.9% (males); PC1 = 11.4%, PC2 = 7.0% (females). Models validated by 10-fold CV (males: accuracy = 0.95, 0.90; Q² = 0.64, 0.69; R² = 0.80, 0.40 females: accuracy = 0.90, 0.83; Q² = 0.62, R² = 0.65, 0.50 for both components). **b,f.** Variable Importance in Projection (VIP) plots of metabolites driving group separation in males (**b**) and females (**f**). Only metabolites with VIP > 1 were considered significant. **c,g.** Robust volcano plot displaying the upregulated (red) and downregulated (blue) metabolites in the serum of male **(c)** and female **(g)** PD patients with PARK2-PINK1-PARK7 mutation. The absolute fold change value was set at 1.5, while the p-value threshold was defined as <0.05. **d,h**. Pathway enrichment analysis based on ^1^H-NMR serum metabolomics from male **(d)** and female **(h)** gPD patients with *PARK2-PINK1-PARK7* mutations compared to sex-matched controls. Bars indicate the number of detected metabolites (hits) per pathway. Pathways were considered significant with Hits > 1, p < 0.05, Holm-adjusted p < 0.05, and FDR < 1. Darker colours denote lower p-values. Pathway analysis was conducted using SMPDB (organism: *Homo sapiens*).

In male patients, the VIP analysis identified L-glutamic acid as the primary molecule that distinguishes them from HCs. Additional distinctions were found from other amino acids, including L-tryptophan, L-ornithine, L-argine, L-asparagine, L-alanine, and L-methionine. Furthermore, metabolites that affect energy homeostasis and membrane biogenesis, such as 3-hydroxybutyric acid, pyruvic acid, acetic acid, isobutyric acid, and choline, also played a significant role **(Figure 3b)**. A robust volcano plot analysis revealed significantly decreased levels of L-glutamic acid, choline, and pyruvic acid, alongside increased levels of L-ornithine, L-tryptophan, L-methionine, and 3-hydroxybutyric acid in male patients compared to matched controls (**Figure 3c, Table S10**).

In female patients, the VIP analysis highlighted the significant ability of L-leucine and choline to differentiate these patients from control subjects, with additional contributions from acetic acid, 3-hydroxybutyric acid, and L-proline (**Figure 3f**). A robusto volcano plot analysis confirmed a significant downregulation of choline and acetic acid, along with an upregulation of L-leucine, 3-hydroxybutyric acid, and L-proline levels in female patients compared to control subjects (**Figure 3g; Table S10)**. Heatmap analysis further showed differences in serum metabolite profiles between cases and controls (**Figure S7**).

Pathway enrichment analysis in male patients with *PARK2*, *PINK1*, *PARK7* mutations, revealed dysregulation in various metabolic processes. The most significantly affected pathways include glycine/serine, tryptophan, aspartate, tyrosine/phenylalanine, alanine, glutamate, arginine/proline, and histidine metabolism. Additionally, there were alterations in glutathione biosynthesis, amino sugar and propionate metabolism, along with nitrogen pathways such as the urea cycle and ammonia recycling (**Figure 3d; Table S11**). In contrast, female patients showed dysregulation primarily in methionine and aspartate metabolism, as well as in BCAA degradation. Furthermore, these patients exhibited serum alterations linked to ketone body metabolism, pyruvate metabolism, the citric acid cycle, and the transfer of acetyl groups into mitochondria (**Figure 3h; Table S12**).

Altogether, our findings highlight a sex-dependent effect on the serum metabolome in patients carrying *PARK2*-*PINK1*-*PARK7* mutations compared to sex-matched controls. In particular, the alterations observed in amino acid and ketone-body metabolism are consistent with previous reports in PD-*PARK2* cohorts ^55,56^. Moreover, disruptions in the urea cycle and other energy-related pathways, including the glucose–alanine cycle, align with the established roles of these genes in regulating mitochondrial function ^57,58^.

### Mutations in TMEM175 are linked to differences in serum metabolomic profiles between male and female PD patients and healthy controls

*TMEM175* encodes a lysosomal cation channel essential for lysosomal homeostasis ^59–61^, and both common and rare variants in this gene are strongly associated with PD ^10^. To assess the metabolic consequences of *TMEM175* mutations, we performed MVA of NMR-based serum metabolomic profiles from PD patients carrying *TMEM175* variants (20 males, 12 females) and age- and sex-matched HCs (23 males, 25 females).

Notably, unsupervised PCA **(Figure S3g, h)** and PLS-DA revealed clear separation between PD-*TMEM175* patients and age- and sex-matched HCs in both sexes, with PC1 Q² and R² values of 0.71 and 0.81 in males and 0.58 and 0.68 in females **(Figure 4a, e).** The classification models were further validated using a Random Forest approach (**Table S3**).

**Figure 4.**
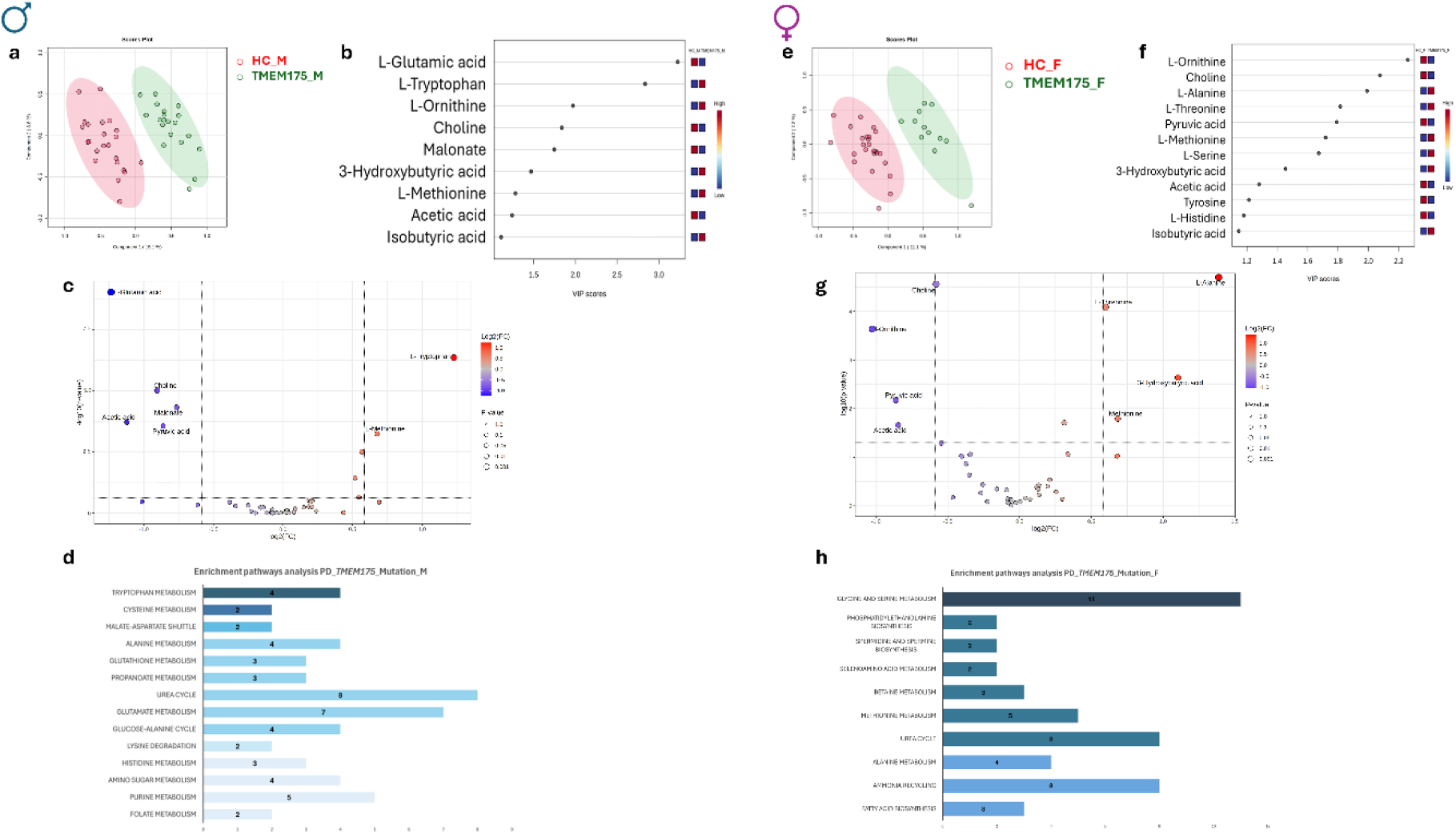
TMEM175 mutations in PD patients impacts serum metabolome profile in a sex-dependent manner. **a, e.** PLS-DA score plots of serum metabolite levels from genetic Parkinson’s disease (gPD) carrying TMEM175 mutations (males: 20 patients, red; 23 healthy controls, green; **panel a**; females: 12 patients, red; 25 healthy controls, green; **panel e**). Variance explained: PC1 = 15.1%, PC2 = 8.6% (males); PC1 = 11.1%, PC2 = 7.2% (females). Models validated by 10-fold CV (males: accuracy = 0.93 for both components; Q² = 0.71, 0.78; R² = 0.81, 0.88 females: accuracy = 0.94, 0.97; Q² = 0.58, 0.68, R² = 0.68, 0.76). **b,f.** Variable Importance in Projection (VIP) plots of metabolites driving group separation in males (**b**) and females (**f**). Only metabolites with VIP > 1 were considered significant. **c,g.** Robust volcano plot displaying the upregulated (red) and downregulated (blue) metabolites in the serum of male **(c)** and female **(g)** PD patients with TMEM175 mutation. The absolute fold change value was set at 1.5, while the p-value threshold was defined as <0.05. **d,h**. Pathway enrichment analysis based on ^1^H-NMR serum metabolomics from male **(d)** and female **(h)** gPD patients with *TMEM175* mutations compared to sex-matched controls. Bars indicate the number of detected metabolites (hits) per pathway. Pathways were considered significant with Hits > 1, p < 0.05, Holm-adjusted p < 0.05, and FDR < 1. Darker colours denote lower p-values. Pathway analysis was conducted using SMPDB (organism: *Homo sapiens*).

In males, VIP analysis identified L-glutamic acid as the metabolite most significantly distinguishing patients from controls. Additional amino acids contributing to group separation included L-tryptophan and L-methionine. Furthermore, choline, followed by malonate isobutyric acid, and acetic acid, delineates a metabolic signature involving phospholipid and energy-related metabolites (**Figure 4b**). A robust volcano plot indicated that male patients with *TMEM175* mutations exhibited lower levels of L-glutamic acid, choline, malonate, acetic acid, and pyruvic acid compared to the control group. In contrast, these patients showed increased levels of L-tryptophan and L-methionine when compared to the matched controls (**Figure 4c; Table S13**).

In females, the VIP analysis identified L-ornithine, L-alanine, L-threonine, Tyrosine, L-serine, L-Histidine and L-methionine as the most notable distinguishing molecules between patients with PD-*TMEM175* and control subjects **(Figure 4f)**. Additional contributors included choline, isobutyric acid pyruvic acid, and acetic acid **(Figure 4f).**

Volcano plot indicated reduced levels of choline, L-ornithine, pyruvic acid and acetic acid, while increased levels of L-alanine, L-threonine, 3-hydroxybutyrate and L-methionine, in female PD*-TMEM175* patients compared to HCs (**Figure 4g; Table S13**). Heatmap analysis confirmed pronounced sex-dependent patterns in metabolite alterations between cases and controls (**Figure S8**).

Consistent with NMR data, pathway enrichment analysis in male patients showed disruptions in tryptophan, histidine, alanine, and L-glutamic acid metabolism. It also identified disturbances in purine metabolism and glutathione biosynthesis compared to HCs **(Figure 4d; Table S14)**.

In female *TMEM175* patients, glycine, serine, methionine, and alanine metabolism were dysregulated **(Figure 4h; Table S15)**. Additional disruptions involved betaine metabolism, phosphatidylethanolamine biosynthesis, the urea cycle, ammonia recycling, and fatty acid biosynthesis compared with matched controls **(Figure 4h; Table S15)**.

These findings demonstrate that sex influences the serum metabolic profile of PD patients carrying *TMEM175* mutations compared with sex-matched HCs. In addition, the present data align with previous lipidomic–metabolomic research in *TMEM175* carriers, which identified alterations in fatty acids, sphingolipids, and glycerophospholipids ^24^.

### ¹H-NMR metabolomics reveals common and distinct metabolite variations among patients with different genetic mutations related to PD

To deliver a comprehensive synthesis of the extensive data from our multivariate metabolomic analyses, we generated an UpSet plot summarising discriminant metabolites identified by VIP scores in comparisons between gPD patients, stratified by sex, and their respective sex-matched HCs (**Figure 5**).

**Figure 5.**
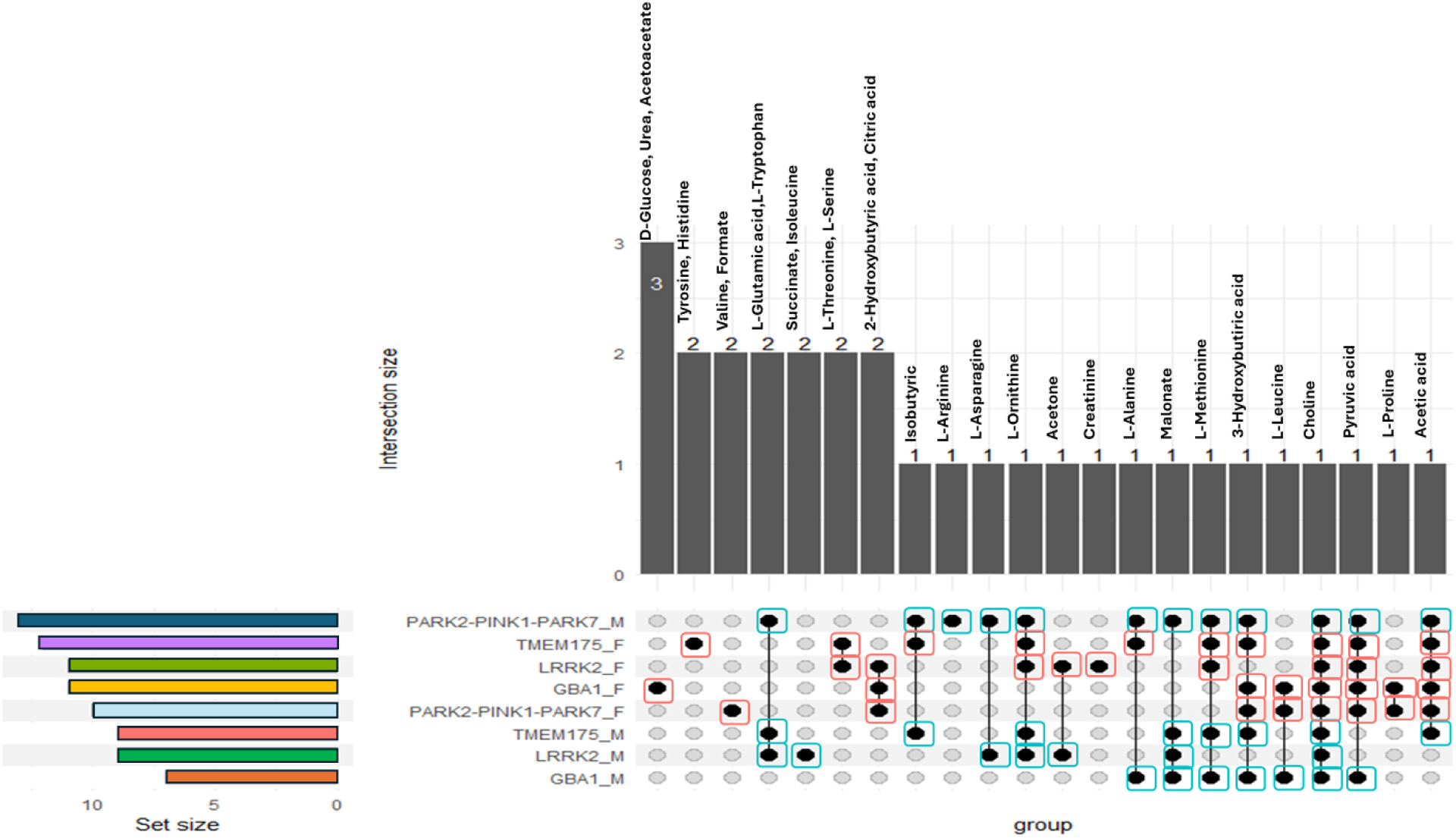
UpSet-Based Integration of VIP Features Reveals Sex-Driven Metabolic Divergence Across gPD Mutations. The UpSet plot, generated using the Upset R package, illustrates the intersections among VIP-selected serum metabolites in male and female gPD patients stratified by pathogenic mutations. The upper bar chart shows the size of each metabolite intersection, while the matrix below indicates the specific genotype–sex groups contributing to each combination. Horizontal bars on the left represent the total number of discriminant metabolites identified within each subgroup. This visualisation summarises the shared and distinct metabolite sets across GBA1, LRRK2, PARK2–PINK1–PARK7, and TMEM175 mutation carriers in both sexes.

The UpSet plot emphasized choline as the only discriminant metabolite across all genotypes and in both sexes, underscoring its central role in distinguishing the serum metabolomic profiles of gPD patients from those of HCs and supporting the presence of neurotransmitter imbalance and phospholipid dysmetabolism in PD patients ^62–64^.

In males, several metabolites recurred across multiple genotypes: malonate emerged as a discriminant metabolite shared by all male subgroups, corroborating a mitochondrial metabolism impairment, while glutamate and tryptophan dysregulation were common to the *PARK2*–*PINK1*–*PARK7*, *TMEM175*, and *GBA1* cohorts.

In contrast, female patients showed a pattern characterised by biomolecules involved in energy metabolism, such as 2-hydroxybutyrate and citric acid, which recurred across the *PARK2*–*PINK1*–*PARK7*, *LRRK2*, and *GBA1* subgroups (**Figure 5**).

Overall, the UpSet plot provides a concise visual summary of a key finding from our analyses: numerous discriminant metabolites are shared across genotypes; however, their recurrence patterns differ markedly between sexes. This highlights that sex is a primary factor influencing the serum metabolomic profile of gPD patients relative to sex-matched HCs.

### ¹H-NMR metabolomics identifies indistinguishable serum profiles across different PD-linked genetic mutations

To further investigate whether patients with gPD, categorized by genotype and sex, displayed distinct metabolic profiles, we performed a multivariate PLS-DA. This analysis directly compared the serum metabolomic profiles of all genetically defined PD subgroups, with and without sex stratification (**Figure 6a-c**). Notably, the PLS-DA revealed no genotype-specific differentiation, indicating that there is a common set of circulating metabolic disturbances in gPD that are largely independent of the specific pathogenic variant.

**Figure 6.**
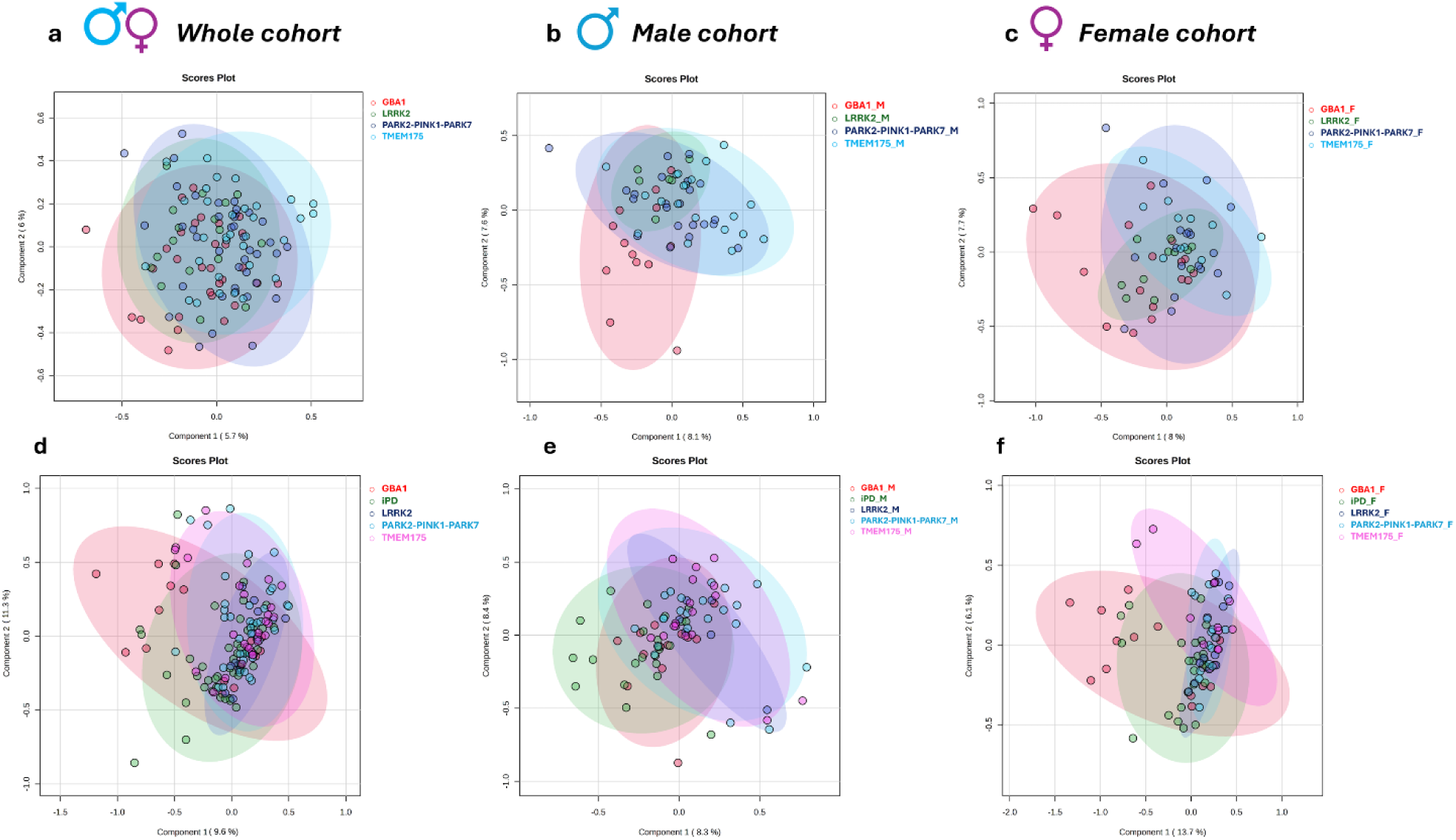
PD patients carrying different pathogenic mutations exhibited indistinguishable serum metabolomic profiles across sexes and when compared with idiopathic cases. (**a**) The PLS-DA score plot of serum metabolite levels from 119 genetic Parkinson’s disease (gPD) patients shows no clear separation among genotypes. Groups included: GBA1 (red, *n* = 30), PARK2–PINK1–PARK7 (blue, *n* = 40), TMEM175 (light blue, *n* = 32), and LRRK2 (green, *n* = 17). Variance explained: PC1 = 5.7%, PC2 = 6.0%. Ten-fold CV: accuracy = 0.27 (PC1) and 0.35 (PC2); Q² = – 0.13, –0.42; R^2^ = 0.30, 0.42. **(b)** PLS-DA score plot of male gPD patients (*n* = 61; GBA1 *n* = 13, PARK2–PINK1–PARK7 *n* = 22, TMEM175 *n* = 20, LRRK2 *n* = 6). Variance explained: PC1 = 8.1%, PC2 = 7.6%. Ten-fold CV: accuracy = 0.38 (PC1), 0.33 (PC2); Q² = –0.37, –0.69; R^2^ = 0.52, 0.43. **(c)** PLS-DA score plot of female gPD patients (*n* = 58; GBA1 *n* = 17, PARK2–PINK1–PARK7 *n* = 18, TMEM175 *n* = 12, LRRK2 *n* = 11). Variance explained: PC1 = 8.0%, PC2 = 7.7%. Ten-fold CV: accuracy = 0.29 (PC1), 0.30 (PC2); Q² = –0.42, –0.85; R^2^ = 0.31, 0.48. **(e)** PLS-DA model comparing 119 gPD patients (stratified by genotype) with 48 idiopathic PD (iPD) patients. Variance explained: PC1 = 9.6%, PC2 = 11.3%. Ten-fold CV: accuracy = 0.35 (PC1), 0.33 (PC2); Q² = –0.04, –0.13; R² = 0.14, 0.22. **(f)** The male-only comparison between gPD and iPD patients. Variance explained: PC1 = 8.3%, PC2 = 8.4%. Ten-fold CV: accuracy = 0.39 (PC1), 0.38 (PC2); Q² = –0.11, –0.18; R² = 0.29, 0.44. (g) The male-only comparison between gPD and iPD patients. Variance explained: PC1 = 13.7%, PC2 = 6.1%. Ten-fold CV: accuracy = 0.37 (PC1), 0.39 (PC2); Q² = –0.07, –0.26; R² = 0.20, 0.38.

We next assessed whether this shared metabolic profile extended to idiopathic PD, which represents the predominant clinical phenotype. To this end, we compared the serum metabolomic profiles of gPD patients (stratified by genotype and sex) with those of sex-matched iPD individuals (23 males and 25 females) **(Figure 6e-g)**. Strikingly, our analysis revealed overlapping metabolic signatures between iPD and gPD patients, supporting the hypothesis of a shared metabolic dysregulation underlying the pathophysiology of PD, regardless of genetic status.

### Integrated modeling indicates that sex is the primary factor driving dysregulation of glutamate, serine, and citric acid in the blood of patients with gPD compared to matched controls

Finally, given that gPD patients with different mutations showed indistinguishable serum metabolomic profiles regardless of sex, we treated all gPD patients as a single group to formally test whether the effect of gPD diagnosis on serum metabolite levels differs between males and females. A linear model with a group × sex interaction term was fitted in the full cohort (HC + gPD; n = 166), adjusting for age. After Benjamini-Hochberg FDR correction across 41 outcomes (43 metabolites excluding methanol and ethanol), two metabolites reached statistical significance: L-glutamic acid (β = −0.680, 95% CI [−1.043, −0.317], p_FDR = 0.0098) and L-serine (β = −0.734, 95% CI [−1.201, −0.267], p_FDR = 0.0424) (**Figure 7**). For L-glutamic acid, the reduction associated with gPD diagnosis was significantly greater in males than in females (**Table S16**). For L-serine, the interaction was qualitative: gPD females showed higher levels relative to HC females, while gPD males showed lower levels relative to HC males (β = −0.734, 95% CI [−1.201, −0.267], p_FDR = 0.0424; **Table S16**). Citric acid showed a borderline interaction (p_FDR = 0.070; **Table S16**).

**Figure 7.**
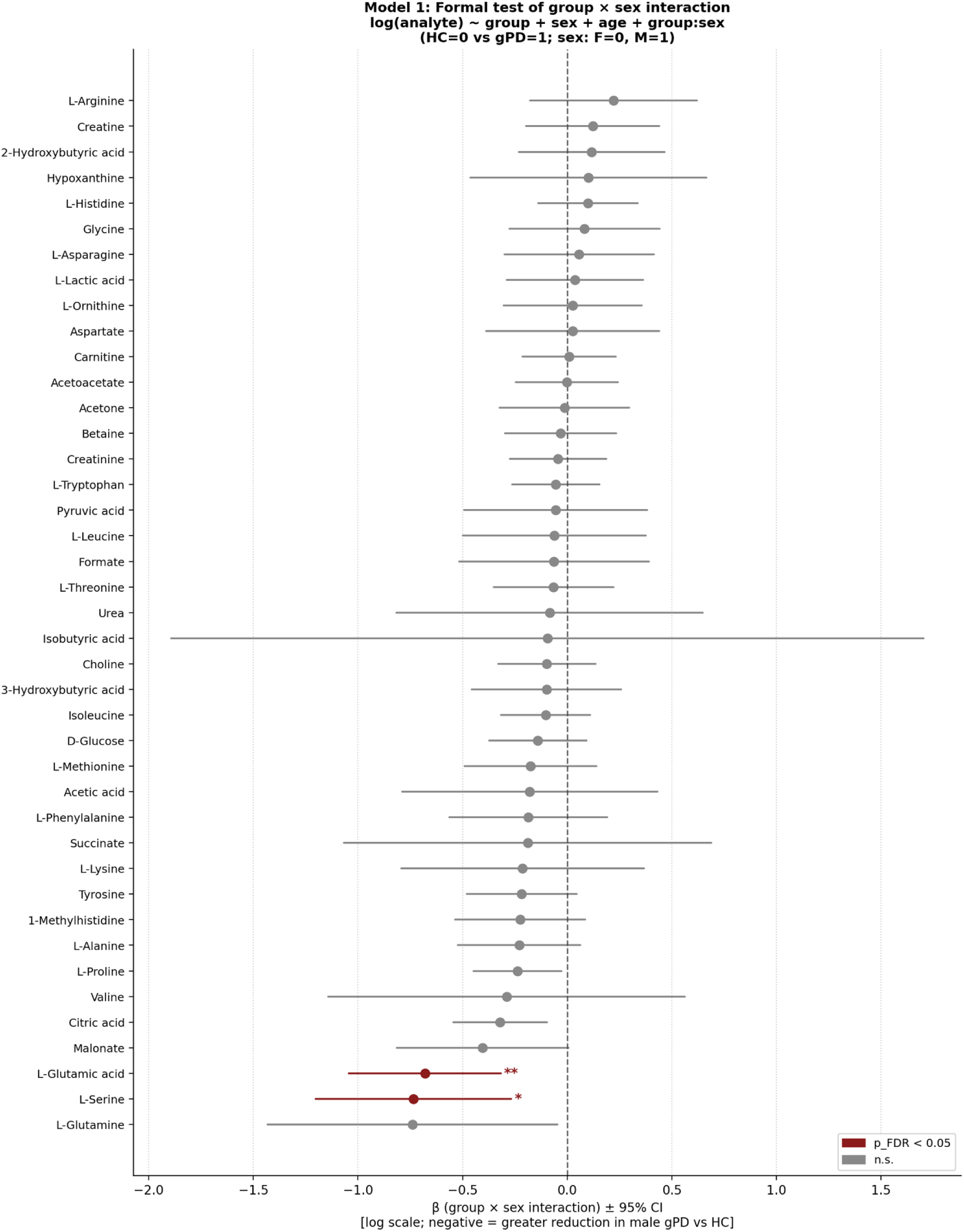
Formal testing of group × sex interaction on serum metabolite levels in the full cohort. Forest plot showing the group × sex interaction coefficients (β) and 95% confidence intervals from OLS linear models fitted separately for each of the 41 serum amino acid and metabolite outcomes (HC + gPD; n = 166). Model formula: log(analyte) ∼ group + sex + age + group:sex, where group = 0 (HC) or 1 (gPD) and sex = 0 (female) or 1 (male). All metabolite concentrations were natural log-transformed prior to modelling. A negative β indicates a greater reduction associated with gPD in males relative to females. Inference based on HC3 robust standard errors; p-values corrected using Benjamini-Hochberg FDR across 41 outcomes. Red: p_FDR < 0.05; grey: not significant.

### 1H-NMR analysis reveals differences in the serum metabolome profiles between male and female patients with familial PD, stratified by specific mutations

To investigate the influence of sex differences on serum metabolomic profiles in patients carrying pathogenic genetic variants, here we analyzed metabolic features in independent cohorts of HCs and gPD patients, stratified by sex and genotype.

Notably, PLS-DA did not reveal any separation between male and female HCs (PC1; Q² = 0.003; R² = 0.05; accuracy = 0.58), confirming the metabolic homogeneity of the control group (**Figure 8a**).

**Figure 8.**
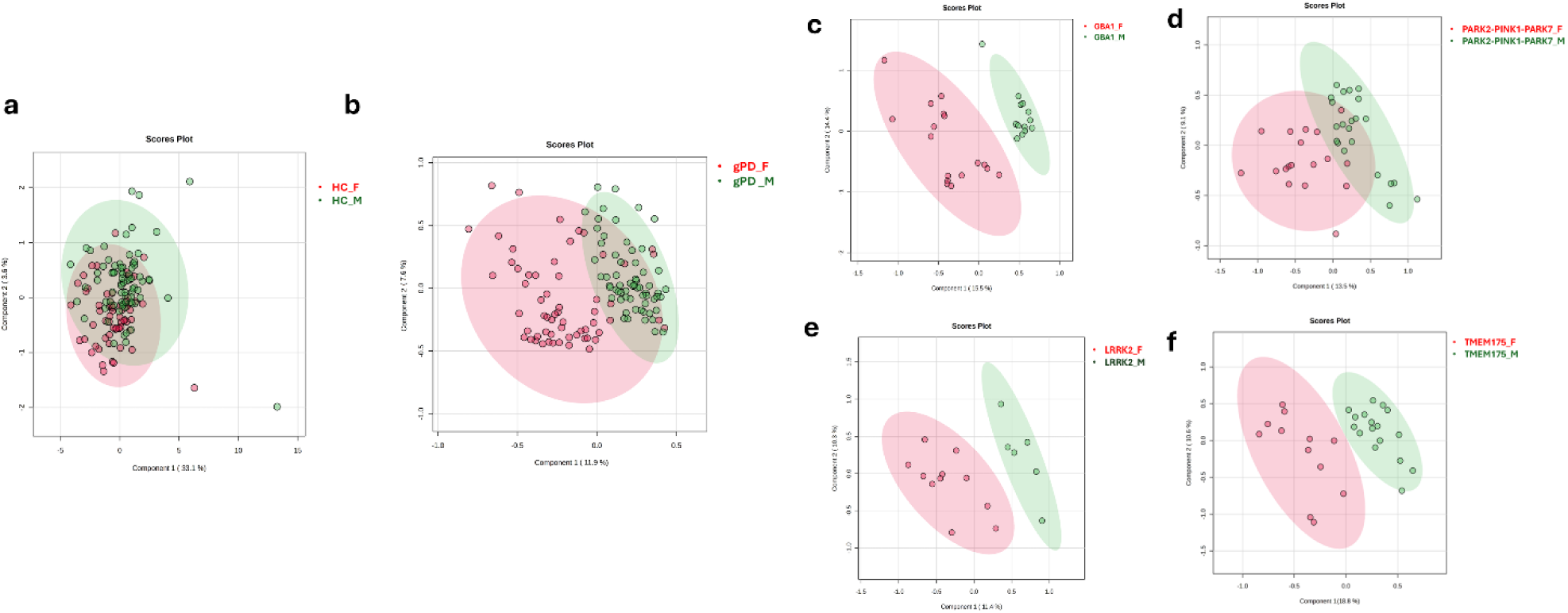
Sex-dependent differences in serum metabolomic profiles across gPD genotypes. **a,** PLS-DA of serum metabolites in male (n = 23) and female (n = 23) healthy controls. The models explained 33.1% (PC1) and 3.6% (PC2) of the variance. Ten-fold cross-validation yielded accuracies of 0.58 and 0.57, Q² values of 0.003 and –0.19, and R² values of 0.05 and 0.25. Negative Q² values indicate the absence of sex-related metabolic separation in healthy individuals. **b,** PLS-DA of the entire gPD cohort (13 males, 17 females). The models explained 11.9% (PC1) and 7.6% (PC2). Cross-validation yielded accuracies of 0.81 and 0.88, Q² values of 0.39 and 0.43, and R² values of 0.53 and 0.61. Group separation was supported by PERMANOVA (p = 0.002, F = 6.27). Random Forest classification showed an OOB error of 0.06. **c,** PLS-DA of GBA1-associated PD patients (13 males, 17 females). The models explained 15.5% (PC1) and 14.4% (PC2). Cross-validation yielded accuracies of 0.86 and 0.97, Q² values of 0.50 and 0.68, and R² values of 0.69 and 0.85. PERMANOVA confirmed group separation (p = 0.005, F = 6.31). Random Forest classification showed an OOB error of 0.03. **d,** PLS-DA of LRRK2-associated PD patients (6 males, 11 females). The models explained 11.4% (PC1) and 10.3% (PC2). Cross-validation yielded accuracies of 0.55 and 0.76, Q² values of 0.30 and 0.36, and R² values of 0.49 and 0.56. PERMANOVA indicated borderline separation (p = 0.05, F = 2.90). Random Forest analysis was not performed due to an unbalanced sample size.**e,** PLS-DA of patients carrying PARK2, PINK1 or PARK7 mutations (22 males, 18 females). The models explained 13.5% (PC1) and 9.1% (PC2). Cross-validation yielded accuracies of 0.77 and 0.72, Q² values of 0.30 and 0.55, and R² values of 0.52 and 0.67. PCA-based permutation testing confirmed separation (p = 0.001, F = 7.82). Random Forest classification showed an OOB error of 0.10. **f** PLS-DA of TMEM175-associated PD patients (20 males, 12 females). The models explained 18.8% (PC1) and 10.6% (PC2). Cross-validation yielded accuracies of 0.72 and 0.66, Q² values of 0.60 and 0.63, and R² values of 0.75 and 0.80. PCA-based permutation testing confirmed separation (p = 0.03, F = 3.78). Random Forest classification showed an OOB error of 0.10.

In contrast, MVA performed on the entire gPD cohort, without stratification by genotype, identified a clear distinction between male and female patients (PC1; Q² = 0.39; R² = 0.53; accuracy = 0.81) (**Figure 8b**). Additionally, PLS-DA stratified by mutation revealed sex-associated differences across all genetic subgroups, including *GBA1* (PC1; Q² = 0.50; R² = 0.69; accuracy = 0.86), *LRRK2* (PC1; Q² = 0.30; R² = 0.49; accuracy = 0.55), *PARK2–PINK1–PARK7* (PC1; Q² = 0.30; R² = 0.52; accuracy = 0.77), and *TMEM175* (PC1; Q² = 0.60; R² = 0.75; accuracy = 0.72) (**Figure 8c–f**).

These findings were further supported by PERMANOVA validation and error-rate estimates, indicating that sex modulates metabolomic serum signatures across all genetic backgrounds. Univariate analysis using a robust volcano plot also demonstrated a consistent increase in glutamate levels in females compared with males across all mutation groups (**Figure S9b,d,f,h,l**), thereby corroborating the results from the linear model.

The central role of glutamate was further reinforced by VIP analysis, which identified this metabolite as a key discriminator across all sex-stratified genotype subgroups (**Figure S9c,e,g,i**). Additionally, in the *GBA1*-mutated cohort, univariate analysis also indicated reduced levels of urea and 3-hydroxybutyrate, consistent with the close association of this gene mutation with alterations in cellular energetics ^65^ (**Figure S9d**).

Overall, our findings illustrate a notable sexual dimorphism in the serum metabolomic profiles of gPD patients compared to HCs and emphasize the role of specific metabolites, particularly glutamate, in distinguishing male and female subjects.

## Discussion

Metabolomics provide a powerful framework for capturing dynamic biochemical changes associated with both physiological and pathological states by quantifying small-molecule intermediates across interconnected metabolic pathways ^66–69^. Circulating plasma and serum metabolites reflect the integrated output of environmental exposures, organ-specific physiology, and hormonal regulation, offering a systemic readout of disease-related metabolic disturbances ^70–73^. In the context of PD, alterations in circulating metabolites have been repeatedly linked to mitochondrial dysfunction, impaired energy metabolism, oxidative stress, and neuroinflammation, supporting the view of PD as a multisystem disorder ^74–78^.

Although previous metabolomic studies have identified serum or plasma “fingerprints” capable of distinguishing PD patients from HCs, inconsistencies in the direction and magnitude of these alterations have impeded a clear biological interpretation of the findings ^35^. In particular, only a minority of metabolomic studies have examined the contribution of sex ^36,79,80^ and genetic status ^24,39,81–83^ despite their well-established impact on PD epidemiology, clinical heterogeneity, and treatment response.

To address this gap, the present study investigates the impact of mutations in the *GBA1*, *LRRK2*, *PARK2*, *PINK1*, *PARK7*, and *TMEM175* genes on serum metabolomics in male and female patients with PD. To carry out our analysis, we employ ^1^H-NMR-based metabolomics, a robust and highly reproducible approach for metabolic profiling that has been widely applied in studies of this neurodegenerative disorder ^36,41,84–94^.

Consistent with the established biological roles of the genes analysed, the metabolic alterations observed across our genetic subgroups converge on biochemical pathways central to PD pathogenesis. Carriers of *PARK2*, *PINK1*, and *PARK7* variants showed metabolic alterations consistent with impaired mitophagy and mitochondrial quality control, including higher concentrations in ketone bodies, such as 3-hydroxybutyrate, and reduced levels of glycolytic metabolites, such as pyruvate, suggesting compensatory metabolic rewiring in response to defective energy production ^95,96^. Although *PARK7* also contributes to mitochondrial homeostasis, its established role in antioxidant defence and oxidative stress responses ^97,98^ may additionally explain the observed perturbations in glutathione-related and redox-associated metabolic pathways.

Similarly, *LRRK2* mutation carriers exhibited alterations in central carbon metabolites, highlighted by an upregulation of succinate and a reduction of pyruvic acid, supporting the presence of mitochondrial metabolic dysfunction consistent with previous evidence implicating LRRK2 in the regulation of mitochondrial fusion–fission dynamics, intracellular trafficking, autophagy, and calcium homeostasis ^99^.

In contrast, the metabolic profiles observed in *GBA1* and *TMEM175* carriers were more consistent with lysosomal dysfunction and altered intracellular homeostasis. In *GBA1* carriers, alterations in ketone bodies and amino acid metabolism may reflect broader metabolic adaptations associated with lysosomal dysfunction ^100^. Given the central role of lysosomes in autophagic degradation and nutrient recycling, reduced lysosomal efficiency may impair amino acid homeostasis and indirectly compromise mitochondrial quality control. Such disturbances are known to induce secondary mitochondrial stress ^101^ and may promote a compensatory shift in cellular energy utilization toward alternative substrates, which could contribute to the observed changes in ketone body metabolism. Likewise, in *TMEM175* carriers, perturbations in metabolites related to amino acid and energy metabolism, including reduced pyruvate and choline levels, suggest altered cellular energy homeostasis. Given the role of TMEM175 in lysosomal ion homeostasis and pH control ^23^, these metabolic changes may be compatible with impaired lysosomal activity and downstream effects on mitochondrial energy metabolism.

Across all genetic groups, the recurrent perturbations in metabolites impacting energy homeostasis, amino acid pathways, and lipid metabolism suggest that distinct genetic defects may converge on shared cellular mechanisms, ultimately resulting in common systemic metabolic alterations observed in PD, despite the underlying genetic heterogeneity. Notably, choline was the only metabolite consistently altered across all genetic variants, showing increased levels in male and female *GBA1* carriers and decreased levels in all other groups, irrespective of sex. Although further studies are needed to clarify the mechanisms underlying these divergent directions across genotypes, alterations in choline levels are particularly relevant given its involvement in several fundamental biochemical pathways, including neurotransmitter synthesis, lipid and one-carbon metabolism, methyl-group donation, lipid transport, and membrane signaling ^102–104^.

Our NMR-based analysis also revealed distinct metabolic differences between each sex-stratified PD mutation subgroup and their age- and sex-matched HCs. Importantly, these alterations exhibited sex-associated patterns that were evident in patients but largely absent in HCs, suggesting that sex may modulate the metabolic expression of PD-linked mutations rather than reflecting baseline differences between males and females.

A notable finding from our study is that male patients, irrespective of the specific mutation, consistently exhibited alterations in circulating malonate levels. Malonate is one of the most potent competitive inhibitors of succinate dehydrogenase (complex II) and a regulator of acidic lipid synthesis ^105,106^, making its dysregulation a relevant indicator of mitochondrial and metabolic stress. In addition, male carriers of *PARK2*, *PINK1*, *PARK7*, *TMEM175* and *GBA1* mutations showed a reduction in circulating glutamate accompanied by increased tryptophan levels, indicating a recurrent alteration in amino-acid-related pathways.

The reduction in glutamate is consistent with previous metabolomic studies of PD ^41,107–110^, except for one investigation conducted in a small, non-stratified Indian cohort (PD = 17, HCs = 22) reporting increased levels in patients ^111^. Such discrepancies likely reflect differences in analytical platforms, cohort composition and, importantly, the absence of sex- or genotype-stratified analyses. In our stratified framework, glutamate alterations were not uniform across the PD population but emerged consistently within specific genetic subgroups and were restricted to male patients, in line with our previous observations in PD carriers of non-pathogenic variants ^112^. This underscores the importance of incorporating biological variables such as sex and genotype when interpreting metabolic signatures in PD.

The analysis of sexual dimorphism, using a linear model with sex-interaction term, further emphasized the critical role of glutamate. Our findings showed that the reduction in glutamate associated with gPD diagnosis was significantly more pronounced in males than in females when each group was compared with its respective sex-matched HCs.

Overall, our metabolomic results indicate that glutamate is among the metabolites most strongly influenced by sex in the context of PD. Beyond its role as an excitatory neurotransmitter, glutamate is a key metabolic intermediate involved in the tricarboxylic acid cycle and in pathways regulating cellular redox balance, including glutathione biosynthesis. Therefore, the sex-associated reduction in glutamate may reflect differential metabolic responses to PD-linked pathology, consistent with broader evidence of sex-dependent vulnerability in mitochondrial and redox pathways ^113,114^.

In the female cohort, our analyses showed a consistent decrease in circulating levels of 2-hydroxybutyrate, alongside a corresponding increase in citrate. This was observed in patients with *LRRK2*, *GBA1*, and *PARK2–PINK1–PARK7* mutations compared to HCs. These metabolic alterations may reflect sex-specific vulnerability in redox homeostasis and mitochondrial energy metabolism, given the role of 2-hydroxybutyrate in glutathione turnover and oxidative stress responses, and the involvement of citrate in tricarboxylic acid cycle function and mitochondrial bioenergetics.

In support of these findings, linear model analysis pointed to citric acid as a metabolite potentially involved in sex-dependent metabolic variation in PD. Although the sex × genotype interaction only approached significance, the model indicated that citric acid concentrations tend to be higher in females than in males across genetic backgrounds when compared with HCs, consistent with previous evidence of citrate dysregulation in PD ^111,115^.

Moreover, interaction analysis highlights the importance of serine levels in distinguishing sexual dimorphism within PD. Notably, serine concentrations were elevated in female and decreased in male patients when each gPD group was compared with its sex-matched HCs. The sex-dependent modulation of this metabolite is particularly relevant in the context of PD given the central physiological functions of L-serine, which include de novo synthesis from glycolytic intermediates. Despite the clinical relevance of our observations, it remains unclear whether the changes in serum metabolome profiles identified in gPD patients by NMR spectroscopy primarily reflect pathophysiological processes originating in the basal ganglia or arise from peripheral organ dysfunction and systemic metabolic adaptations. In this context, recent Ultra-Performance Liquid Chromatography–Mass Spectrometry (UPLC-MS)-based metabolomic analyses of post-mortem caudate-putamen tissue evaluated a panel of 44 amino acids and related metabolites, revealing selective increases in serine and proline levels together with reduced phosphoethanolamine concentrations in PD brains compared with controls ^116,117^. By integrating these *post-mortem* findings with the results of the current and previous studies showing a broader spectrum of altered serum amino acid concentrations in PD patients relative to controls ^32,41,112,118^, we infer that the disruption of metabolic homeostasis in PD likely involves both the central nervous system and peripheral organs. In this context, it is worth mentioning a recent HPLC-based study from our group showing that late-stage PD patients, with a median disease duration of approximately 23 years, who received subthalamic nucleus deep brain stimulation (STN-DBS) in combination with L-DOPA treatment exhibited normalization of plasma concentrations of glutamatergic system-related D- and L-amino acids and their precursors ^107^. These metabolites, which were found to be altered in both early-stage (L-DOPA-naïve) and mid-stage (L-DOPA-treated) PD patients, reached levels comparable to those observed in HCs ^107^. These findings suggest that a centrally targeted intervention such as STN-DBS may exert effects beyond basal ganglia circuitry, influencing peripheral metabolic homeostasis as reflected by the normalization of plasma amino acid profiles even at advanced stages of the disease. Interestingly, our findings show that serum metabolome profiles are largely overlapping among patients carrying different PD-associated genetic mutations across the entire study cohort, even after stratification by sex. Moreover, our multivariate analysis of NMR data failed to distinguish serum metabolome profiles between gPD and iPD patients, irrespective of sex. We interpret this lack of distinct clustering as indicative of common metabolic disruptions across PD patients, independent of their genetic background. These shared alterations likely reflect underlying biochemical pathways involved in PD, such as mitochondrial dysfunction, disrupted amino acid metabolism, oxidative stress mechanisms, and energy imbalance. Consequently, our study supports the hypothesis that, despite the clinical heterogeneity characteristic of PD, a common systemic metabolic core exists across subtypes and genetic mutations ^112^.

Nonetheless, given the potential biological and clinical implications of these observations, further metabolomic investigations in independent, genetically stratified cohorts—using multiple and complementary analytical approaches—are warranted. Indeed, although NMR is highly reproducible and quantitative, it has lower sensitivity and metabolite coverage compared to mass spectrometry ^119^. Therefore, orthogonal validation using liquid chromatography-mass spectrometry or gas chromatography-mass spectrometry is crucial for confirming and extending our current observations.

One of the main strengths of our study is its integrated design, which is illustrated by a thorough comparative NMR analysis of serum metabolomes in male and female patients with different genetic mutations commonly associated with PD. Additionally, an important aspect of this work is that all comparisons of metabolomic data include cohorts of genetically characterized HCs and idiopathic patients, matched for both sex and age. Importantly, HCs were individuals without neurological disorders or a family history of neurological diseases and were further characterized by Whole Exome Sequencing (WES) to exclude mutations or variants in PD-related genes.

Nevertheless, we recognize certain limitations within our study. Firstly, the cross-sectional design employed did not permit the establishment of causative relationships among serum metabolome alterations, sex differences, and PD-linked genetic mutations. Future studies should consider longitudinal approaches involving larger PD cohorts and adopting a multidimensional methodology, including the assessment of blood biomarkers reflecting brain pathology (e.g., neurofilament light chain), as well as peripheral organ damage and inflammatory processes, to further investigate these associations. Additionally, we acknowledge other limitations: (i) the relatively small sample size limits the generalizability of our findings and hampers the ability to establish causality regarding sex or mutation-specific metabolic variations; (ii) the inclusion of patients undergoing antiparkinsonian therapy may have impacted the metabolomic profiles observed; (iii) grouping heterozygous carriers of variants in recessive PD-associated genes (*PARK2*, *PINK1*, and *PARK7*), while necessary to maintain cohort integrity and statistical power, may have concealed gene-specific biological and metabolic signatures; and (iv) aggregating *GBA1* variant carriers, despite the known functional heterogeneity affecting glucocerebrosidase activity, lysosomal function, and disease severity, may have diminished the capacity to identify variant-specific metabolic effects and disease mechanisms. These limitations emphasize the need for validating our results with larger cohorts and using higher-sensitivity, quantitative omics platforms, such as mass spectrometry.

In conclusion, our study highlights the significant impact of sex on the circulating metabolomic profiles of genetically defined PD patients. In turn, it underscores the necessity of stratifying PD cohorts by sex and genotype to better understand the diverse biochemical landscape of the disease.

## Methods

### Study cohort

#### PD-cohort

This is a case-control observational study. 167 independent and unrelated PD patients (84 males and 83 females; 119 genetic PD and 48 idiopathic cases) were enrolled in this study. PD patients were selected from the Mediterranean Neuromed Institute (MNI)-PD cohort, which includes 804 independent and unrelated patients (501 males; 300 familial and 504 sporadic cases), belonging to the Parkinson’s disease Biobank of the IRCCS Neuromed and of the Institute of Genetics and Biophysics (CNR) ^11,120^. Only individuals aged ≥40 years were included. All the subjects were of European ancestry and were evaluated by qualified neurologists of the Parkinson Centre of the IRCCS INM Neuromed from June 2015 to December 2017, and from June 2021 to December 2023, with a thorough protocol comprising neurological examination and evaluation of non-motor domains. Information about family history, demographic characteristics, anamnesis, and pharmacological therapy was also collected (the treatment of the PD groups consisted for the most part of a combination of levodopa and dopamine agonists) ^9,11^. Clinical criteria for diagnosis required the presence of at least two cardinal motor signs: asymmetric resting tremor, bradykinesia and rigidity, as well as a good response to levodopa and absence of other atypical features and causes of parkinsonism. Exclusion Criteria for enrolment were: *i*) pre-existing psychiatric conditions; *ii*) presence of neurodegenerative neurological diseases such as multiple sclerosis, lateral sclerosis amyotrophic, Alzheimer’s, neuromuscular pathologies, epilepsy; *iii*) diagnosis of dementia; *iv*) depression; *v*) prolonged intake of anxiolytics, antidepressants, antipsychotics, hypnotic drugs, cognitive stimulants.

The Movement Disorder Society revised version of the Unified Parkinson’s Disease Rating Scale Part III (33 items, maximum score 132; hereafter called UPDRS) ^121^ was used to assess clinical motor symptoms which include: language, facial expressions, tremor, rigidity, agility in movements, stability, gait and bradykinesia. Patients were analyzed during the ON period.

Non-motor symptoms were assessed through an Italian validated version of Non Motor Symptoms Scale (NMS) for Parkinson Disease ^122^. This scale tests 9 items, including cardiovascular domain, sleep/fatigue, mood/cognition, perceptual problems/hallucinations, attention/memory, gastrointestinal, urinary, sexual function, and ability to taste or smell.

The PD cohort includes 48 idiopathic and 119 genetic PD patients carrying at least one pathogenic mutation in the most frequently mutated PD genes such as *LRRK2*, *GBA1*, *TMEM175* and *PARK2/PINK1/PARK7* (see next paragraph “Mutation analysis of the study cohort” for details about the patient selection). The cohort was the same that we used in our recent HPLC and NMR studies^86,112,120^.

#### Ethical Compliance

All procedures involving human participants were approved by the Institutional Review Board of the IRCCS Neuromed Italy. The study protocols N°9/2015, N°19/2020, N°4/2023 have been registered in clinicaltrial.gov with the numbers NCT02403765 (Release Date: 04/01/2015), NCT04620980 (Release Date: 11/03/2020), NCT05721911 (Release Date: 30/01/2023).

Clinical investigations were conducted according to the principles expressed in the Declaration of Helsinki. Written informed consent was obtained from all participants.

The research was carried out following the recommendations set out in the Global Code of Conduct for Research in Resource-Poor Settings.

#### Reporting Standards

“Reporting adheres to the STROBE guidelines for case-control observational studies and MSI guidelines for metabolomics data.”

No formal sample size calculation was performed; the sample size was determined based on the availability of well-characterized subjects from the biobank.

#### MNI-HC cohort

48 neurological healthy controls (HC) were recruited by the same group of neurologists, among the patients’ wives/husbands, after having ascertained the lack of neurological pathologies and the absence of affected family members. Only individuals aged ≥40 years were included. The HC cohort matched for sex and age with PD cohort and was negative for mutation/variant in PD genes.

To minimize potential sources of bias, standardized protocols for clinical assessment, sample collection, and processing were applied. Laboratory analyses were performed on anonymized samples. Sex-matched controls were included to reduce confounding effects.

### Mutation analysis of the Study Cohort

To clarify the impact of the genetic background on the metabolic profile, we enrolled in this study one hundred and sixty-seven PD patients and forty-eight healthy subjects characterized by whole-exome sequencing (WES) (**Table 1 and Table S1**).

In particular, the first group of 119 PD patients, named “genetic-PD”, includes PD-affected subjects carrying of at least 1 pathogenic mutation in one of the most frequently mutated PD genes which include *GBA1*, *LRRK2*, *PARK2*, *PARK7*, *PINK1*, and *TMEM175.* In particular, we selected 17 patients with *LRRK2* mutations, 30 with *GBA1* mutations, 40 with at least 1 mutation in *PARK2/PINK1/PARK7*, and 32 with *TMEM175* mutations (see **Table S2** for the detailed list of mutations/variants).

The second group of 48 PD patients, defined as “idiopathic,” based on WES analysis, which showed no mutations or variants in PD-related genes, as reported in ^9,11^

Similarly, healthy controls (HCs) were selected based on WES analysis confirming the absence of mutations or variants in PD-related genes.

### Collection and storage of serum samples

Blood sampling was performed after a 6-h fasting. Whole blood was collected by peripheral venipuncture into clot activator tubes and gently mixed. Sample was stored upright for 30 min at room temperature to allow blood to clot and centrifuged at 2000 ×g for 10 min at room temperature. Serum was aliquoted (0.5 ml) in polypropylene cryotubes and stored at −80 °C before usage. Unique anonymized codes have been assigned to the samples for processing and subsequent analysis, maintaining the confidentiality of personal data.

### NMR sample preparation

Serum samples were prepared following the established NMR metabolomics standard operating procedures (SOP), as previously reported ^123^. To avoid any manipulation that could change the metabolomic profile, only serum dilution in the acquisition buffer was performed. Subsequently, spectral filtering to remove macromolecular signals was carried out as described in the guidelines ^124^. For NMR tubes preparation, 100 μL of phosphate buffer (0.075 M Na2HPO4·7H2O, 4% NaN3, and water) was combined with 100 μL of blood serum and transferred to a 3 mm NMR tube. Trimethylsilyl propionic acid and sodium salt (0.1% TSP in D_2_O) served as an internal reference for calibrating and measuring NMR signals. NMR analyses were conducted utilizing a Bruker DRX 600 MHz spectrometer (Bruker, Karlsruhe, Germany), outfitted with a 5 mm triple-resonance z-gradient TXI Probe and SampleJet equipped with a rack thermostat. We used TOPSPIN version 3.2 (Bruker Biospin, Fällanden, Switzerland) for spectrometer control and data analysis.

### NMR sample acquisition

Acquiring the Carr-Purcell-Meiboom-Gill (CPMG) spectrum was crucial due to the presence of macromolecules, such as proteins, in serum that could disrupt signals associated with other components and metabolites ^125^.

CPMG experiments were conducted with a spectral width of 7 kHz and 32,000 data points. Water presaturation lasted for 5 seconds during the relaxation delay, and a spin-echo delay of 0.3 ms was utilised. A weighted Fourier transform was applied to the time-domain data with a line broadening of 0.5 Hz, followed by manual phase and baseline adjustments in preparation for targeted profiling analysis. Resonance assignments were conducted utilising Chenomx NMR Suite software, a tool that facilitates spectral deconvolution and signal annotation within an extensive database of reference metabolic profiles. All peaks observed in the ^1^H NMR spectra were meticulously analysed and manually assigned using the Profiler module, which overlays theoretical spectral models onto experimental signals, thereby optimising parameters such as chemical shift, intensity, and peak shape. The manual assignment of peaks and quantification by Chenomx was subsequently verified using the automated software Bayesil. ^123^ In the present study, all amino acid metabolites were annotated in their L-form, as ¹H-NMR spectroscopy does not allow discrimination between D- and L-enantiomers, instead detecting the combined D+L signal. Accordingly, the nomenclature ^126^ reported in the dataset refers to the most abundant enantiomeric species, which, for amino acids in human biofluids, typically corresponds to the L-form.

### Statistical analysis

The primary outcome was serum metabolomic profile. The main exposures were PD subtype (iPD, PD-*GBA1*, PD-*LRRK2*, PD-*PARK2/PINK1/PARK7* and PD-*TMEM175*) and sex. Covariates included age, disease duration, and L-DOPA equivalent daily dose (LEDD).

The biostatistical analysis was conducted on the quantification matrix, which reports the concentrations of the 43 metabolites for the analysed samples.

Firstly, we examined the concentration matrices using a univariate method that combined a T-test and fold-change analysis, as depicted in a comprehensive Robust volcano plot. We set an absolute fold-change threshold at |FC| > 1.51 and a p-value threshold of less than 0.05 ^127^.

The matrices were normalised using the sum and Pareto scaling methods before analysis. Considering the limited number of samples and the potential for overfitting associated with supervised methods, metabolite concentration matrices were initially employed to conduct an unsupervised analysis through Principal Component Analysis (PCA)^128,129^, validated by a Permutational Multivariate Analysis of Variance (PERMANOVA) test. The PERMANOVA test was also utilised to validate models comparing more than two clusters ^130^.

Partial least-squares discriminant analysis (PLS-DA) was performed on the normalised metabolomics data using MetaboAnalyst 6.0 (http://www.metaboanalyst.ca/) ^131^.

The reduced sample size rendered permutation validation infeasible; therefore, the performance of the PLS-DA model was evaluated using the Q2 coefficient (employing a 10-fold internal cross-validation procedure) and the R2 coefficient, which indicate the variance predicted and explained by the model, respectively. ^132^. Both metrics are considered significant if they yield positive values, and the model’s accuracy was also assessed.

Furthermore, classification models with a non-extremely unbalanced class distribution were evaluated using Random Forest by calculating the model’s out-of-bag (OOB) error and the class-related error, and by explicitly reporting the number of samples belonging to the correct class and the number of incorrectly classified samples ^133,134^.

Discriminatory metabolites were arranged and prioritised based on their variable influence on projection (VIP) scores. The VIP scores, which are computed as the weighted sums of the squares of the PLS-DA weights, indicate the importance of each variable and are deemed statistically significant when they surpass 1. To create an intuitive visualisation of metabolomic profiles, we employed heatmaps utilising normalised data, average group concentrations, and Euclidean distance ^135^. We conducted pathway analysis utilizing the Enrichment tool within MetaboAnalyst 6.0 ^131^, which performs metabolite set enrichment analysis (MSEA) on the quantification matrices based on the established global test method. All enrichment analyses were executed by selecting the Small Molecules Pathways Database (SMPDB) ^136^ specific to Homo sapiens as the database ^137^. The selection of SMPDB was motivated by its connection with the Human Metabolome Database, including the Serum Metabolome Database ^138,139^, where metabolites also acquired through Nuclear Magnetic Resonance are available. Furthermore, the analysis anticipated the presence of a background set comprising all detected metabolites in the spectrum, as shown in **Figure S1.** Only pathways with a false discovery rate (FDR) and adjusted p-values below 0.05, along with more than one associated metabolite (hits), were included. Upset graph was performed using Upset R packages^140^.

To formally test whether the effect of gPD diagnosis on serum metabolite levels was modulated by sex, ordinary least squares (OLS) linear models were fitted for each of the 41/43 outcomes (excluding ethanol and methanol) in the full cohort (HC + gPD; n = 166). The model formula was: log(analyte) ∼ group + sex + age + group:sex, where group = 0 (HC) or 1 (gPD) and sex = 0 (female) or 1 (male). All concentrations were natural log-transformed prior to modelling. Inference was based on heteroscedasticity-consistent robust standard errors (HC3). The group × sex interaction coefficient quantifies whether the difference in analyte levels between gPD and HC differs significantly between males and females. P-values for the interaction term were corrected using the Benjamini-Hochberg false discovery rate (FDR) procedure across 41 outcomes.

## Declaration statements

### Availability of data and materials

Metabolomics data have been deposited to the EMBL-EBI MetaboLights database (https://www.ebi.ac.uk/metabolights/) with the identifier MTBLS12753.

### Code Availability

Not applicable

## Acknowledgments

The authors are grateful to all the patients, their caregivers, the Clinical Parkinson’s Disease Center of IRCCS Pozzilli and the PD biobank of the IRCCS Neuromed and of the IGB-CNR for the kind cooperation with this study.

This study was partially funded by Italian Ministry of University and Research (PRIN 2022 - COD. 2022XF7YYL_02 to AU and PRIN 2022 – COD. 2022W3RKLJ to TE). The work of A.U., T.N. and T.E. was supported by NEXTGENERATIONEU (NGEU) and funded by the Ministry of University and Research (MUR), National Recovery and Resilience Plan (NRRP), project MNESYS (PE0000006) – A Multiscale integrated approach to the study of the nervous system in health and disease (DN. 1553 11.10.2022).

The work of T.E. was supported by Next Generation EU - PNRR M6C2 Investimento 2.1 valorizzazione e potenziamento della ricerca biomedica del SSN grant n. PNRR-MAD-2022-12375960 and grant n. PNRR-MCNT2-2023-12377375. The study was partially funded by Ricerca Corrente of Ministry of Health to TE. The study of TE was partially funded by Ministry of Enterprises and Made in Italy (MIMIT) project Neurotechno n. F/180029/01/X43.

Funders had no role in the study design, data collection, data analysis, manuscript preparation, or decision to publish.

## Author contributions

CM: Investigation, Data analysis, Writing – original draft, Writing – review & editing; FC: Data analysis, Writing – original draft, Writing – review & editing; MS: Writing – review & editing; TN: Data analysis, Writing –review & editing; MV: Data analysis, Writing –review & editing; SP: Resources, Writing – review & editing; NM: Resources, Writing – review & editing; MG: Investigation, Writing – review & editing; FE: Writing – review & editing; AMDU: Supervision, Writing – review & editing; TE: Funding acquisition, Project administration, Supervision, Resources, Writing – review & editing; AU: Conceptualization, Funding acquisition, Project administration, Supervision, Writing – review & editing.

## Competing interests

The authors declare no competing financial or non-financial interests.

## Notes

### Competing Interest Statement

The authors have declared no competing interest.

### Author Declarations

All procedures involving human participants were approved by the Institutional Review Board of the IRCCS Neuromed Italy. The study protocols N.9/2015, N.19/2020, N.4/2023 have been registered in clinicaltrial.gov with the numbers NCT02403765 (Release Date: 04/01/2015), NCT04620980 (Release Date: 11/03/2020), NCT05721911 (Release Date: 30/01/2023). Clinical investigations were conducted according to the principles expressed in the Declaration of Helsinki. Written informed consent was obtained from all participants. The research was carried out following the recommendations set out in the Global Code of Conduct for Research in Resource-Poor Settings.

### Summary of Updates

Compared with the previous medRxiv version of the manuscript, the present study introduces several important conceptual and analytical advances. While the earlier version focused on both sex- and genotype-specific metabolic alterations in genetic Parkinson's disease (gPD), the revised analysis demonstrates that serum metabolomic profiles are largely shared across different PD-associated mutations, with no detectable genotype-specific metabolic signatures when genetic subgroups are directly compared. To strengthen this conclusion, we incorporated additional multivariate analyses, including comparisons among GBA1 severity classes, PARK2/PINK1/PARK7 subgroups, and direct comparisons between genetic and idiopathic PD cohorts. Furthermore, we added an integrated linear modeling approach with a group x sex interaction term, formally demonstrating that sex is a major determinant of metabolite dysregulation in gPD. The revised manuscript also includes new validation procedures based on Random Forest classification, expanded unsupervised PCA analyses, and UpSet plot visualizations to identify shared and sex-dependent discriminant metabolites across genetic subgroups. Collectively, these additions shift the primary focus of the study from genotype-specific metabolic differences toward the identification of common metabolic alterations across genetic forms of PD and the prominent influence of biological sex on circulating metabolomic profiles.

## References

1. Bloem, B. R., Okun, M. S. & Klein, C. Parkinson’s disease. The Lancet vol. 397 2284–2303 Preprint at 10.1016/S0140-6736(21)00218-X (2021).

2. Moisan, F. et al. Parkinson disease male-to-female ratios increase with age: French nationwide study and meta-analysis. J. Neurol. Neurosurg. Psychiatry 87, 952–957 (2016).

3. Taylor, K. S. M., Cook, J. A. & Counsell, C. E. Heterogeneity in male to female risk for Parkinson’s disease. Journal of Neurology, Neurosurgery & Psychiatry 78, 905–906 (2007).

4. Nandipati, S. & Litvan, I. Environmental Exposures and Parkinson’s Disease. Int. J. Environ. Res. Public Health 13, 881 (2016).

5. Jurado-Coronel, J. C. et al. Sex differences in Parkinson’s disease: Features on clinical symptoms, treatment outcome, sexual hormones and genetics. Front. Neuroendocrinol. 50, 18–30 (2018).

6. Reeve, A., Simcox, E. & Turnbull, D. Ageing and Parkinson’s disease: Why is advancing age the biggest risk factor? Ageing Res. Rev. 14, 19–30 (2014).

7. Collier, T. J., Kanaan, N. M. & Kordower, J. H. Ageing as a primary risk factor for Parkinson’s disease: evidence from studies of non-human primates. Nat. Rev. Neurosci. 12, 359–366 (2011).

8. Funayama, M., Nishioka, K., Li, Y. & Hattori, N. Molecular genetics of Parkinson’s disease: Contributions and global trends. J. Hum. Genet. 68, 125–130 (2023).

9. Gialluisi, A. et al. Identification of sixteen novel candidate genes for late onset Parkinson’s disease. Mol. Neurodegener. 16, (2021).

10. Palomba, N. P. et al. Common and Rare Variants in TMEM175 Gene Concur to the Pathogenesis of Parkinson’s Disease in Italian Patients. Mol. Neurobiol. 60, 2150–2173 (2023).

11. Carrillo, F. et al. ANKK1, ANKRD50, GRK5, PACSIN1 and VPS8 are novel candidate genes associated with late onset Parkinson’s disease: Definition of a novel predictive protocol based on polygenic model of inheritance. Neurobiol. Dis. 213, 106996 (2025).

12. Ramirez, A. et al. Hereditary parkinsonism with dementia is caused by mutations in ATP13A2, encoding a lysosomal type 5 P-type ATPase. Nat. Genet. 38, 1184–1191 (2006).

13. Strauss, K. M. et al. Loss of function mutations in the gene encoding Omi/HtrA2 in Parkinson’s disease. Hum. Mol. Genet. 14, 2099–2111 (2005).

14. Paisan-Ruiz, C. et al. Characterization of PLA2G6 as a locus for dystonia-parkinsonism. Ann. Neurol. 65, 19–23 (2009).

15. Fonzo, A. Di et al. *FBXO7* mutations cause autosomal recessive, early-onset parkinsonian-pyramidal syndrome. Neurology 72, 240–245 (2009).

16. Chartier-Harlin, M.-C. et al. Translation Initiator EIF4G1 Mutations in Familial Parkinson Disease. The American Journal of Human Genetics 89, 398–406 (2011).

17. Edvardson, S. et al. A Deleterious Mutation in DNAJC6 Encoding the Neuronal-Specific Clathrin-Uncoating Co-Chaperone Auxilin, Is Associated with Juvenile Parkinsonism. PLoS One 7, e36458 (2012).

18. Krebs, C. E. et al. The Sac1 Domain of *<scp>SYNJ</scp> 1* Identified Mutated in a Family with Early-Onset Progressive <scp>P</scp> arkinsonism with Generalized Seizures. Hum. Mutat. 34, 1200–1207 (2013).

19. Quadri, M. et al. Mutation in the *SYNJ1* Gene Associated with Autosomal Recessive, Early-Onset Parkinsonism. Hum. Mutat. 34, 1208–1215 (2013).

20. Bonifati, V. et al. Mutations in the *DJ-1* Gene Associated with Autosomal Recessive Early-Onset Parkinsonism. Science (1979). 299, 256–259 (2003).

21. Rocha, E. M., Keeney, M. T., Di Maio, R., De Miranda, B. R. & Greenamyre, J. T. LRRK2 and idiopathic Parkinson’s disease. Trends Neurosci. 45, 224–236 (2022).

22. Hu, M. et al. Parkinson’s disease-risk protein TMEM175 is a proton-activated proton channel in lysosomes. Cell 185, 2292–2308.e20 (2022).

23. Wu, L. et al. TMEM175: A lysosomal ion channel associated with neurological diseases. Neurobiol. Dis. 185, 106244 (2023).

24. Carrillo, F. et al. Multiomics approach identifies dysregulated lipidomic and proteomic networks in Parkinson’s disease patients mutated in TMEM175. NPJ Parkinsons Dis. 11, 23 (2025).

25. Salles, P. A., Pizarro-Correa, X. & Chaná-Cuevas, P. Genetics of Parkinsońs disease: Recessive forms. Neurology Perspectives 4, 100147 (2024).

26. Heremans, I. P. et al. Parkinson’s disease protein PARK7 prevents metabolite and protein damage caused by a glycolytic metabolite. Proceedings of the National Academy of Sciences 119, (2022).

27. Dolgacheva, L. P., Berezhnov, A. V., Fedotova, E. I., Zinchenko, V. P. & Abramov, A. Y. Role of DJ-1 in the mechanism of pathogenesis of Parkinson’s disease. J. Bioenerg. Biomembr. 51, 175–188 (2019).

28. Sidransky, E. et al. Multicenter Analysis of Glucocerebrosidase Mutations in Parkinson’s Disease. New England Journal of Medicine 361, 1651–1661 (2009).

29. Blauwendraat, C. et al. Genetic modifiers of risk and age at onset in GBA associated Parkinson’s disease and Lewy body dementia. Brain 143, 234–248 (2020).

30. Bogdanov, M. et al. Metabolomic profiling to develop blood biomarkers for Parkinson’s disease. Brain 131, 389–396 (2008).

31. Hassan-Smith, G., Wallace, G. R., Douglas, M. R. & Sinclair, A. J. The role of metabolomics in neurological disease. J. Neuroimmunol. 248, 48–52 (2012).

32. Jiménez-Jiménez, F. J., Alonso-Navarro, H., García-Martín, E. & Agúndez, J. A. G. Cerebrospinal and blood levels of amino acids as potential biomarkers for Parkinson’s disease: review and meta-analysis. Eur. J. Neurol. 27, 2336–2347 (2020).

33. Kumari, S. et al. Identification of potential urine biomarkers in idiopathic parkinson’s disease using NMR. Clinica Chimica Acta 510, 442–449 (2020).

34. LeWitt, P. A., Li, J., Lu, M., Guo, L. & Auinger, P. Metabolomic biomarkers as strong correlates of Parkinson disease progression. Neurology 88, 862–869 (2017).

35. Luo, X., Liu, Y., Balck, A., Klein, C. & Fleming, R. M. T. Identification of metabolites reproducibly associated with Parkinson’s Disease via meta-analysis and computational modelling. NPJ Parkinsons Dis. 10, 126 (2024).

36. Meoni, G. et al. Metabolite and lipoprotein profiles reveal sex-related oxidative stress imbalance in de novo drug-naive Parkinson’s disease patients. NPJ Parkinsons Dis. 8, 14 (2022).

37. Shao, Y. & Le, W. Recent advances and perspectives of metabolomics-based investigations in Parkinson’s disease. Mol. Neurodegener. 14, 3 (2019).

38. Stoessel, D. et al. Promising Metabolite Profiles in the Plasma and CSF of Early Clinical Parkinson’s Disease. Front. Aging Neurosci. 10, (2018).

39. Yakhine-Diop, S. M. S. et al. Metabolic alterations in plasma from patients with familial and idiopathic Parkinson’s disease. Aging 12, 16690–16708 (2020).

40. Carrillo, F. et al. Multiomics approach discloses lipids and metabolites profiles associated to Parkinson’s disease stages and applied therapies. Neurobiol. Dis. 202, 106698 (2024).

41. Gervasoni, J. et al. Independent serum metabolomics approaches identify disrupted glutamic acid and serine metabolism in Parkinson’s disease patients. NPJ Parkinsons Dis. 11, 274 (2025).

42. Lanore, A. et al. Classification of GBA1 variants and their impact on Parkinson’s disease: an in silico score analysis. NPJ Parkinsons Dis. 11, 226 (2025).

43. Huh, Y. E., Usnich, T., Scherzer, C. R., Klein, C. & Chung, S. J. GBA1 Variants and Parkinson’s Disease: Paving the Way for Targeted Therapy. J. Mov. Disord. 16, 261–278 (2023).

44. Vieira, S. R. L. & Schapira, A. H. V. Glucocerebrosidase mutations and Parkinson disease. J. Neural Transm. 129, 1105–1117 (2022).

45. Koros, C. et al. A Global Perspective of GBA1-Related Parkinson’s Disease: A Narrative Review. Genes (Basel). 15, 1605 (2024).

46. Greuel, A. et al. GBA Variants in Parkinson’s Disease: Clinical, Metabolomic, and Multimodal Neuroimaging Phenotypes. Movement Disorders 35, 2201–2210 (2020).

47. Wallings, R., Manzoni, C. & Bandopadhyay, R. Cellular processes associated with LRRK2 function and dysfunction. FEBS J. 282, 2806–2826 (2015).

48. Bakshi, R. et al. Higher urate in *LRRK2* mutation carriers resistant to Parkinson disease. Ann. Neurol. 85, 593–599 (2019).

49. Bougea, A. et al. Serum Uric Acid in LRRK2 Related Parkinson’s Disease: Longitudinal Data from the PPMI Study. J. Parkinsons Dis. 11, 633–640 (2021).

50. Crotty, G. F. et al. Association of caffeine and related analytes with resistance to Parkinson disease among LRRK2 mutation carriers. Neurology 95, (2020).

51. Johansen, K. K. et al. Metabolomic Profiling in LRRK2-Related Parkinson’s Disease. PLoS One 4, e7551 (2009).

52. Dong, C. et al. Plasma Metabolite Signature Classifies Male LRRK2 Parkinson’s Disease Patients. Metabolites 12, 149 (2022).

53. Hauser, D. N., Primiani, C. T. & Cookson, M. R. The Effects of Variants in the Parkin, PINK1, and DJ-1 Genes along with Evidence for their Pathogenicity. Curr. Protein Pept. Sci. 18, 702–714 (2017).

54. Coukos, R. & Krainc, D. Key genes and convergent pathogenic mechanisms in Parkinson disease. Nat. Rev. Neurosci. 25, 393–413 (2024).

55. Okuzumi, A. et al. Metabolomics-based identification of metabolic alterations in PARK2. Ann. Clin. Transl. Neurol. 6, 525–536 (2019).

56. Balck, A. et al. The role of dopaminergic medication, lipid, and endocannabinoid pathway alterations in idiopathic and PRKN/PINK1-mediated Parkinson’s disease – a large-scale targeted metabolomics study. Preprint at 10.1101/2024.03.01.24303613 (2024).

57. Lucchesi, M. et al. Mitochondrial Dysfunction in Genetic and Non-Genetic Parkinson’s Disease. Int. J. Mol. Sci. 26, 4451 (2025).

58. Okarmus, J. et al. Identification of bioactive metabolites in human iPSC-derived dopaminergic neurons with PARK2 mutation: Altered mitochondrial and energy metabolism. Stem Cell Reports 16, 1510–1526 (2021).

59. Cang, C., Aranda, K., Seo, Y., Gasnier, B. & Ren, D. TMEM175 Is an Organelle K+ Channel Regulating Lysosomal Function. Cell 162, 1101–1112 (2015).

60. Schulze, T. et al. Unraveling pH Regulation of TMEM175, an Endolysosomal Cation Channel With a Role in Parkinson’s Disease. J. Cell. Physiol. 240, (2025).

61. Wang, J. et al. What We Know About TMEM175 in Parkinson’s Disease. CNS Neurosci. Ther. 31, (2025).

62. Cannas, F. et al. Parkinson’s Disease Through the Lens of Metabolomics: A Targeted Systematic Review on Human Studies (2019–2024). J. Clin. Med. 14, 6277 (2025).

63. Farmer, K., Smith, C., Hayley, S. & Smith, J. Major Alterations of Phosphatidylcholine and Lysophosphotidylcholine Lipids in the Substantia Nigra Using an Early Stage Model of Parkinson’s Disease. Int. J. Mol. Sci. 16, 18865–18877 (2015).

64. Su, D. et al. Higher levels of plasma phosphatidylcholine (17:0_18:1) raise the risk of developing Parkinson’s disease. Sci. Rep. 15, 28093 (2025).

65. Plotegher, N. et al. Impaired cellular bioenergetics caused by GBA1 depletion sensitizes neurons to calcium overload. Cell Death Differ. 27, 1588–1603 (2020).

66. Clinical Metabolomics. vol. 2855 (Springer US, New York, NY, 2025).

67. Kennedy, A. D. et al. Metabolomics in the clinic: A review of the shared and unique features of untargeted metabolomics for clinical research and clinical testing. Journal of Mass Spectrometry 53, 1143–1154 (2018).

68. Collino, S., Martin, F. J. & Rezzi, S. Clinical metabolomics paves the way towards future healthcare strategies. Br. J. Clin. Pharmacol. 75, 619–629 (2013).

69. Damiani, C., Gaglio, D., Sacco, E., Alberghina, L. & Vanoni, M. Systems metabolomics: from metabolomic snapshots to design principles. Curr. Opin. Biotechnol. 63, 190–199 (2020).

70. Lawton, K. A. et al. Analysis of the Adult Human Plasma Metabolome. Pharmacogenomics 9, 383–397 (2008).

71. Beckonert, O. et al. Metabolic profiling, metabolomic and metabonomic procedures for NMR spectroscopy of urine, plasma, serum and tissue extracts. Nat. Protoc. 2, 2692–2703 (2007).

72. Beuchel, C. et al. Clinical and lifestyle related factors influencing whole blood metabolite levels – A comparative analysis of three large cohorts. Mol. Metab. 29, 76–85 (2019).

73. Qiu, S. et al. Small molecule metabolites: discovery of biomarkers and therapeutic targets. Signal Transduct. Target. Ther. 8, 132 (2023).

74. Blesa, J., Trigo-Damas, I., Quiroga-Varela, A. & Jackson-Lewis, V. R. Oxidative stress and Parkinson’s disease. Front. Neuroanat. 9, (2015).

75. Dai, C. et al. Glucose metabolism impairment in Parkinson’s disease. Brain Res. Bull. 199, 110672 (2023).

76. Fanning, S., Selkoe, D. & Dettmer, U. Parkinson’s disease: proteinopathy or lipidopathy? npj Parkinson’s Disease vol. 6 Preprint at 10.1038/s41531-019-0103-7 (2020).

77. Henrich, M. T., Oertel, W. H., Surmeier, D. J. & Geibl, F. F. Mitochondrial dysfunction in Parkinson’s disease – a key disease hallmark with therapeutic potential. Mol. Neurodegener. 18, 83 (2023).

78. Tansey, M. G. et al. Inflammation and immune dysfunction in Parkinson disease. Nat. Rev. Immunol. 22, 657–673 (2022).

79. Hu, L. et al. Plasma metabolomics profiles indicate sex differences of lipid metabolism in patients with Parkinson’s disease. Sci. Rep. 14, 31262 (2024).

80. Hamid, Z. et al. Gender specific decrease of a set of circulating N-acylphosphatidyl ethanolamines (NAPEs) in the plasma of Parkinson’s disease patients. Metabolomics 15, 74 (2019).

81. Malik, M. et al. Metabolomic breath landscape analysis unravels lipid biomarker candidates in patients with genetic and idiopathic Parkinson’s disease. NPJ Parkinsons Dis. 12, 33 (2026).

82. Yilmaz, A. et al. Metabolic Profiling of CSF from People Suffering from Sporadic and LRRK2 Parkinson’s Disease: A Pilot Study. Cells 9, 2394 (2020).

83. Cotrin, J. C. et al. Plasma and urinary metabolomic signatures differentiate genetic and idiopathic Parkinson’s disease. Brain Res. 1858, 149625 (2025).

84. Bonanno, L., Caporlingua, M., Castellano, J., Quartarone, A. & Ciurleo, R. Using 1H-Magnetic Resonance Spectroscopy to Evaluate the Efficacy of Pharmacological Treatments in Parkinson’s Disease: A Systematic Review. Int. J. Mol. Sci. 26, 9351 (2025).

85. Lei, S. & Powers, R. NMR Metabolomics Analysis of Parkinson&#39;s Disease. Curr. Metabolomics 1, 191–209 (2013).

86. Marino, C. et al. Untargeted 1H NMR-based metabolomics reveal sex-based differences in blood metabolome profiles among patients with Parkinson’s disease, regardless of their idiopathic or genetic subtype. Preprint at 10.1101/2025.10.30.25339159 (2025).

87. Otto, C. et al. Comprehensive analysis of the cerebrospinal fluid and serum metabolome in neurological diseases. J. Neuroinflammation 21, 234 (2024).

88. Pathan, M., Wu, J., Lakso, H.-Å., Forsgren, L. & Öhman, A. Plasma Metabolite Markers of Parkinson’s Disease and Atypical Parkinsonism. Metabolites 11, 860 (2021).

89. Öhman, A. & Forsgren, L. NMR metabonomics of cerebrospinal fluid distinguishes between Parkinson’s disease and controls. Neurosci. Lett. 594, 36–39 (2015).

90. Ahmed, S. S., Santosh, W., Kumar, S. & Christlet, H. T. T. Metabolic profiling of Parkinson’s disease: evidence of biomarker from gene expression analysis and rapid neural network detection. J. Biomed. Sci. 16, 63 (2009).

91. Berezhnoy, G. et al. Application of IVDr NMR spectroscopy to stratify Parkinson’s disease with absolute quantitation of blood serum metabolites and lipoproteins. Sci. Rep. 15, 17738 (2025).

92. Chaudhary, S. et al. In vitro and in vivo NMR based metabolomics in Parkinson’s disease. J. Magn. Reson. Open 10–11, 100050 (2022).

93. Kumari, S. et al. Quantitative metabolomics of saliva using proton NMR spectroscopy in patients with Parkinson’s disease and healthy controls. Neurological Sciences 41, 1201–1210 (2020).

94. Holmes, E., Tsang, T. M. & Tabrizi, S. J. The application of NMR-based metabonomics in neurological disorders. NeuroRX 3, 358–372 (2006).

95. Wright, R. Mitochondrial dysfunction and Parkinson’s disease. Nat. Neurosci. 25, 2–2 (2022).

96. Pereira, S. L., Grossmann, D., Delcambre, S., Hermann, A. & Grünewald, A. Novel insights into Parkin-mediated mitochondrial dysfunction and neuroinflammation in Parkinson’s disease. Curr. Opin. Neurobiol. 80, 102720 (2023).

97. Bao, W. et al. DJ-1/PARK7 in Parkinson’s disease: mechanisms of pathogenesis and therapeutic potential. Neuroscience 587, 1–13 (2025).

98. Zhang, F. et al. PARK7 enhances antioxidative-stress processes of BMSCs via the ERK1/2 pathway. J. Cell. Biochem. 122, 222–234 (2021).

99. Singh, A., Zhi, L. & Zhang, H. LRRK2 and mitochondria: Recent advances and current views. Brain Res. 1702, 96–104 (2019).

100. Wang, R., Hatano, T., Hattori, N. & Cossu, D. Interplay of GBA1 with lysosomal dysfunction and inflammation in Parkinson’s disease. Neural Regen. Res. https://doi.org/10.4103/NRR.NRR-D-25-01082 (2025) doi:10.4103/NRR.NRR-D-25-01082.

101. Stepien, K. M. et al. Secondary Mitochondrial Dysfunction as a Cause of Neurodegenerative Dysfunction in Lysosomal Storage Diseases and an Overview of Potential Therapies. Int. J. Mol. Sci. 23, 10573 (2022).

102. Vance, J. E. Phospholipid Synthesis and Transport in Mammalian Cells. Traffic 16, 1–18 (2015).

103. van der Veen, J. N. et al. The critical role of phosphatidylcholine and phosphatidylethanolamine metabolism in health and disease. Biochimica et Biophysica Acta (BBA) - Biomembranes 1859, 1558–1572 (2017).

104. Li, Z. & Vance, D. E. Thematic Review Series: Glycerolipids. Phosphatidylcholine and choline homeostasis. J. Lipid Res. 49, 1187–1194 (2008).

105. Greene, J. G., Porter, R. H. P., Eller, R. V. & Greenamyre, J. Inhibition of Succinate Dehydrogenase by Malonic Acid Produces an “Excitotoxic” Lesion in Rat Striatum. J. Neurochem. 61, 1151–1154 (1993).

106. López-Lara, I. M. & Soto, M. J. Fatty Acid Synthesis and Regulation. in Biogenesis of Fatty Acids, Lipids and Membranes 1–17 (Springer International Publishing, Cham, 2018). doi:10.1007/978-3-319-43676-0_26-1.

107. Nuzzo, T. et al. Deep Brain Stimulation rescues the homeostasis disruption of circulating D- and L-amino acids level in men with Parkinson’s Disease. Preprint at 10.1101/2025.08.13.668904 (2025).

108. Figura, M. et al. Serum amino acid profile in patients with Parkinson’s disease. PLoS One 13, e0191670 (2018).

109. Tong, Q. et al. Correlations between plasma levels of amino acids and nonmotor symptoms in Parkinson’s disease. J. Neural Transm. 122, 411–417 (2015).

110. Toczylowska, B., Zieminska, E., Michałowska, M., Chalimoniuk, M. & Fiszer, U. Changes in the metabolic profiles of the serum and putamen in Parkinson’s disease patients – In vitro and in vivo NMR spectroscopy studies. Brain Res. 1748, 147118 (2020).

111. Nagesh Babu, G., et al. Serum metabolomics study in a group of Parkinson’s disease patients from northern India. Clinica Chimica Acta 480, 214–219 (2018).

112. Marino, C. et al. Untargeted 1H NMR-based metabolomics and high-pressure liquid chromatography analysis reveal sexual dimorphism in the serum metabolic profiles of Parkinson’s disease patients harbouring rare genetic variants. Preprint at 10.1101/2025.11.20.25340670 (2025).

113. Torrens-Mas, M., Pons, D.-G., Sastre-Serra, J., Oliver, J. & Roca, P. Sexual hormones regulate the redox status and mitochondrial function in the brain. Pathological implications. Redox Biol. 31, 101505 (2020).

114. Demarest, T. G. & McCarthy, M. M. Sex differences in mitochondrial (dys)function: Implications for neuroprotection. J. Bioenerg. Biomembr. 47, 173–188 (2015).

115. Wu, J. et al. NMR analysis of the CSF and plasma metabolome of rigorously matched amyotrophic lateral sclerosis, Parkinson’s disease and control subjects. Metabolomics 12, 101 (2016).

116. Gervasoni, J. et al. Ultra-performance liquid chromatography-mass spectrometry analysis of post-mortem brain tissue reveals specific amino acid profile dysregulation in Parkinson’s Disease and Alzheimer’s Disease patients. Preprint at 10.1101/2025.07.22.666071 (2025).

117. Di Maio, A. et al. Homeostasis of serine enantiomers is disrupted in the post-mortem caudate putamen and cerebrospinal fluid of living Parkinson’s disease patients. Neurobiol. Dis. 184, 106203 (2023).

118. Yang, J. et al. Targeted serum metabolomics reveals alterations in amino acid and neurotransmitter pathways in Parkinson’s disease. Analyst 151, 1639–1649 (2026).

119. Nagana Gowda, G. A. & Raftery, D. NMR-Based Metabolomics. in 19–37 (2021). doi:10.1007/978-3-030-51652-9_2.

120. Yahyavi, I. et al. Differences in sex and genetic status affect the disruption of NMDAR-related amino acid homeostasis in Parkinson’s disease. Preprint at 10.1101/2025.08.08.25333287 (2025).

121. Goetz, C. G. et al. Movement Disorder Society-sponsored revision of the Unified Parkinson’s Disease Rating Scale (MDS-UPDRS): Scale presentation and clinimetric testing results. Movement Disorders 23, 2129–2170 (2008).

122. Chaudhuri, K. R. et al. International multicenter pilot study of the first comprehensive self-completed nonmotor symptoms questionnaire for Parkinson’s disease: The NMSQuest study. Movement Disorders 21, 916–923 (2006).

123. Marino, C. et al. The Metabolomic Profile in Amyotrophic Lateral Sclerosis Changes According to the Progression of the Disease: An Exploratory Study. Metabolites 12, 837 (2022).

124. Emwas, A.-H. et al. Recommendations for sample selection, collection and preparation for NMR-based metabolomics studies of blood. Metabolomics 21, 66 (2025).

125. Ghini, V., Quaglio, D., Luchinat, C. & Turano, P. NMR for sample quality assessment in metabolomics. N. Biotechnol. 52, 25–34 (2019).

126. Han, B., He, J. & Huang, S. Chiral recognition of some D and L-amino acids by microcrystalline cellulose assisted diffusion-ordered NMR spectroscopy. J. Magn. Reson. Open 22, 100189 (2025).

127. Lötsch, J., Kringel, D. & Ultsch, A. Revisiting Fold-Change Calculation: Preference for Median or Geometric Mean over Arithmetic Mean-Based Methods. Biomedicines 12, 1639 (2024).

128. Worley, B. & Powers, R. PCA as a Practical Indicator of OPLS-DA Model Reliability. Curr. Metabolomics 4, 97–103 (2016).

129. Ruiz-Perez, D., Guan, H., Madhivanan, P., Mathee, K. & Narasimhan, G. So you think you can PLS-DA? BMC Bioinformatics 21, 2 (2020).

130. Anderson, M. J. Permutational Multivariate Analysis of Variance (<scp>PERMANOVA</scp>). in Wiley StatsRef: Statistics Reference Online 1–15 (Wiley, 2017). doi:10.1002/9781118445112.stat07841.

131. Pang, Z. et al. MetaboAnalyst 6.0: towards a unified platform for metabolomics data processing, analysis and interpretation. Nucleic Acids Res. 52, W398–W406 (2024).

132. Szymańska, E., Saccenti, E., Smilde, A. K. & Westerhuis, J. A. Double-check: validation of diagnostic statistics for PLS-DA models in metabolomics studies. Metabolomics 8, 3–16 (2012).

133. Faiz, N. et al. Pruning tree forest and re-sampling for class imbalanced problem. Sci. Rep. 16, 8087 (2026).

134. Zhu, T. Analysis on the Applicability of the Random Forest. J. Phys. Conf. Ser. 1607, 012123 (2020).

135. Xia, J., Sinelnikov, I. V., Han, B. & Wishart, D. S. MetaboAnalyst 3.0—making metabolomics more meaningful. Nucleic Acids Res. 43, W251–W257 (2015).

136. Frolkis, A. et al. SMPDB: The Small Molecule Pathway Database. Nucleic Acids Res. 38, D480–D487 (2010).

137. Jewison, T. et al. SMPDB 2.0: Big Improvements to the Small Molecule Pathway Database. Nucleic Acids Res. 42, D478–D484 (2014).

138. Xia, J., Psychogios, N., Young, N. & Wishart, D. S. MetaboAnalyst: a web server for metabolomic data analysis and interpretation. Nucleic Acids Res. 37, W652–W660 (2009).

139. Psychogios, N. et al. The Human Serum Metabolome. PLoS One 6, e16957 (2011).

140. Conway, J. R., Lex, A. & Gehlenborg, N. UpSetR: an R package for the visualization of intersecting sets and their properties. Bioinformatics 33, 2938–2940 (2017).

