## Supplementary data for "Mutations in the *GBA1*, *LRRK2*, *TMEM175*, *PARK2*, *PINK1*, and *PARK7* genes lead to sex-specific serum metabolic changes in patients with Parkinson’s disease"

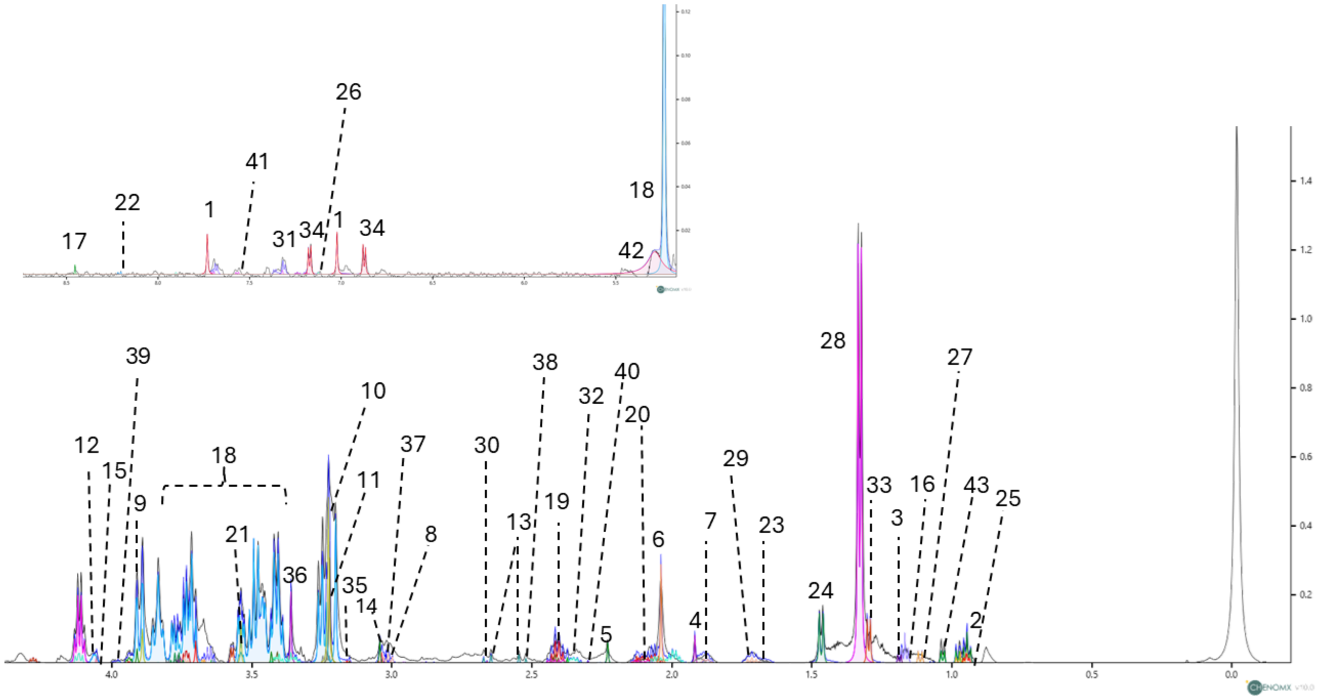

**Figure S1. CPMG spectra related to 1D-$^1$H-CPMG spectrum obtained from the serum of genetic Parkinson's disease patients**. The spectrum is recorded at 600 MHz with a temperature of 298 K. A total of 43 metabolites are identified and labelled as follows: 1:1-Methylhistidine; 2:2-Hydroxybutyric acid; 3:3-Hydroxybutyrate; 4:Acetic acid; 5:Acetoacetate; 6:Acetone; 7:L-Arginine; 8:L-Asparagine; 9:Aspartate; 10: Betaine; 11:Carnitine; 12:Choline; 13:Citric acid; 14:Creatine; 15:Creatinine; 16:Ethanol;17:Formate; 18: D-Glucose; 19: L-Glutamic acid; 20:L-Glutamine; 21:Glycine; 22:Hypoxantine; 23:Isobutyrate; 24:L-Alanine; 25:L-Leucine; 26: L-Histidine; 27:Isoleucine; 28:L-Lactic acid; 29:L-Lysine; 30:L-Methionine; 31:L-Phenylalanine, 32.Proline; 33:L-Threonine; 34:Tyrosine; 35:Malonate;36:Methanol; 37:L-Ornithine; 38:Pyruvic acid; 39: L-Serine, 40:Succinate; 41:L-Tryptophan; 42:Urea; 43:Valine.

a

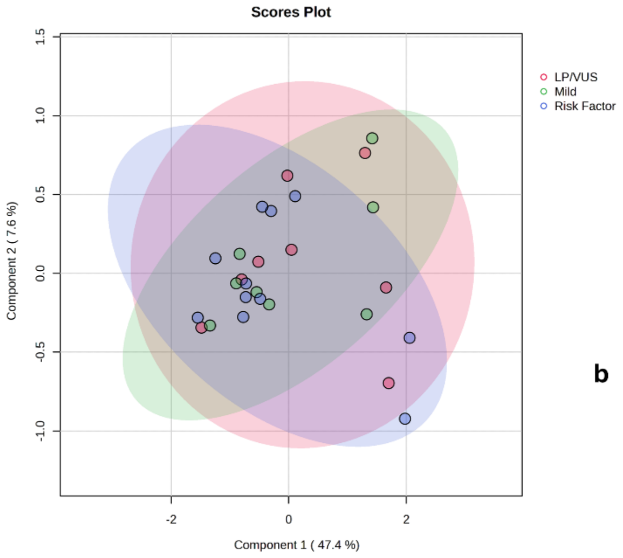

b

**Pair-wise PERMANOVA results**

| | F.Model | R2 | pval | p.adj |
|---|---|---|---|---|
| LP/VUS vs Mild | 0.95265 | 0.06371 | 0.40300 | 0.60450 |
| LP/VUS vs Risk Factor | 1.05390 | 0.05838 | 0.33100 | 0.60450 |
| Mild vs Risk Factor | 0.04093 | 0.00240 | 0.95900 | 0.95900 |

**Figure S2. Partial least squares analysis identified an indistinguishable metabolomic profile among *GBA1*-mutated gPD patients stratified by risk variant status.**

**a.** Scatter plot of the PLS-DA scores (PC1: 47.4%, PC2: 7.6%) was generated for the metabolomic profile of gPD patients to clarify differences between patients with *GBA1* mutations and those with different risk variants (mild (N: 8), risk factor (N: 11), Likely pathogenic/Variants of uncertain significance (LP/VUS) (N: 8)). The PLS-DA approach was validated by 10-fold cross-validation, which reported no separation and non-significant $Q^2$ and $R^2$ indices (for the main component 1: -0.65, 0.64, respectively; for the main component 2: -0.49, 0.79) **b.** The PERMANOVA analysis of the unsupervised principal component analysis model confirmed the absence of clustering (distance F-value: 0.30, p-value: 0.81).

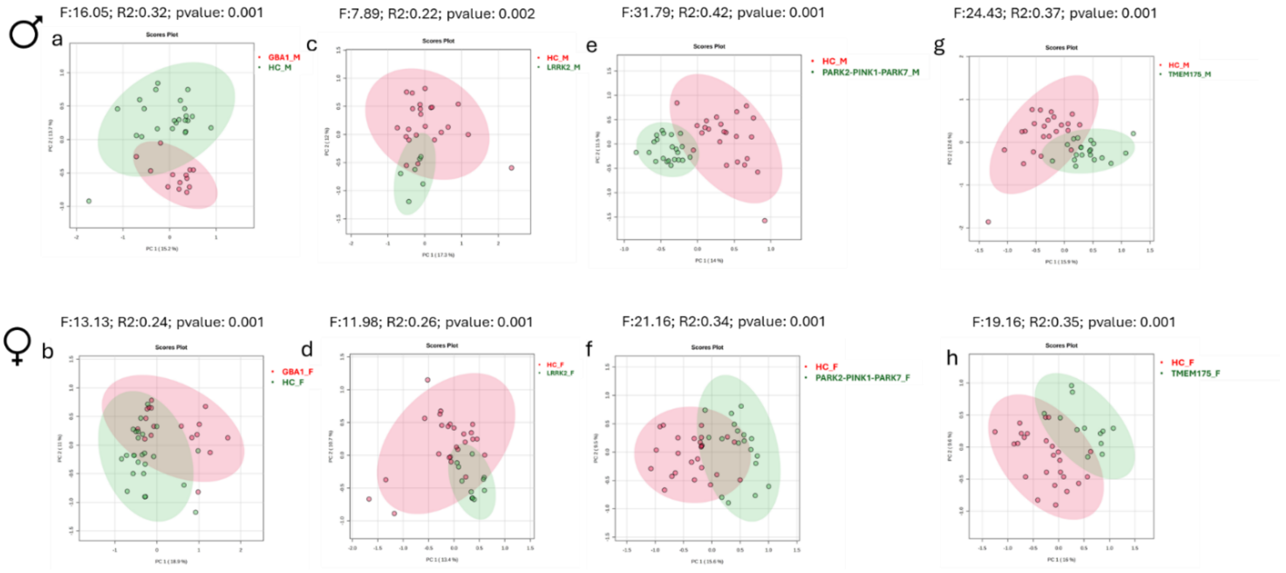

**Figure S3. Principal component analysis of serum metabolomic profiles from gPD patients, stratified by specific gene mutations and compared to healthy controls, validated the absence of overfitting.**

The score plots derived from principal component analysis, validated using PERMANOVA, indicated the absence of overfitting in the PLS-DA multivariate models of the serum metabolomic profile of the gPD cohort, stratified by genotype and gender, compared with sex-matched healthy controls (HC). These plots demonstrate a significant separation of serum metabolomic profiles within the analysed cohort. Specifically, GBA1 (panel a: male gender; PC1: 15.2%, PC2: 13.7%; panel b: female gender; PC1: 18.9%, PC2: 11%), LRRK2 (panel c: male gender; PC1: 17.3%, PC2: 12.0%; panel d: female gender; PC1: 13.4%, PC2: 10.7%), PARK2, PINK1, PARK7 (panel e: male gender; PC1: 14.0%, PC2: 11.5%; panel f: female gender; PC1: 15.6%, PC2: 9.5%), TMEM175 (panel g: male gender; PC1: 15.9%, PC2: 12.9%; panel h: female gender; PC1: 16.6%, PC2: 9.6%). Each model was validated using PERMANOVA, reporting the F value—measuring the ratio of between-group to within-group variability; R2, indicating the variance explained between groups; and p.value, signifying that no random permutation yielded a separation more pronounced than that observed.

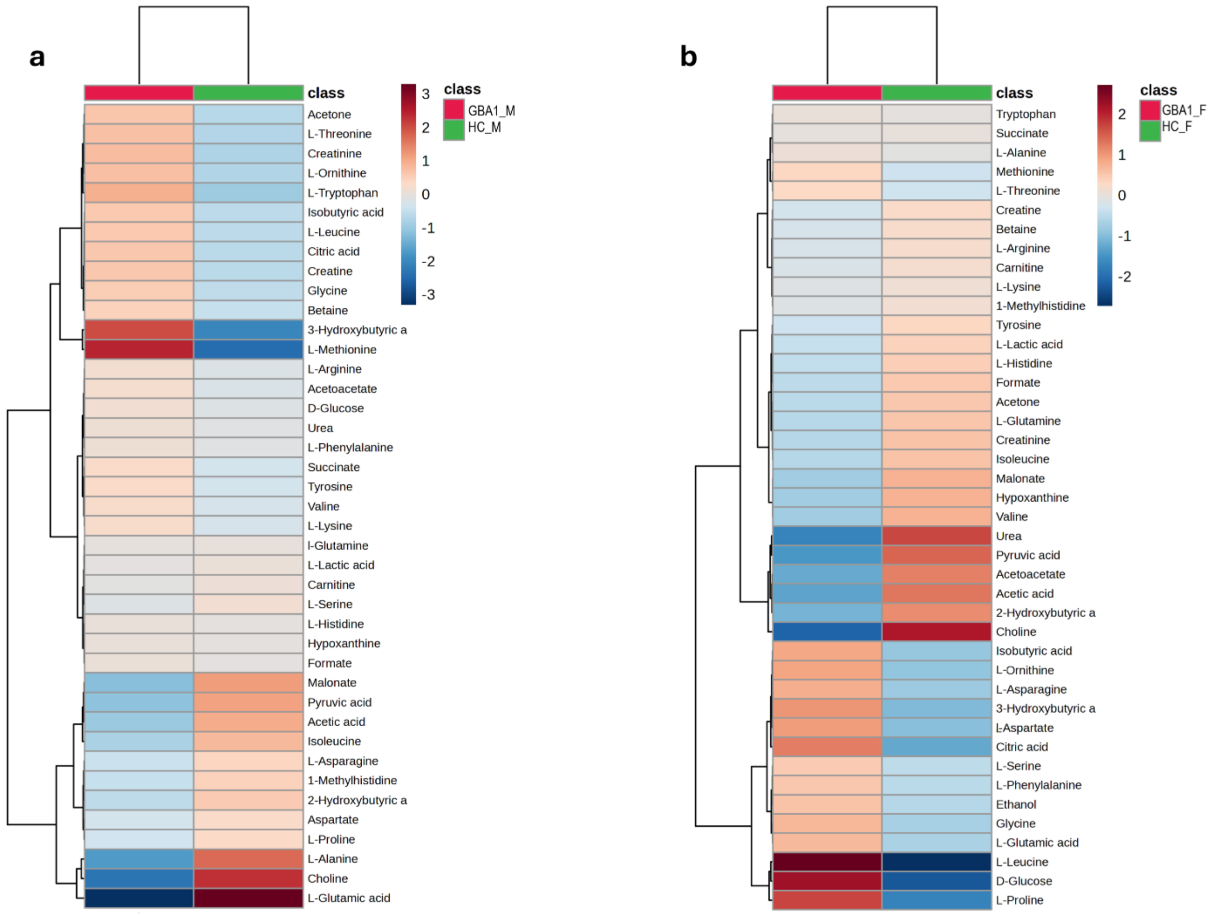

**Figure S4. Average serum metabolomic profile of gPD patients with *GBA1* mutation, stratified by sex.** Heatmap of serum metabolites relative to male **(a)** and female **(b)** gPD patients with *GBA1* mutations (red cluster) compared to HC (green cluster). The colour of each section corresponds to a concentration value of each metabolite calculated from a normalised concentration matrix (red, upregulated; blue, downregulated).

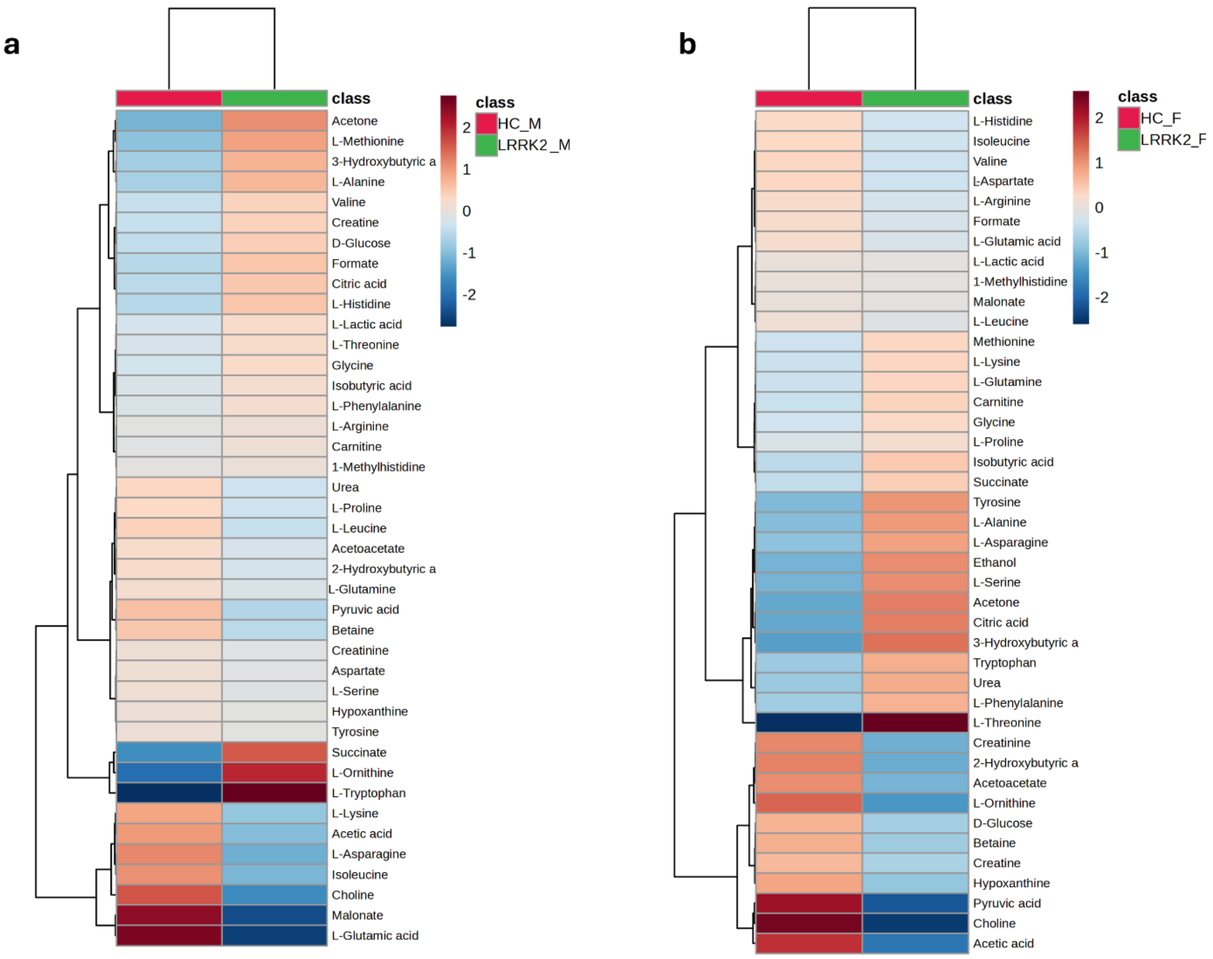

**Figure S5. Average serum metabolomic profile of gPD patients with *LRRK2* mutation, stratified by sex.** Heatmap of serum metabolites relative to male **(a)** and female **(b)** gPD patients with *LRRK2* mutations (green cluster) compared to HC (red cluster). The colour of each section corresponds to a concentration value of each metabolite calculated from a normalised concentration matrix (red, upregulated; blue, downregulated).

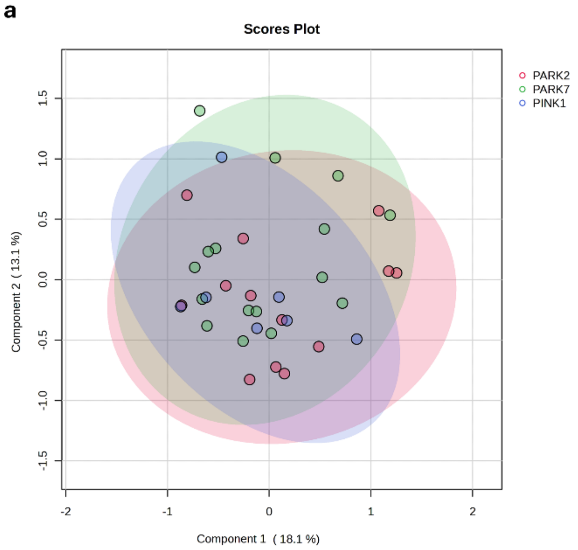

**b** **Pair-wise PERMANOVA results**

| | F.Model | R2 | pval | p.adj |
|---|---|---|---|---|
| PARK2 vs PARK7 | 1.24330 | 0.04402 | 0.29100 | 0.65000 |
| PARK2 vs PINK1 | 0.45141 | 0.02447 | 0.65000 | 0.65000 |
| PARK7 vs PINK1 | 0.60075 | 0.02781 | 0.55600 | 0.65000 |

**Figure S6. PLS-DA analysis identified an indistinguishable metabolomic profile among gPD patients with PARK2, PINK1, and PARK7.**

**a**. Partial Least Squares Discriminant Analysis (PLS-DA) scatter plot (PC: 18.1%, PC2: 13.1%, Principal Component Analysis Q2: -0.50; accuracy: 0.78) was performed on the metabolomic profile of gPD patients to elucidate differences among patients harbouring PARK2 (N:14), PINK1 (N:7), and PARK7 (N:16) mutations. **b**; The PLS-DA model was validated using the PERMANOVA test, which reports the F value, representing the ratio of between-group to within-group variability; R2, indicating the proportion of variance explained between groups; p-value and adjusted p-value (p.adj), demonstrating that no random permutation yielded a stronger separation than the observed. It was necessary to exclude three male subjects—one with a double PINK1–PARK7 mutation and two with PARK2–PARK7 mutations—from the original cohort (male, N=22; female, N=18), as these unique or combined genotypes precluded meaningful statistical comparison.

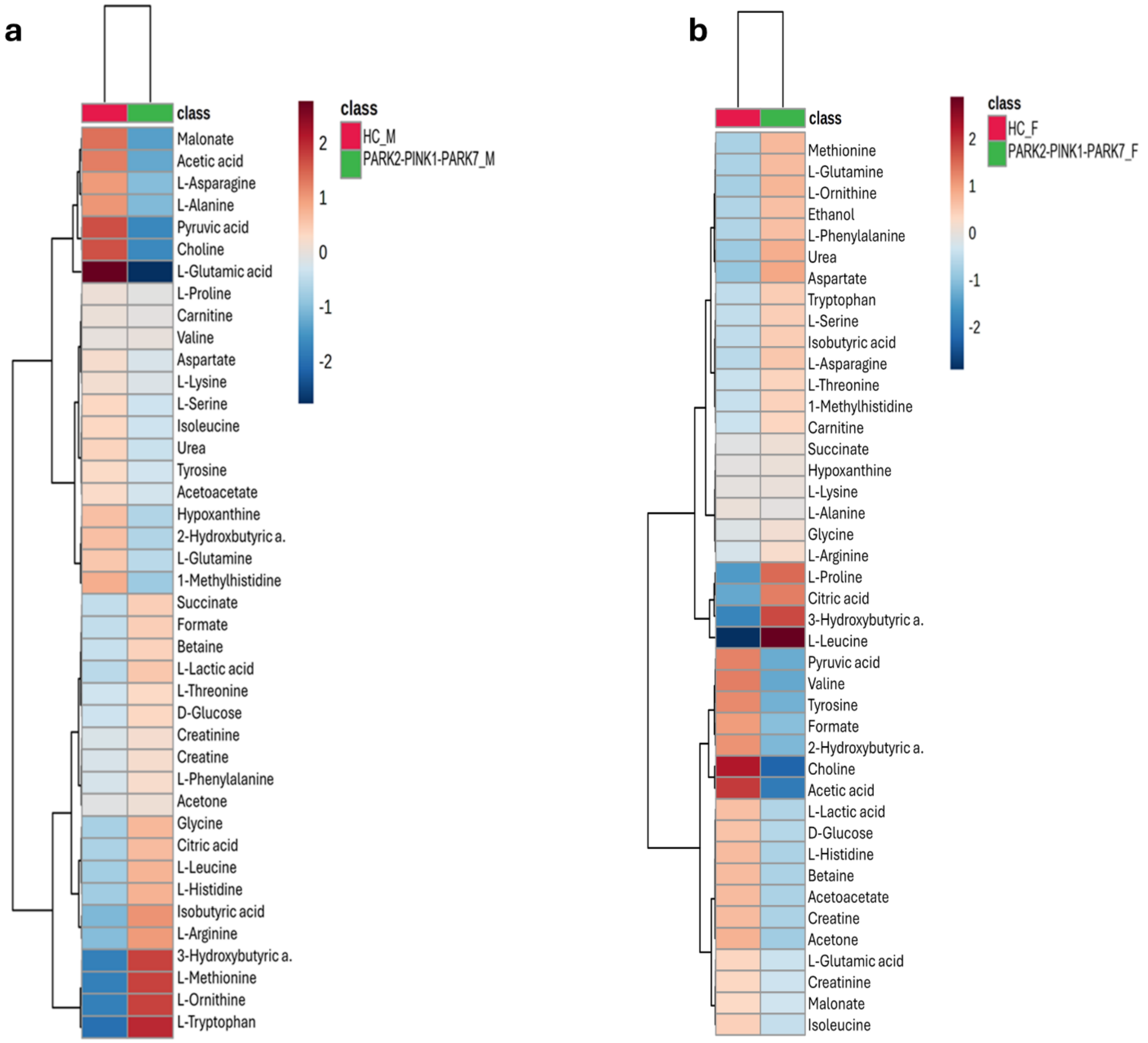

**Figure S7. Average serum metabolomic profile of gPD patients with *PARK2/PINK1/PARK7* mutation, stratified by sex.** Heatmap of serum metabolites relative to male **(a)** and female **(b)** gPD patients with *PARK2/PINK1/PARK7* mutations (green cluster) compared to HC (red cluster). The colour of each section corresponds to a concentration value of each metabolite calculated from a normalised concentration matrix (red, upregulated; blue, downregulated).

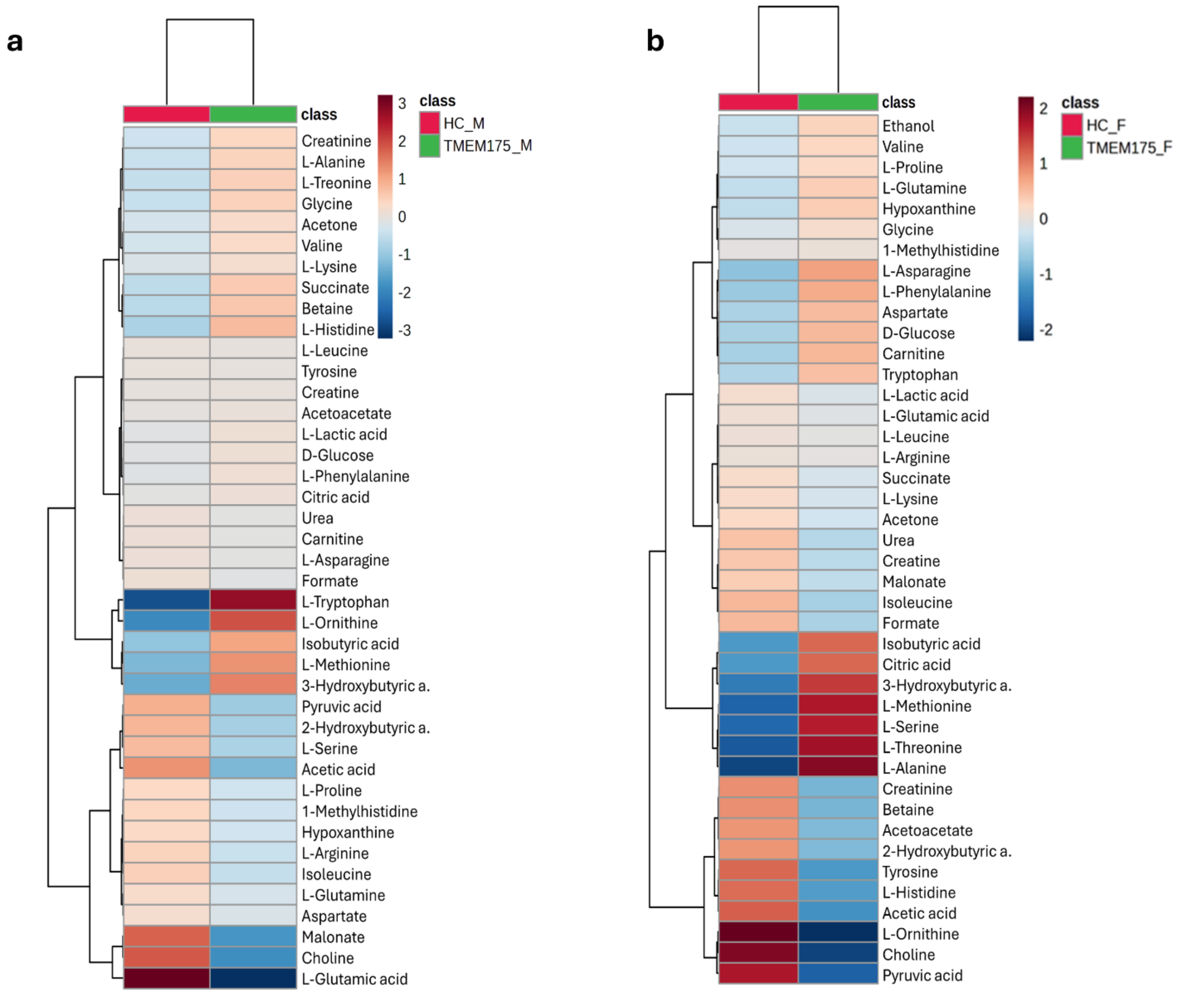

**Figure S8. Average serum metabolomic profile of gPD patients with *TMEM175* mutation, stratified by sex.** Heatmap of serum metabolites relative to male **(a)** and female **(b)** gPD patients with *TMEM175* mutations (green cluster) compared to HC (red cluster). The colour of each section corresponds to a concentration value of each metabolite calculated from a normalised concentration matrix (red, upregulated; blue, downregulated).

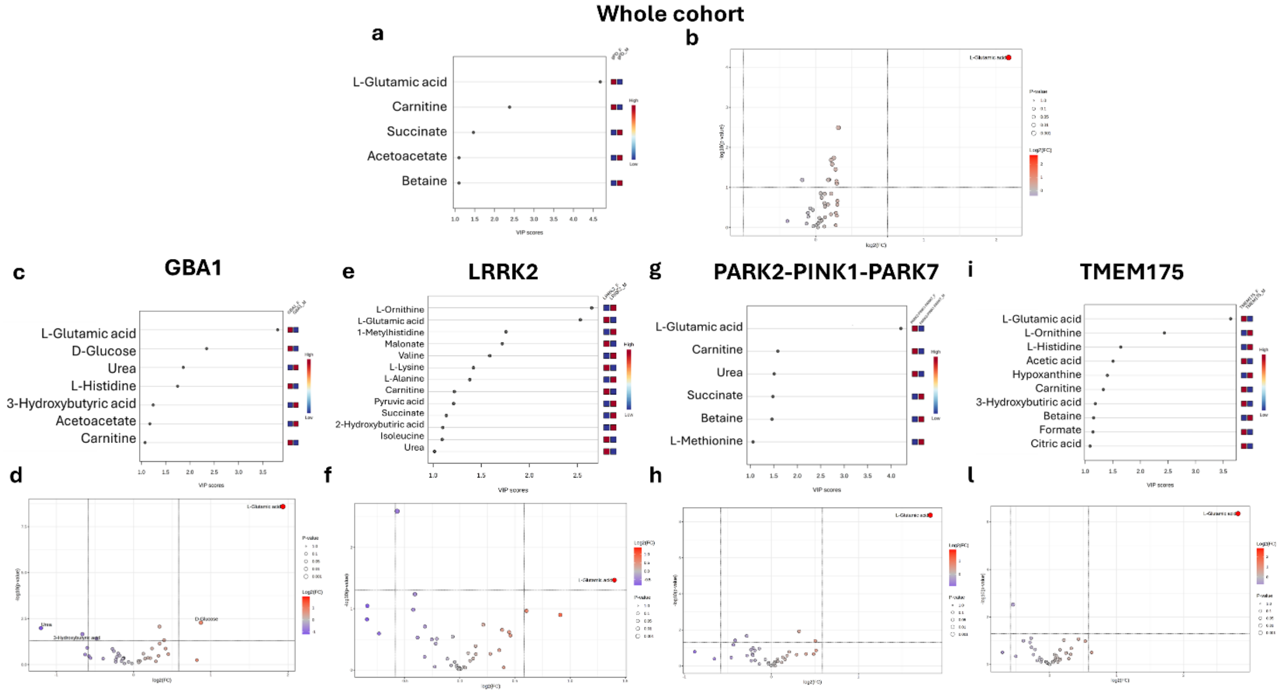

**Figure S9. Sex-dependent metabolic signatures in gPD patients across genetic subgroups.**

**a, c, e, g, i,** Variable Importance in Projection (VIP) analysis identifying metabolites contributing to sex-related differences in the serum metabolome of the entire gPD cohort (a) and of patients stratified by specific pathogenic mutations (GBA1, LRRK2, PARK2–PINK1–PARK7, TMEM175) (c, e, g, i). Only metabolites with VIP > 1 were considered significant. b, d, f, h, l, Robust Volcano plots showing metabolites upregulated (red) or downregulated (blue) in female gPD patients (b) and in female patients within each genetic subgroup (GBA1, LRRK2, PARK2–PINK1–PARK7, TMEM175) (**d, f, h, l**). The absolute fold-change threshold was set to 1.5, and statistical significance was defined as $p < 0.05$

**Table S1.** Comparison of demographic and clinical characteristics between male and female Parkinson's disease patients across different pathogenic mutation groups.

| Pathogenic variants | Demographic and clinical information | Male | | | | Female | | | | p value [a] |
|---|---|---|---|---|---|---|---|---|---|---|
| | | N | Median | IQR | | N | Median | IQR | | |
| ***GBA1*** | Age (years) | 13 | 66.00 | 63.00 | 69.00 | 17 | 69.00 | 65.00 | 74.00 | 0.241 |
| | Age at disease onset (years) | 13 | 61.00 | 57.00 | 66.00 | 17 | 62.00 | 57.00 | 67.00 | 0.900 |
| | Duration of disease (years) | 13 | 5.00 | 2.00 | 8.00 | 17 | 6.00 | 4.00 | 8.00 | 0.256 |
| | LEDD (mg/die) | 13 | 500.00 | 320.00 | 600.00 | 17 | 460.00 | 200.00 | 518.00 | 0.490 |
| | MDS-UPDRS III | 13 | 24.00 | 16.00 | 34.00 | 17 | 16.00 | 10.00 | 23.00 | 0.071 |
| ***LRRK2*** | Age (years) | 6 | 60.50 | 55.00 | 68.00 | 11 | 68.00 | 59.00 | 78.00 | 0.314 |
| | Age at disease onset (years) | 6 | 55.50 | 48.00 | 62.00 | 11 | 58.00 | 50.00 | 67.00 | 0.763 |
| | Duration of disease (years) | 6 | 6.00 | 4.00 | 8.00 | 11 | 11.00 | 8.00 | 17.00 | 0.077 |
| | LEDD (mg/die) | 6 | 525.00 | 286.00 | 700.00 | 11 | 780.00 | 352.00 | 850.00 | 0.269 |
| | MDS-UPDRS III | 6 | 14.50 | 10.00 | 17.00 | 11 | 14.00 | 12.00 | 31.00 | 0.580 |
| ***PARK2-PINK1-PARK7*** | Age (years) | 22 | 69.00 | 61.00 | 75.00 | 18 | 67.50 | 59.00 | 71.00 | 0.226 |
| | Age at disease onset (years) | 22 | 59.50 | 55.00 | 68.00 | 18 | 56.50 | 47.00 | 64.00 | 0.301 |
| | Duration of disease (years) | 22 | 8.00 | 3.00 | 14.00 | 18 | 7.50 | 3.00 | 15.00 | 0.795 |
| | LEDD (mg/die) | 22 | 402.50 | 360.00 | 779.00 | 18 | 439.50 | 336.00 | 620.00 | 0.892 |
| | MDS-UPDRS III | 22 | 24.50 | 14.00 | 29.00 | 18 | 17.50 | 13.00 | 38.00 | 0.644 |
| ***TMEM175*** | Age (years) | 20 | 69.50 | 64.00 | 71.50 | 12 | 67.50 | 66.00 | 74.50 | 0.845 |
| | Age at disease onset (years) | 20 | 61.00 | 57.00 | 65.00 | 12 | 64.00 | 59.00 | 65.00 | 0.638 |
| | Duration of disease (years) | 20 | 6.00 | 2.50 | 9.50 | 12 | 6.50 | 3.00 | 12.00 | 0.740 |
| | LEDD (mg/die) | 20 | 462.50 | 305.00 | 760.00 | 12 | 510.00 | 260.00 | 625.00 | 0.953 |
| | MDS-UPDRS III | 20 | 20.50 | 15.00 | 37.50 | 12 | 22.00 | 16.00 | 34.50 | 0.907 |

Abbreviations: N, number of subjects; IQR, interquartile range, LEDD, Levodopa equivalent daily dose; MDS-UPDRS III, Movement Disorders Society Unified Parkinson's Disease Rating Scale, part III.

[a] Mann-Whitney test

**Table S2. Pathogenic mutations in *LRRK2, GBA1, PARK2, PINK1, PARK7* and *TMEM175* in genetic-PD subtype**

| CHR | Genomic position (hg19) | dbSNP | Gene | RefSEq | Nucleotide Change | AA Change | Exonic Function | Cl | MAF gnomAD v 4.1 | CADD phred | PD patients N |
|---|---|---|---|---|---|---|---|---|---|---|---|
| 12 | 40734202 | rs34637584 | LRRK2 | NM_198578 | c.G6055A | p.G2019S | NSV | P | 0.0002721 | 35 | 14 |
| 12 | 40704236 | rs33939927 | LRRK2 | NM_198578 | c.C4321T | p.R1441C | NSV | P | 0.0000195 | 26.7 | 2 |
| 12 | 40252984 | rs34594498 | LRRK2 | NM_198578 | c.C1256T | p.A419V | NSV | P | 0.0001493 | 24.9 | 1 |
| 1 | 155235843 | rs76763715 | GBA1 | NM_000157 | c.A1226G | p.N409S | NSV | P | 0.001728 | 24.1 | 9 |
| 1 | 155236376 | rs2230288 | GBA1 | NM_000157 | c.G1093A | p.E365K | NSV | P | 0.01338 | 16.1 | 7 |
| 1 | 155236246 | rs75548401 | GBA1 | NM_000157 | c.C1223T | p.T408M | NSV | P | 0.008175 | 21.3 | 3 |
| 1 | 155235252 | rs421016 | GBA1 | NM_000157 | c.T1448C | p.L483P | NSV | P | 0.00007547 | 24.7 | 2 |
| 1 | 155237458 | rs367968666 | GBA1 | NM_000157 | c.T882G | p.H294Q | NSV | P | 0.0001678 | 12.25 | 2 |
| 1 | 155240696 | rs139626710 | GBA1 | NM_000157 | c.A49G | p.R17G | NSV | P | 0.00002119 | 8.03 | 1 |
| 1 | 155239995 | NA | GBA1 | NM_000157 | c.C198A | p.D66E | NSV | P | NA | 6.32 | 1 |
| 1 | 155238206 | rs381427 | GBA1 | NM_000157 | c.T689A | p.V230E | NSV | P | 0.00000339 | 22.3 | 1 |
| 1 | 155238177 | NA | GBA1 | NM_000157 | c.C718T | p.P240S | NSV | P | NA | 22.4 | 1 |
| 1 | 155236370 | NA | GBA1 | NM_000157 | c.C1099T | p.H367Y | NSV | P | NA | 25.7 | 1 |
| 1 | 155236261 | rs121908307 | GBA1 | NM_000157 | c.G1208C | p.S403T | NSV | P | 8.476e-7 | 22.8 | 1 |
| 1 | 155235790 | rs149171124 | GBA1 | NM_000157 | c.G1279A | p.E427K | NSV | P | 0.0002229 | 19.29 | 1 |
| 1 | 155235727 | rs1064651 | GBA1 | NM_000157 | c.G1342C | p.D448H | NSV | P | 0.0001043 | 23.3 | 1 |
| 1 | 155235205 | rs369068553 | GBA1 | NM_000157 | c.G1495C | p.V499L | NSV | P | 0.00004747 | 17.25 | 1 |
| 1 | 155235091 | NA | GBA1 | NM_000157 | c.G1515T | p.K505N | NSV | P | NA | 19.17 | 1 |

| | | | | | | | | | | | |
|---|---|---|---|---|---|---|---|---|---|---|---|
| 6 | 161350139 | rs775091228 | PRKN | NM_004562 | c.G1358A | p.W453X | Stop_gain | P | 0.00001186 | 44 | 2 |
| 6 | 161785820 | rs34424986 | PRKN | NM_004562 | c.C823T | p.R275W | NSV | P | 0.003732 | 26.1 | 2 |
| 6 | 161360169 | rs55830907 | PRKN | NM_004562 | c.C1204T | p.R402C | NSV | P | 0.001937 | 25.1 | 4 |
| 6 | 162262692 | rs55774500 | PRKN | NM_004562 | c.C245A | p.A82E | NSV | P | 0.002815 | 3.14 | 5 |
| 6 | 162054135 | rs9456735 | PRKN | NM_004562 | c.A574C | p.M192L | NSV | P | 0.0001356 | 20.7 | 1 |
| 6 | 161973335 | rs144032774 | PRKN | NM_004562 | c.G701A | p.R234Q | NSV | P | 0.0001280 | 20.9 | 1 |
| 6 | 161973317 | rs137853054 | PRKN | NM_004562 | c.C719T | p.T240M | NSV | P | 0.0001802 | 23.4 | 1 |
| 6 | 161973306 | NA | PRKN | NM_004562 | c.G730C | p.V244L | NSV | P | NA | 6.93 | 1 |
| 6 | 161360148 | NA | PRKN | NM_004562 | c.G1225T | p.E409X | Stop_gain | P | NA | 40 | 1 |
| 6 | 161360129 | rs778125254 | PRKN | NM_004562 | c.C1244A | p.T415N | NSV | P | 0.00000339 | 25.2 | 1 |
| 1 | 20638041 | rs138302371 | PINK1 | NM_032409 | c.C587T | p.P196L | NSV | P | 0.0003610 | 17.46 | 2 |
| 1 | 20649054 | rs74315356 | PINK1 | NM_032409 | c.G1311A | p.W437X | Stop_gain | P | 0.00000677 | 47 | 2 |
| 1 | 20637956 | rs768091663 | PINK1 | NM_032409 | c.G502C | p.A168P | NSV | P | 0.00000847 | 22.2 | 1 |
| 1 | 20638012 | rs143204084 | PINK1 | NM_032409 | c.G558C | p.K186N | NSV | P | 0.0002568 | 14.84 | 1 |
| 1 | 20644585 | NA | PINK1 | NM_032409 | c.C872A | p.A291D | NSV | P | NA | 26.3 | 1 |
| 1 | 20645576 | rs376323248 | PINK1 | NM_032409 | c.C976T | p.R326C | NSV | P | 0.00000508 | 26.9 | 1 |
| 1 | 20650518 | rs531477772 | PINK1 | NM_032409 | c.G1573A | p.D525N | NSV | P | 0.00009152 | 23.3 | 1 |
| 1 | 7970934 | rs71653619 | PARK7 | NM_007262 | c.G293A | p.R98Q | NSV | RF | 0.01043 | 20.9 | 14 |
| 1 | 7969406 | rs781094807 | PARK7 | NM_007262 | c.252+2->A | | splicing | N | 0.0002733 | NA | 2 |
| 4 | 952418 | rs752406714 | TMEM175 | NM_032326 | c.430_431del | p.V147Dfs*104 | Fs_del | P | 0.00000256 | NA | 4 |
| 4 | 958262 | rs565504915 | TMEM175 | NM_032326 | c.1281_1282del | p.A429Qfs*120 | Fs_del | P | 0.0003095 | NA | 3 |

| | | | | | | | | | | | |
|---|---|---|---|---|---|---|---|---|---|---|---|
| 4 | 952434 | NA | TMEM175 | NM_032326 | c.446_462del | p.A149Gfs*97 | Fs_del | P | NA | NA | 3 |
| 4 | 947842 | rs542936413 | TMEM175 | NM_032326 | c.C103T | p.R35C | NSV | P | 0.00002542 | 28.7 | 3 |
| 4 | 958194 | rs75307864 | TMEM175 | NM_032326 | c.C1213G | p.L405V | NSV | P | 0.003832 | 23.3 | 4 |
| 4 | 958221 | rs140597786 | TMEM175 | NM_032326 | c.C1240T | p.R414W | NSV | P | 0.004833 | 23.2 | 3 |
| 4 | 957985 | rs147762522 | TMEM175 | NM_032326 | c.G1004A | p.R335H | NSV | P | 0.001076 | 24.6 | 3 |
| 4 | 950461 | rs142778595 | TMEM175 | NM_032326 | c.T233C | p.I78T | NSV | P | 0.00005424 | 23.4 | 2 |
| 4 | 951229 | rs200834686 | TMEM175 | NM_032326 | c.A313G | p.T105A | NSV | P | 0.0001136 | 22.21 | 2 |
| 4 | 955826 | rs746980739 | TMEM175 | NM_032326 | c.C778T | p.R260C | NSV | P | 0.00004068 | 25.1 | 1 |
| 4 | 955856 | rs750645874 | TMEM175 | NM_032326 | c.G808A | p.A270T | NSV | P | 0.00000762 | 25.0 | 1 |
| 4 | 955888 | NA | TMEM175 | NM_032326 | c.C840G | p.I280M | NSV | P | NA | 24.7 | 1 |
| 4 | 957838 | rs778399444 | TMEM175 | NM_032326 | c.C857T | p.P286L | NSV | P | 0.00000254 | 23.9 | 1 |
| 4 | 957958 | rs148627215 | TMEM175 | NM_032326 | c.C977T | p.A326V | NSV | P | 0.00000169 | 7.41 | 1 |
| 4 | 958024 | rs147975675 | TMEM175 | NM_032326 | c.C1043T | p.S348L | NSV | P | 0.00005171 | 24.5 | 1 |
| 4 | 958252 | rs142744759 | TMEM175 | NM_032326 | c.C1271T | p.A424V | NSV | P | 0.00001188 | 22.6 | 1 |
| 4 | 958422 | rs201314478 | TMEM175 | NM_032326 | c.C1441T | p.R481W | NSV | P | 0.002678 | 10.6 | 1 |

CHR, Chromosome, hg19, human genome build to which these variants are annotated, dbSNP, reference number in SNP database, ref seq, reference number of the gene transcript, AA Change, amino acid change, CI, clinical interpretation, P, Pathogenic, RF, risk factor, N, novel, PD, Parkinson's disease, CADD phred, Combined Annotation Dependent Depletion, Fs_del, frameshift deletion, NSV, non-synonymous variant, MAF, Minor Allele Frequency, was referred to gnomAD v4.1 database, NA, Not Annotated.

**Table S3. OOB error calculated using a random forest approach for the classification of clustering patterns among metabolomic profiles of gPDs affected by different mutations, stratified by gender.**

| | GBA1_M | HC_M | Class.error | OOB error |
|---|---|---|---|---|
| GBA1_M | 13 | 0 | 0.0 | 0.0 |
| HC_M | 0 | 23 | 0.0 | |
| | GBA1_F | HC_F | Class.error | OOB error |
| GBA1_F | 15 | 2 | 0.11 | 0.0476 |
| HC_F | 0 | 25 | 0.0 | |
| | PARK2-PINK1-PARK7_M | HC_M | Class.error | OOB error |
| PARK2-PINK1-PARK7_M | 22 | 0 | 0.0 | 0.0 |
| HC_M | 0 | 23 | 0.0 | |
| | PARK2-PINK1-PARK7_F | HC_F | Class.error | OOB error |
| PARK2-PINK1-PARK7_F | 16 | 2 | 0.11 | 0.0465 |
| HC_F | 0 | 25 | 0 | |
| | TMEM175_M | HC_M | Class.error | OOB error |
| TMEM175_M | 19 | 1 | 0.05 | 0.0233 |
| HC_M | 0 | 23 | 0.0 | |
| | TMEM175_F | HC_F | Class.error | OOB error |
| TMEM175_F | 11 | 1 | 0.08 | 0.0270 |
| HC_F | 0 | 25 | 0.0 | |

The table summarises the performance of Random Forest classification models applied to different genetic groups (*GBA1, PARK2-PINK1-PARK7, TMEM175*), stratified by sex and compared with their respective controls. For each model, the class error (class.error) and the out-of-bag error (OOB error) are reported, with the latter serving as an internal estimate of the model's accuracy. Random Forest models showed very low OOB errors (0.0–0.0476) across all comparisons, with class errors observed only in the mutated groups (0.05–0.11) and absent in the controls. Due to the small size of the *LRRK2* group, classification models were not estimated for this comparison to avoid unstable and potentially misleading results.

**Table S4. Robust Volcano plot results related to Parkinson's disease patients with *GBA1* mutation compared to age- and sex-matched healthy controls.**

| Metabolites *GBA1*_M | FC | \|FC\| | log2(FC) | p.value |
|---|---|---|---|---|
| L-Glutamic acid | 0.45 | 2.22 | -1.15 | 1.26*E-08 |
| Choline | 0.50 | 2.00 | -1.00 | 0.006 |
| L-Methionine | 1.81 | 1.81 | 0.86 | 0.004 |
| 3-Hydroxybutyric acid | 1.70 | 1.70 | 0.77 | 0.01 |
| Pyruvic acid | 0.53 | 1.89 | -0.92 | 0.03 |
| **Metabolites *GBA1*_F** | **FC** | **\|FC\|** | **log2(FC)** | **p.value** |
| Choline | 0.59 | 1.69 | -0.76 | 5.62*E-07 |
| Pyruvic acid | 0.54 | 1.85 | -0.89 | 0.003 |
| L-Leucine | 1.83 | 1.83 | 0.87 | 1.0*E-09 |
| L-Proline | 1.74 | 1.74 | 0.80 | 3.98*E-05 |
| D-Glucose | 2.14 | 2.14 | 1.10 | 0.008 |

Results from univariate analysis were presented using a Robust Volcano plot of metabolite concentrations detected in the 1d-CPMG NMR spectrum. The analysis combines the Fold change (FC) test, calculated as the ratio of average concentrations between the GBA1_M/HC_M and GBA1_F/HC_F clusters—using a threshold of absolute fold change |FC| > 1.5—and the p-value from the T-Test, which is considered significant if less than 0.05

**Table S5. Pathway enrichment analysis of male *GBA1* carriers**

| | Hits | Raw p | Holm p | FDR | Metabolites |
|---|---|---|---|---|---|
| Glycine and Serine Metabolism | 11 | 1.27E-08 | 8.91E-07 | 5.91E-07 | Betaine, Creatine, Glycine, L-Glutamic acid, L-Alanine, L-Threonine, L-Serine, L-Ornithine, Pyruvic acid, L-Methionine, L-Arginine. |
| Amino Sugar Metabolism | 4 | 2.84E-07 | 1.87E-05 | 3.98E-06 | Acetic acid, L-Glutamic acid, Pyruvic acid, L-Glutamine. |
| Alanine Metabolism | 4 | 6.92E-07 | 4.43E-06 | 6.92E-06 | Glycine, L-Glutamic acid, L-Alanine, Pyruvic acid. |
| Glucose-Alanine Cycle | 4 | 1.03E-06 | 6.07E-04 | 6.00E-05 | D-Glucose, L-Glutamic acid, L-Alanine, Pyruvic acid. |
| Glutamate Metabolism | 7 | 2.49E-06 | 1.57E-04 | 2.18E-05 | L-Glutamic acid, Pyruvic acid, Aspartate, Glycine, L- Alanine, L-Glutamine, Succinic acid. |
| Malate-Aspartate Shuttle | 2 | 3.00E-06 | 1.86E-04 | 2.34E-05 | Aspartate, L-Glutamic acid. |
| Histidine Metabolism | 3 | 4.13E-06 | 2.52E-04 | 2.89E-05 | Glutamic acid, L-Histidine, 1-Methylhistidine. |
| Glutathione Metabolism | 3 | 5.25E-06 | 3.15E-04 | 3.34E-05 | Glycine, L-Glutamic acid, L-Ornithine. |
| Tryptophan Metabolism | 4 | 1.10E-05 | 5.84E-03 | 4.29E-04 | L-Glutamic acid, L-Tryptophan, Formic acid, L-Alanine. |
| Propanoate Metabolism | 3 | 1.31E-05 | 7.58E-04 | 6.63E-06 | 2-Hydroxybutyric acid, L-Glutamic acid, L-Valine. |
| Purine Metabolism | 5 | 1.33E-05 | 7.58E-04 | 6.63E-06 | L-Glutamic acid, Hypoxanthine, L-Glutamine, Aspartate, Glycine. |
| Aspartate Metabolism | 6 | 4.04E-05 | 2.26E-03 | 1.89E-04 | L-Glutamic acid, Acetic acid, L-Asparagine, Aspartate, L-Glutamine, L-Arginine. |
| Urea Cycle | 8 | 1.05E-04 | 5.76E-03 | 4.29E-04 | L-Glutamic acid, L-Alanine, Aspartate, Ornithine, Pyruvic acid, Urea, L-Arginine, L-Glutamine. |
| Beta-Alanine Metabolism | 3 | 1.05E-04 | 5.76E-03 | 4.29E-04 | L-Glutamic acid, Aspartate, L-Histidine. |
| Ammonia Recycling | 8 | 4.40E-04 | 2.29E-02 | 1.62E-03 | L-Glutamic acid, Pyruvic acid, L-Asparagine, Glycine, Serine, Aspartate, L-Histidine, L-Glutamine. |

Pathway enrichment analysis performed on $^{1}$H-NMR and related to male and female gPD with GBA1 mutations and HC serum metabolomic results. Hits represent the numbers of metabolites involved in the pathways and specified in the 'Metabolites' column. The pathways were considered statistically significant with Hits >1, p-value < 0.05, adjusted p-value performed using Holm Bonferroni test (Holm p) and False Discovery Rate (FDR) < 1.

### **Table S6. Pathway enrichment analysis of female *GBA1* carriers**

| | Hits | Raw p | Holm p | FDR | Metabolites |
|---|---|---|---|---|---|
| Valine Leucine and Isoleucine Degradation | 6 | 5.83E-08 | 4.08E-04 | 4.08E-02 | Acetoacetic acid, L-Glutamic acid, Isoleucine, Succinic acid, L-Leucine, Valine. |
| Phosphatidylethanolamine Biosynthesis | 2 | 1.02E-06 | 6.84E-04 | 1.79E-02 | Choline, L-Serine. |
| Methionine Metabolism | 5 | 1.30E-05 | 8.60E-04 | 1.80E-04 | Betaine¸Glycine, L-Serine, L-Methionine, Choline. |
| Betaine Metabolism | 3 | 1.66E-05 | 1.10E-03 | 1.90E-04 | Betaine, L-Methionine, Choline. |
| Amino Sugar Metabolism | 4 | 5.40E-04 | 3.40E-02 | 5.40E-03 | Acetic acid, L-Glutamic acid, Pyruvic acid, L- Glutamine. |

Pathway enrichment analysis performed on $^1$H-NMR and related to male and female gPD with GBA1 mutations and HC serum metabolomic results. Hits represent the numbers of metabolites involved in the pathways and specified in the 'Metabolites' column. The pathways were considered statistically significant with Hits >1, p-value < 0.05, adjusted p-value performed using Holm Bonferroni test (Holm p) and False Discovery Rate (FDR) < 1.

**Table S7. Robust Volcano plot results related to Parkinson's disease patients with *LRRK2* mutation compared to age- and sex-matched healthy controls.**

| **Metabolites *LRRK2*_M** | **FC** | **\|FC\|** | **log2(FC)** | **p.value** |
|---|---|---|---|---|
| L-Tryptophan | 2.19 | 2.19 | 1.13 | 1.00E-10 |
| L-Glutamic acid | 0.43 | 2.33 | -1.22 | 7.49E-09 |
| Malonate | 0.47 | 2.13 | -1.09 | 0.0002 |
| L-Ornithine | 1.58 | 1.58 | 0.66 | 0.0003 |
| Choline | 0.55 | 1.82 | -0.86 | 0.004 |
| Succinate | 1.54 | 1.54 | 0.62 | 0.02 |
| **Metabolites *LRRK2*_F** | **FC** | **\|FC\|** | **log2(FC)** | **p.value** |
| Choline | 0.61 | 1.64 | -0.71 | 5.62E-06 |
| L-Threonine | 1.54 | 1.54 | 0.62 | 2.51E-05 |
| Pyruvic acid | 0.50 | 2.00 | -1.00 | 0.0003 |
| Acetic acid | 0.42 | 2.38 | -1.25 | 0.001 |
| 3-Hydroxybutyric acid | 2 | 2.00 | 1.00 | 0.03 |

Results from univariate analysis were presented using a Robust Volcano plot of metabolite concentrations detected in the 1d-CPMG NMR spectrum. The analysis combines the Fold change (FC) test, calculated as the ratio of average concentrations between the LRRK2_M/HC_M and LRRK2_F/HC_F clusters—using a threshold of absolute fold change |FC| > 1.5—and the p-value from the T-Test, which is considered significant if less than 0.05

**Table S8. Pathways enrichment analysis of male *LRRK2* carriers**

| | Hits | Raw p | Holm p | FDR | Metabolites |
|---|---|---|---|---|---|
| Cysteine Metabolism | 2 | 4.52E-09 | 3.16E-06 | 1.95E-03 | L-Glutamic acid, Pyruvic acid. |
| Tryptophan Metabolism | 4 | 1.04E-08 | 7.09E-05 | 1.95E-03 | L-Glutamic acid, Tryptophan, Formic acid, L-Alanine. |
| Nicotinate and Nicotinamide Metabolism | 2 | 1.11E-07 | 7.45E-05 | 1.95E-03 | L-Glutamic acid, L-Glutamine |
| Amino Sugar Metabolism | 4 | 7.36E-06 | 4.86E-04 | 1.03E-03 | L-Glutamic acid, Pyruvic acid, Acetoacetic acid, L-Glutamine. |
| Lysine Degradation | 2 | 1.22E-05 | 7.95E-04 | 1.43E-03 | L-Glutamic acid, Lysine. |
| Malate-Aspartate Shuttle | 2 | 2.04E-05 | 1.30E-03 | 2.04E-03 | Aspartate, L-Glutamic acid. |
| Urea Cycle | 8 | 2.29E-05 | 1.40E-03 | 2.00E-04 | L-Glutamic acid, L-Alanine, Aspartate, Ornithine, Pyruvic acid, Urea, L-Arginine, L-Glutamine. |
| Alanine Metabolism | 4 | 3.20E-04 | 2.00E-03 | 2.40E-04 | Glycine, L-Glutamic acid, L-Alanine, Pyruvic acid. |
| Glutathione Metabolism | 3 | 4.27E-04 | 2.60E-03 | 2.60E-04 | Glycine, L-Glutamic acid, L-Ornithine. |
| Arginine and Proline Metabolism | 9 | 4.42E-04 | 2.60E-03 | 2.60E-04 | L-Glutamic acid, Proline, Aspartate Succinic acid, Urea, L-Arginine, L-Ornithine, Creatine, Glycine. |
| Histidine Metabolism | 3 | 7.45E-04 | 4.30E-03 | 4.00E-04 | 1-Methylhistidine, L-Glutamic acid, L-Histidine. |
| Propanoate Metabolism | 3 | 8.69E-04 | 4.90E-03 | 4.30E-04 | 2-Hydroxybutyric acid, L-Glutamic acid, Valine. |
| Glutamate Metabolism | 7 | 1.44E-03 | 8.00E-03 | 6.70E-04 | L-Glutamic acid, Pyruvic acid, Aspartate, Glycine, L -Alanine, L-Glutamine, Succinic acid. |
| Aspartate Metabolism | 6 | 1.67E-03 | 9.10E-03 | 7.30E-04 | Acetic acid, L-Glutamic acid, L-Asparagine, Aspartate, L-Arginine, L-Glutamine. |
| Beta-Alanine Metabolism | 3 | 1.99E-03 | 1.00E-02 | 8.10E-04 | L-Glutamic acid, L-Histidine, Aspartate. |
| Glucose-Alanine Cycle | 4 | 2.36E-03 | 1.20E-02 | 9.10E-04 | D-Glucose, L-Glutamic acid, L-Alanine, Pyruvic acid. |

Pathway enrichment analysis performed on $^1$H-NMR and related to male and female gPD with LRRK2 mutations and HC serum metabolomic results. Hits represent the numbers of metabolites involved in the pathways and specified in the 'Metabolites' column. The pathways were considered statistically significant with Hits >1, p-value < 0.05, adjusted p-value performed using Holm Bonferroni test (Holm p) and False Discovery Rate (FDR) < 1.

### Table S9. Pathways enrichment analysis of female *LRRK2* carriers

| | Hits | Raw p | Holm p | FDR | Metabolites |
|---|---|---|---|---|---|
| Amino Sugar Metabolism | 4 | 7.40E-06 | 5.03E-04 | 1.34E-04 | L-Glutamic acid, Pyruvic acid, Acetoacetic acid, L-Glutamine. |
| Glycine and Serine Metabolism | 11 | 8.71E-06 | 5.83E-04 | 1.34E-04 | Betaine, Creatine, Glycine, L-Glutamic acid, L-Alanine, L-Threonine, L-Serine, L-Ornithine, Pyruvic acid, L-Methionine, L-Arginine. |
| Phosphatidylethanolamine Biosynthesis | 2 | 1.41E-05 | 9.16E-04 | 1.64E-04 | Choline, L-Serine. |
| Betaine Metabolism | 3 | 5.38E-05 | 3.39E-03 | 4.71E-04 | Betaine, L-Methionine, Choline. |
| Pyruvate Metabolism | 3 | 6.57E-05 | 6.32E-04 | 1.34E-04 | Acetic acid, Lactic acid, Pyruvic acid. |
| Methionine Metabolism | 5 | 7.09E-05 | 4.40E-03 | 5.52E-04 | Betaine, Glycine, L-Serine, L-Methionine, Choline. |
| Fatty Acid Biosynthesis | 3 | 1.29E-04 | 7.90E-03 | 9.06E-04 | Acetic acid, Acetoacetic acid,3-Hydroxybutyric acid. |
| Transfer of Acetyl Groups into Mitochondria | 3 | 4.40E-04 | 2.60E-02 | 2.57E-03 | Citric acid, D-Glucose, Pyruvic acid. |
| Cysteine Metabolism | 2 | 7.46E-04 | 4.33E-02 | 4.02E-03 | L-Glutamic acid, Pyruvic acid. |

Pathway enrichment analysis performed on $^{1}$H-NMR and related to male and female gPD with LRRK2 mutations and HC serum metabolomic results. Hits represent the number of metabolites involved in the pathways and specified in the 'Metabolites' column. The pathways were considered statistically significant with Hits >1, p-value < 0.05, adjusted p-value performed using Holm Bonferroni test (Holm p) and False Discovery Rate (FDR) < 1.

**Table S10. Robust Volcano plot results related to Parkinson's disease patients with *PARK2-PINK1-PARK7* mutation compared to age- and sex-matched healthy controls.**

| ***Metabolites PARK2-PINK1-PARK7_M*** | **FC** | **\|FC\|** | **log2(FC)** | **p.value** |
|---|---|---|---|---|
| L-Glutamic acid | 0.40 | 2.50 | -1.32 | 3.16E-13 |
| L-Ornithine | 1.61 | 1.61 | 0.69 | 5.00E-07 |
| L-Tryptophan | 2.37 | 2.37 | 1.24 | 2.50E-05 |
| L-Methionine | 1.77 | 1.77 | 0.82 | 3.16E-05 |
| Choline | 0.59 | 1.69 | -0.76 | 3.16E-05 |
| 3-Hydroxybutyric acid | 1.74 | 1.74 | 0.80 | 0.0003 |
| Pyruvic acid | 0.49 | 2.04 | -1.03 | 0.001 |
| ***Metabolites PARK2-PINK1-PARK7_F*** | **FC** | **\|FC\|** | **log2(FC)** | **p.value** |
| L-Leucine | 1.70 | 1.70 | 0.77 | 1.58E-09 |
| Choline | 0.63 | 1.59 | -0.67 | 3.98E-07 |
| 3-Hydroxybutyric acid | 1.84 | 1.84 | 0.88 | 0.0002 |
| Acetic acid | 0.46 | 2.17 | -1.12 | 0.0002 |
| L-Proline | 2.14 | 2.14 | 1.10 | 0.003 |

Results from univariate analysis were presented using a Robust Volcano plot of metabolite concentrations detected in the 1d-CPMG NMR spectrum. The analysis combines the Fold change (FC) test, calculated as the ratio of average concentrations between the PARK2-PINK1-PARK7 _M/HC_M and PARK2-PINK1-PARK7 _F/HC_F clusters—using a threshold of absolute fold change |FC| > 1.5—and the p-value from the T-Test, which is considered significant if less than 0.05.

**Table S11. Pathway enrichment analysis of male PARK2-PINK1-PARK7 carriers**

| | Hits | Raw p | Holm p | FDR | Metabolites |
|---|---|---|---|---|---|
| Cysteine Metabolism | 2 | 7.22E-13 | 4.98E-10 | 1.73E-10 | L-Glutamic acid, Pyruvic acid. |
| Nicotinate and Nicotinamide Metabolism | 2 | 7.43E-12 | 5.05E-10 | 1.73E-10 | L-Glutamic acid, L-Glutamine. |
| Amino Sugar Metabolism | 4 | 5.22E-11 | 3.50E-08 | 9.13E-09 | L-Glutamic acid, Pyruvic acid, Acetoacetic acid, L-Glutamine. |
| Malate-Aspartate Shuttle | 2 | 3.61E-09 | 2.38E-07 | 5.06E-08 | Aspartate, L-Glutamic acid. |
| Glycine and Serine Metabolism | 11 | 4.95E-08 | 3.22E-07 | 5.78E-07 | Betaine, Creatine, Glycine, L-Glutamic acid, L-Alanine, L-Threonine, L-Serine, L-Ornithine, Pyruvic acid, L-Methionine, L-Arginine. |
| Lysine Degradation | 2 | 8.27E-08 | 5.30E-06 | 8.27E-07 | Choline, L-Serine. |
| Urea Cycle | 8 | 2.40E-07 | 1.51E-05 | 2.10E-06 | L-Glutamic acid, L-Alanine, Aspartate,L- Ornithine, Pyruvic acid, Urea, L-Arginine, L-Glutamine. |
| Tryptophan Metabolism | 4 | 8.16E-06 | 5.06E-04 | 6.34E-05 | L-Glutamic acid, Tryptophan, Formic acid, L-Alanine. |
| Aspartate Metabolism | 6 | 1.79E-05 | 1.09E-07 | 1.25E-04 | Acetic acid, L-Glutamic acid, L-Asparagine, Aspartate, L-Arginine, L-Glutamine. |
| Tyrosine Metabolism | 4 | 2.91E-05 | 1.75E-03 | 1.85E-04 | Tyrosine, Acetic acid, Aspartate, L-Glutamic acid. |
| Alanine Metabolism | 4 | 6.23E-05 | 3.68E-03 | 3.63E-05 | Glycine, L-Glutamic acid, L-Alanine, Pyruvic acid. |
| Spermidine and Spermine Biosynthesis | 2 | 1.83E-04 | 1.06E-02 | 9.84E-04 | L-Ornithine, L-Methionine. |
| Ammonia Recycling | 8 | 2.89E-04 | 1.65E-02 | 1.44E-03 | Glycine, L-Glutamic acid,L-Asparagine, Histidine, L-Serine, Aspartate, Pyruvic acid,L-Glutamine. |
| Glutamate Metabolism | 7 | 3.08E-04 | 1.72E-02 | 1.44E-03 | L-Glutamic acid, Pyruvic acid, Aspartate, Glycine, L-Alanine, L-Glutamine, Succinic acid. |
| Arginine and Proline Metabolism | 9 | 5.57E-04 | 3.07E-02 | 2.44E-03 | L-Glutamic acid, Proline, Aspartate Succinic acid, Urea, L-Arginine, L-Ornithine, Creatine, Glycine. |
| Glutathione Metabolism | 3 | 7.03E-04 | 3.80E-02 | 2.90E-03 | Glycine, L-Glutamic acid, L-Ornithine. |
| Glucose-Alanine Cycle | 4 | 7.88E-04 | 4.17E-02 | 3.06E-03 | D-Glucose, L-Glutamic acid, L-Alanine, Pyruvic acid. |
| Histidine Metabolism | 3 | 1.01E-03 | 5.25E-02 | 3.72E-03 | 1-Methylhistidine, L-Glutamic acid, Histidine. |

| | | | | | |
|---|---|---|---|---|---|
| Propanoate Metabolism | 3 | 1.84E-03 | 9.22E-02 | 6.15E-03 | 2-Hydroxybutyric acid, L-Glutamic acid, Valine. |
| Phenylalanine and Tyrosine Metabolism | 4 | 1.99E-03 | 9.75E-02 | 6.33E-03 | Tyrosine, Acetic acid, Aspartate, L-Glutamic acid. |
| Beta-Alanine Metabolism | 3 | 2.64E-03 | 1.27E-02 | 8.02E-03 | L-Glutamic acid, Histidine, Aspartate. |

Pathway enrichment analysis performed on $^1$H-NMR and related to male and female genetic Parkinson's disease (PD) patients with PARK2-PINK1-PARK7 mutations and HC serum metabolomic results. Hits represent the numbers of metabolites involved in the pathways and specified in the 'Metabolites' column. The pathways were considered statistically significant with Hits >1, p-value < 0.05, adjusted p-value performed using Holm Bonferroni test (Holm p) and False Discovery Rate (FDR) < 1.

**Table S12. Pathway enrichment analysis of female PARK2-PINK1-PARK7 carriers.**

| | Hits | Raw p | Holm p | FDR | Metabolites |
|---|---|---|---|---|---|
| Valine Leucine and Isoleucine Degradation | 6 | 3.26E-06 | 2.28E-03 | 2.28E-03 | Succinic acid, Valine, L-Leucine, Acetoacetic acid, L-Glutamic acid, L-Isoleucine. |
| Betaine Metabolism | 3 | 1.10E-05 | 7.62E-02 | 3.86E-02 | Betaine, L-Methionine, Choline. |
| Methionine Metabolism | 5 | 6.43E-05 | 4.24E-02 | 8.03E-02 | Betaine, L-Methionine, Choline, Glycine, L-Serine. |
| Pyruvate Metabolism | 3 | 1.01E-04 | 6.29E-03 | 7.89E-04 | Acetic acid, Lactic acid, Pyruvic acid. |
| Ketone Body Metabolism | 4 | 2.63E-03 | 1.58E-02 | 1.68E-02 | 3-Hydroxybutyric acid, Acetoacetic acid, Succinic acid, Acetone. |
| Transfer of Acetyl Groups into Mitochondria | 3 | 3.19E-03 | 1.88E-02 | 1.86E-02 | Citric acid, Pyruvic acid, Glucose. |
| Propanoate Metabolism | 3 | 4.03E-03 | 2.34E-02 | 2.17E-02 | 2-Hydroxybutyric acid, L-Glutamic acid, Valine. |
| Aspartate Metabolism | 6 | 5.20E-03 | 2.97E-02 | 2.48E-02 | L-Glutamic acid, Acetic acid, L-Asparagine, Aspartate, L-Glutamine, L-Arginine |
| Citric Acid Cycle | 3 | 5.30E-03 | 9.70E-02 | 2.48E-02 | Citric acid, Pyruvic acid, Succinic acid. |

Pathway enrichment analysis performed on $^1$H-NMR and related to male and female gPD with PARK2-PINK1-PARK7 mutations and HC serum metabolomic results. Hits represent the numbers of metabolites involved in the pathways and specified in the 'Metabolites' column. The pathways were considered statistically significant with Hits >1, p-value < 0.05, adjusted p-value performed using Holm Bonferroni test (Holm p) and False Discovery Rate (FDR) < 1.

**Table S13. Robust Volcano plot results related to Parkinson's disease patients with *TMEM175* mutation compared to age- and sex-matched healthy controls.**

| ***Metabolites TMEM175_M*** | ***FC*** | **\|FC\|** | **log2(FC)** | ***p.value*** |
|---|---|---|---|---|
| L-Glutamic acid | 0.42 | 2.38 | -1.25 | 1.0E-09 |
| Choline | 0.53 | 1.89 | -0.92 | 1.0E-05 |
| Malonate | 0.59 | 1.69 | -0.76 | 4.73E-05 |
| Acetic acid | 0.46 | 2.17 | -1.12 | 0.0002 |
| Pyruvic acid | 0.54 | 1.85 | -0.89 | 0.0003 |
| L-Tryptophan | 2.37 | 2.37 | 1.24 | 5.62E-07 |
| L-Methionine | 1.62 | 1.62 | 0.70 | 0.0003 |
| ***Metabolites TMEM175_F*** | ***FC*** | **\|FC\|** | **log2(FC)** | ***p.value*** |
| Choline | 0.66 | 1.52 | -0.60 | 2.82E-05 |
| L-Ornithine | 0.46 | 2.17 | -1.12 | 6.30E-05 |
| Pyruvic acid | 0.54 | 1.85 | -0.89 | 0.0056 |
| Acetic acid | 0.57 | 1.75 | -0.81 | 0.02 |
| L-Methionine | 1.55 | 1.55 | 0.63 | 0.01 |
| 3-Hydroxybutyric acid | 2.18 | 2.18 | 1.12 | 0.002 |
| L-Threonine | 1.54 | 1.54 | 0.62 | 7.08E-05 |
| L-Alanine | 2.59 | 2.59 | 1.37 | 1.78E-05 |

Results from univariate analysis were presented using a Robust Volcano plot of metabolite concentrations detected in the 1d-CPMG NMR spectrum. The analysis combines the Fold change (FC) test, calculated as the ratio of average concentrations between the TMEM175 _M/HC_M and TMEM175 _F/HC_F clusters—using a threshold of absolute fold change |FC| > 1.51—and the p-value from the T-Test, which is considered significant if less than 0.05.

**Table S14. Pathway enrichment analysis of male *TMEM175* carriers**

| | Hits | Raw p | Holm p | FDR | Metabolites |
|---|---|---|---|---|---|
| Tryptophan Metabolism | 4 | 1.47E-08 | 1.03E-05 | 1.03E-02 | L-Glutamic acid, Tryptophan, Formic acid, L-Alanine. |
| Cysteine Metabolism | 2 | 4.61E-05 | 3.09E-04 | 8.07E-03 | L-Glutamic acid, Pyruvate. |
| Malate-Aspartate Shuttle | 2 | 1.04E-04 | 6.78E-04 | 1.22E-04 | Aspartate, L-Glutamic acid. |
| Alanine Metabolism | 4 | 1.20E-04 | 6.84E-03 | 6.00E-04 | Glycine, L-Glutamic acid, L-Alanine, Pyruvate. |
| Glutathione Metabolism | 3 | 1.53E-04 | 8.59E-03 | 7.16E-04 | Glycine, L-Glutamic acid, L-Ornithine. |
| Propanoate Metabolism | 3 | 2.68E-04 | 1.71E-03 | 2.36E-04 | 2-Hydroxybutyric acid, L-Glutamic acid, L-Valine. |
| Urea Cycle | 8 | 6.25E-04 | 3.44E-02 | 2.74E-03 | L-Glutamic acid, L-Alanine, Aspartate, L-Ornithine, Pyruvate, Urea, Arginine, L-Glutamine. |
| Glutamate Metabolism | 7 | 7.02E-04 | 3.79E-02 | 2.89E-03 | L-Glutamic acid, Pyruvate, Aspartate, Glycine, L-Alanine, L-Glutamine, Succinic acid |
| Glucose-Alanine Cycle | 4 | 3.42E-03 | 2.12E-03 | 2.36E-04 | D-Glucose, L-Glutamic acid, L-Alanine, Pyruvate. |
| Lysine Degradation | 2 | 3.49E-03 | 4.13E-03 | 2.36E-04 | L-Glutamic acid, L-Lysine. |
| Histidine Metabolism | 3 | 3.71E-03 | 2.22E-03 | 2.36E-04 | L-Glutamic acid, L-Histidine, 1-Methylhistidine. |
| Amino Sugar Metabolism | 4 | 4.53E-03 | 2.67E-03 | 2.58E-04 | L-Glutamic acid, Hypoxanthine, L-Glutamine, Aspartate. |
| Purine Metabolism | 5 | 4.79E-03 | 2.78E-03 | 2.58E-04 | L-Glutamic acid, Hypoxanthine, L-Glutamine, Aspartate, Glycine. |
| Folate Metabolism | 2 | 5.85E-03 | 3.86E-04 | 8.19E-03 | Formic acid, L-Glutamic acid. |

Pathway enrichment analysis performed on $^1$H-NMR and related to male and female gPD genetic Parkinson’s disease (gPD) patients with TMEM175 mutations and HC serum metabolomic results. Hits represent the number of metabolites involved in the pathways and specified in the ‘Metabolites’ column. The pathways were considered statistically significant with Hits >1, p-value < 0.05, adjusted p-value performed using Holm Bonferroni test (Holm p) and False Discovery Rate (FDR) < 1.

### Table S15. Pathway enrichment analysis of female *TMEM175* carriers

| | Hits | Raw p | Holm p | FDR | Metabolites |
|---|---|---|---|---|---|
| Glycine and Serine Metabolism | 11 | 7.12E-05 | 4.99E-03 | 4.99E-03 | Betaine, Creatine,Glycine, L-Glutamic acid, L-Alanine, L-Threonine, L-Serine, L-Ornithine, Pyruvic acid,L-Arginine, L-Methionine. |
| Phosphatidylethanolamine Biosynthesis | 2 | 1.12E-04 | 7.42E-04 | 1.54E-04 | Choline,L-Serine. |
| Spermidine and Spermine Biosynthesis | 2 | 1.32E-04 | 8.60E-04 | 1.54E-04 | L-Ornithine, L-Methionine. |
| Selenoamino Acid Metabolism | 2 | 3.95E-04 | 2.73E-04 | 1.31E-04 | L-Alanine, L-Serine. |
| Betaine Metabolism | 3 | 4.32E-04 | 2.68E-03 | 3.36E-04 | Betaine, Choline, L-Methionine. |
| Methionine Metabolism | 5 | 7.25E-04 | 4.93E-04 | 1.31E-04 | Betaine,Glycine, L-Methionine,L-Serine,Choline. |
| Urea Cycle | 8 | 7.47E-04 | 5.00E-04 | 1.31E-04 | L-Glutamic acid, L-Alanine, Aspartate,L-Ornithine,Pyruvic acid, Urea, L-Arginine,L-Glutamine. |
| Alanine Metabolism | 4 | 1.10E-03 | 6.58E-03 | 6.98E-04 | Glycine, L-Glutamic acid, L-Alanine,Pyruvic acid |
| Ammonia Recycling | 8 | 7.15E-03 | 4.22E-02 | 3.96E-03 | Glycine, L-Glutamic acid, L-Asparagine, Histidine, L-Serine, Aspartate, Pyruvic acid, L-Glutamine |
| Fatty Acid Biosynthesis | 3 | 7.35E-03 | 4.26E-02 | 3.96E-03 | Acetic acid, Acetoacetic acid, 3-Hydroxybutyric acid |

Pathway enrichment analysis performed on H-NMR and related to male and female genetic Parkinson’s disease (gPD) patients with TMEM175 mutations and HC serum metabolomic results. Hits represent the number of metabolites involved in the pathways and specified in the ‘Metabolites’ column. The pathways were considered statistically significant with Hits >1, p-value < 0.05, adjusted p-value performed using Holm Bonferroni test (Holm p) and False Discovery Rate (FDR) < 1.

**Supplementary Table 16. Linear model results: group × sex interaction on serum amino acid levels (full cohort, HC vs gPD).**

| Outcome | Term | β | SE | t | p (raw) | p (FDR) |
|---|---|---|---|---|---|---|
| **1-Methylhistidine** | Intercept | 5.904 | 0.225 | 26.229 | < 0.001 | |
| | Group (gPD vs HC) | -0.055 | 0.102 | -0.533 | 0.5938 | |
| | Sex (M vs F) | 0.252 | 0.144 | 1.746 | 0.0808 | |
| | Age | -0.006 | 0.003 | -2.132 | 0.0330 | |
| | **Group × Sex (interaction)** | **-0.224** | **0.159** | **-1.412** | **0.1579** | 0.7193 |
| **2-Hydroxybutyric acid** | Intercept | 6.661 | 0.261 | 25.480 | < 0.001 | |
| | Group (gPD vs HC) | -0.465 | 0.101 | -4.621 | < 0.001 | |
| | Sex (M vs F) | 0.000 | 0.157 | -0.001 | 0.9990 | |
| | Age | -0.008 | 0.004 | -2.196 | 0.0281 | |
| | **Group × Sex (interaction)** | **0.117** | **0.178** | **0.660** | **0.5094** | 0.9603 |
| **Acetic acid** | Intercept | 3.070 | 0.398 | 7.719 | < 0.001 | |
| | Group (gPD vs HC) | -0.611 | 0.205 | -2.980 | 0.0029 | |
| | Sex (M vs F) | 0.013 | 0.294 | 0.045 | 0.9637 | |
| | Age | 0.003 | 0.006 | 0.615 | 0.5388 | |
| | **Group × Sex (interaction)** | **-0.179** | **0.311** | **-0.574** | **0.5659** | 0.9603 |
| **Betaine** | Intercept | 3.630 | 0.231 | 15.726 | < 0.001 | |
| | Group (gPD vs HC) | -0.030 | 0.108 | -0.276 | 0.7828 | |

| Outcome | Term | β | SE | t | p (raw) | p (FDR) |
|---|---|---|---|---|---|---|
| | Sex (M vs F) | 0.110 | 0.115 | 0.954 | 0.3400 | |
| | Age | 0.000 | 0.003 | 0.122 | 0.9032 | |
| | **Group × Sex (interaction)** | **-0.031** | **0.136** | **-0.231** | **0.8174** | 0.9603 |
| **Acetoacetate** | Intercept | 3.362 | 0.215 | 15.625 | < 0.001 | |
| | Group (gPD vs HC) | -0.187 | 0.084 | -2.237 | 0.0253 | |
| | Sex (M vs F) | -0.056 | 0.111 | -0.508 | 0.6111 | |
| | Age | -0.002 | 0.003 | -0.842 | 0.3999 | |
| | **Group × Sex (interaction)** | **-0.001** | **0.124** | **-0.009** | **0.9926** | 0.9926 |
| **Carnitine** | Intercept | 5.700 | 0.170 | 33.485 | < 0.001 | |
| | Group (gPD vs HC) | -0.104 | 0.072 | -1.442 | 0.1494 | |
| | Sex (M vs F) | -0.172 | 0.101 | -1.697 | 0.0896 | |
| | Age | -0.003 | 0.002 | -1.410 | 0.1584 | |
| | **Group × Sex (interaction)** | **0.010** | **0.114** | **0.087** | **0.9303** | 0.9603 |
| **Creatine** | Intercept | 4.769 | 0.217 | 21.942 | < 0.001 | |
| | Group (gPD vs HC) | -0.161 | 0.103 | -1.556 | 0.1197 | |
| | Sex (M vs F) | -0.138 | 0.148 | -0.926 | 0.3544 | |
| | Age | -0.009 | 0.003 | -2.723 | 0.0065 | |
| | **Group × Sex (interaction)** | **0.122** | **0.162** | **0.752** | **0.4521** | 0.9603 |

| Outcome | Term | β | SE | t | p (raw) | p (FDR) |
|---|---|---|---|---|---|---|
| **Citric acid** | Intercept | 4.385 | 0.192 | 22.810 | < 0.001 | |
| | Group (gPD vs HC) | 0.266 | 0.091 | 2.910 | 0.0036 | |
| | Sex (M vs F) | 0.203 | 0.106 | 1.907 | 0.0565 | |
| | Age | 0.000 | 0.002 | -0.197 | 0.8437 | |
| | **Group × Sex (interaction)** | **-0.320** | **0.114** | **-2.801** | **0.0051** | 0.0696 |
| **Choline** | Intercept | 5.105 | 0.189 | 27.030 | < 0.001 | |
| | Group (gPD vs HC) | -0.624 | 0.079 | -7.862 | < 0.001 | |
| | Sex (M vs F) | 0.060 | 0.098 | 0.611 | 0.5410 | |
| | Age | -0.004 | 0.003 | -1.412 | 0.1580 | |
| | **Group × Sex (interaction)** | **-0.097** | **0.119** | **-0.816** | **0.4146** | 0.9603 |
| **D-Glucose** | Intercept | 7.696 | 0.178 | 43.336 | < 0.001 | |
| | Group (gPD vs HC) | 0.061 | 0.059 | 1.028 | 0.3038 | |
| | Sex (M vs F) | 0.182 | 0.106 | 1.712 | 0.0869 | |
| | Age | -0.001 | 0.002 | -0.514 | 0.6071 | |
| | **Group × Sex (interaction)** | **-0.140** | **0.119** | **-1.177** | **0.2394** | 0.9505 |
| **Tyrosine** | Intercept | 5.108 | 0.188 | 27.218 | < 0.001 | |
| | Group (gPD vs HC) | 0.015 | 0.071 | 0.207 | 0.8361 | |
| | Sex (M vs F) | 0.056 | 0.114 | 0.487 | 0.6262 | |
| | Age | -0.003 | 0.003 | -1.053 | 0.2921 | |

| Outcome | Term | β | SE | t | p (raw) | p (FDR) |
|---|---|---|---|---|---|---|
| | **Group × Sex (interaction)** | **-0.217** | **0.134** | **-1.618** | **0.1057** | 0.6191 |
| **Formate** | Intercept | 3.560 | 0.365 | 9.746 | < 0.001 | |
| | Group (gPD vs HC) | 0.046 | 0.139 | 0.328 | 0.7428 | |
| | Sex (M vs F) | -0.078 | 0.214 | -0.367 | 0.7136 | |
| | Age | -0.004 | 0.005 | -0.739 | 0.4596 | |
| | **Group × Sex (interaction)** | **-0.063** | **0.232** | **-0.271** | **0.7865** | 0.9603 |
| **L-Glutamic acid** | Intercept | 4.021 | 0.494 | 8.139 | < 0.001 | |
| | Group (gPD vs HC) | -0.305 | 0.176 | -1.738 | 0.0822 | |
| | Sex (M vs F) | -0.805 | 0.124 | -6.495 | < 0.001 | |
| | Age | 0.004 | 0.007 | 0.515 | 0.6064 | |
| | **Group × Sex (interaction)** | **-0.680** | **0.185** | **-3.673** | **< 0.001** | **0.0098** |
| **Hypoxanthine** | Intercept | -1.599 | 0.493 | -3.240 | 0.0012 | |
| | Group (gPD vs HC) | -0.038 | 0.198 | -0.190 | 0.8494 | |
| | Sex (M vs F) | -0.037 | 0.258 | -0.143 | 0.8866 | |
| | Age | 0.004 | 0.007 | 0.571 | 0.5681 | |
| | **Group × Sex (interaction)** | **0.101** | **0.288** | **0.352** | **0.7249** | 0.9603 |
| **Glycine** | Intercept | 2.362 | 0.265 | 8.899 | < 0.001 | |
| | Group (gPD vs HC) | -0.186 | 0.133 | -1.400 | 0.1615 | |

| Outcome | Term | β | SE | t | p (raw) | p (FDR) |
|---|---|---|---|---|---|---|
| | Sex (M vs F) | -0.143 | 0.166 | -0.866 | 0.3866 | |
| | Age | 0.003 | 0.004 | 0.756 | 0.4494 | |
| | **Group × Sex (interaction)** | **0.083** | **0.183** | **0.454** | **0.6499** | 0.9603 |
| **L-Phenylalanine** | Intercept | 2.502 | 0.331 | 7.553 | < 0.001 | |
| | Group (gPD vs HC) | 0.050 | 0.137 | 0.362 | 0.7175 | |
| | Sex (M vs F) | 0.139 | 0.181 | 0.766 | 0.4436 | |
| | Age | 0.003 | 0.004 | 0.721 | 0.4707 | |
| | **Group × Sex (interaction)** | **-0.186** | **0.193** | **-0.967** | **0.3335** | 0.9603 |
| **L-Alanine** | Intercept | 4.097 | 0.255 | 16.053 | < 0.001 | |
| | Group (gPD vs HC) | -0.020 | 0.106 | -0.187 | 0.8519 | |
| | Sex (M vs F) | 0.132 | 0.117 | 1.128 | 0.2592 | |
| | Age | 0.003 | 0.004 | 0.779 | 0.4361 | |
| | **Group × Sex (interaction)** | **-0.230** | **0.150** | **-1.534** | **0.1251** | 0.6413 |
| **L-Proline** | Intercept | 6.472 | 0.257 | 25.174 | < 0.001 | |
| | Group (gPD vs HC) | 0.156 | 0.095 | 1.647 | 0.0997 | |
| | Sex (M vs F) | 0.182 | 0.101 | 1.798 | 0.0722 | |
| | Age | 0.001 | 0.003 | 0.315 | 0.7524 | |
| | **Group × Sex (interaction)** | **-0.237** | **0.107** | **-2.217** | **0.0266** | 0.2728 |

| Outcome | Term | β | SE | t | p (raw) | p (FDR) |
|---|---|---|---|---|---|---|
| **L-Threonine** | Intercept | 6.656 | 0.248 | 26.851 | < 0.001 | |
| | Group (gPD vs HC) | -0.014 | 0.124 | -0.111 | 0.9119 | |
| | Sex (M vs F) | 0.042 | 0.137 | 0.305 | 0.7607 | |
| | Age | -0.003 | 0.003 | -0.999 | 0.3177 | |
| | **Group × Sex (interaction)** | **-0.065** | **0.146** | **-0.444** | **0.6571** | 0.9603 |
| **L-Ornithine** | Intercept | 3.508 | 0.467 | 7.510 | < 0.001 | |
| | Group (gPD vs HC) | 0.193 | 0.151 | 1.282 | 0.1997 | |
| | Sex (M vs F) | 0.075 | 0.152 | 0.492 | 0.6226 | |
| | Age | 0.007 | 0.005 | 1.402 | 0.1610 | |
| | **Group × Sex (interaction)** | **0.026** | **0.168** | **0.155** | **0.8766** | 0.9603 |
| **Isoleucine** | Intercept | 5.237 | 0.187 | 28.052 | < 0.001 | |
| | Group (gPD vs HC) | -0.168 | 0.073 | -2.314 | 0.0207 | |
| | Sex (M vs F) | 0.007 | 0.091 | 0.080 | 0.9362 | |
| | Age | -0.004 | 0.003 | -1.426 | 0.1537 | |
| | **Group × Sex (interaction)** | **-0.103** | **0.109** | **-0.948** | **0.3432** | 0.9603 |
| **L-Histidine** | Intercept | 5.695 | 0.218 | 26.155 | < 0.001 | |
| | Group (gPD vs HC) | -0.027 | 0.084 | -0.316 | 0.7519 | |
| | Sex (M vs F) | -0.027 | 0.105 | -0.257 | 0.7973 | |
| | Age | -0.004 | 0.003 | -1.321 | 0.1864 | |

| Outcome | Term | β | SE | t | p (raw) | p (FDR) |
|---|---|---|---|---|---|---|
| | **Group × Sex (interaction)** | **0.099** | **0.122** | **0.813** | **0.4161** | 0.9603 |
| **L-Lysine** | Intercept | 3.751 | 0.420 | 8.935 | < 0.001 | |
| | Group (gPD vs HC) | 0.058 | 0.264 | 0.222 | 0.8247 | |
| | Sex (M vs F) | 0.101 | 0.264 | 0.382 | 0.7025 | |
| | Age | -0.004 | 0.007 | -0.629 | 0.5296 | |
| | **Group × Sex (interaction)** | **-0.213** | **0.296** | **-0.719** | **0.4720** | 0.9603 |
| **L-Methionine** | Intercept | 5.065 | 0.329 | 15.392 | < 0.001 | |
| | Group (gPD vs HC) | 0.522 | 0.113 | 4.600 | < 0.001 | |
| | Sex (M vs F) | 0.278 | 0.119 | 2.335 | 0.0196 | |
| | Age | -0.003 | 0.004 | -0.740 | 0.4593 | |
| | **Group × Sex (interaction)** | **-0.175** | **0.161** | **-1.084** | **0.2782** | 0.9505 |
| **L-Lactic acid** | Intercept | 7.761 | 0.220 | 35.327 | < 0.001 | |
| | Group (gPD vs HC) | -0.117 | 0.110 | -1.067 | 0.2859 | |
| | Sex (M vs F) | 0.148 | 0.153 | 0.966 | 0.3340 | |
| | Age | -0.007 | 0.003 | -2.113 | 0.0346 | |
| | **Group × Sex (interaction)** | **0.037** | **0.166** | **0.222** | **0.8241** | 0.9603 |
| **Aspartate** | Intercept | 4.786 | 0.391 | 12.239 | < 0.001 | |
| | Group (gPD vs HC) | -0.278 | 0.106 | -2.628 | 0.0086 | |

| Outcome | Term | β | SE | t | p (raw) | p (FDR) |
|---|---|---|---|---|---|---|
| | Sex (M vs F) | -0.148 | 0.196 | -0.753 | 0.4516 | |
| | Age | -0.005 | 0.006 | -0.938 | 0.3483 | |
| | **Group × Sex (interaction)** | **0.026** | **0.211** | **0.123** | **0.9021** | 0.9603 |
| **L-Asparagine** | Intercept | 2.175 | 0.306 | 7.105 | < 0.001 | |
| | Group (gPD vs HC) | -0.529 | 0.151 | -3.502 | < 0.001 | |
| | Sex (M vs F) | -0.208 | 0.138 | -1.502 | 0.1331 | |
| | Age | -0.001 | 0.004 | -0.233 | 0.8159 | |
| | **Group × Sex (interaction)** | **0.057** | **0.182** | **0.313** | **0.7543** | 0.9603 |
| **Pyruvic acid** | Intercept | 2.895 | 0.507 | 5.709 | < 0.001 | |
| | Group (gPD vs HC) | -0.721 | 0.156 | -4.620 | < 0.001 | |
| | Sex (M vs F) | -0.011 | 0.143 | -0.076 | 0.9394 | |
| | Age | 0.006 | 0.007 | 0.799 | 0.4244 | |
| | **Group × Sex (interaction)** | **-0.056** | **0.223** | **-0.249** | **0.8036** | 0.9603 |
| **Succinate** | Intercept | 3.873 | 0.717 | 5.401 | < 0.001 | |
| | Group (gPD vs HC) | 0.120 | 0.406 | 0.296 | 0.7674 | |
| | Sex (M vs F) | 0.500 | 0.358 | 1.398 | 0.1620 | |
| | Age | -0.012 | 0.010 | -1.132 | 0.2575 | |
| | **Group × Sex (interaction)** | **-0.189** | **0.448** | **-0.422** | **0.6732** | 0.9603 |

| Outcome | Term | β | SE | t | p (raw) | p (FDR) |
|---|---|---|---|---|---|---|
| **Urea** | Intercept | 10.084 | 0.690 | 14.614 | < 0.001 | |
| | Group (gPD vs HC) | -0.074 | 0.287 | -0.259 | 0.7955 | |
| | Sex (M vs F) | 0.240 | 0.321 | 0.749 | 0.4540 | |
| | Age | -0.018 | 0.009 | -1.959 | 0.0502 | |
| | **Group × Sex (interaction)** | **-0.084** | **0.374** | **-0.224** | **0.8225** | 0.9603 |
| **3-Hydroxybutyric acid** | Intercept | 2.695 | 0.388 | 6.950 | < 0.001 | |
| | Group (gPD vs HC) | 0.492 | 0.129 | 3.821 | < 0.001 | |
| | Sex (M vs F) | 0.138 | 0.153 | 0.899 | 0.3687 | |
| | Age | 0.002 | 0.006 | 0.387 | 0.6985 | |
| | **Group × Sex (interaction)** | **-0.099** | **0.182** | **-0.542** | **0.5878** | 0.9603 |
| **L-Arginine** | Intercept | 2.152 | 0.404 | 5.323 | < 0.001 | |
| | Group (gPD vs HC) | -0.330 | 0.157 | -2.106 | 0.0352 | |
| | Sex (M vs F) | -0.132 | 0.174 | -0.762 | 0.4461 | |
| | Age | 0.002 | 0.005 | 0.428 | 0.6684 | |
| | **Group × Sex (interaction)** | **0.222** | **0.203** | **1.089** | **0.2760** | 0.9505 |
| **Creatinine** | Intercept | 4.675 | 0.259 | 18.024 | < 0.001 | |
| | Group (gPD vs HC) | -0.044 | 0.088 | -0.496 | 0.6198 | |
| | Sex (M vs F) | 0.016 | 0.099 | 0.160 | 0.8727 | |
| | Age | -0.005 | 0.003 | -1.332 | 0.1830 | |

| Outcome | Term | β | SE | t | p (raw) | p (FDR) |
|---|---|---|---|---|---|---|
| | **Group × Sex (interaction)** | **-0.043** | **0.118** | **-0.368** | **0.7128** | 0.9603 |
| **L-Tryptophan** | Intercept | 2.799 | 0.269 | 10.391 | < 0.001 | |
| | Group (gPD vs HC) | 0.608 | 0.070 | 8.700 | < 0.001 | |
| | Sex (M vs F) | -0.124 | 0.062 | -1.986 | 0.0470 | |
| | Age | 0.004 | 0.004 | 0.912 | 0.3615 | |
| | **Group × Sex (interaction)** | **-0.054** | **0.106** | **-0.511** | **0.6093** | 0.9603 |
| **L-Leucine** | Intercept | 1.952 | 0.330 | 5.909 | < 0.001 | |
| | Group (gPD vs HC) | 0.069 | 0.092 | 0.759 | 0.4479 | |
| | Sex (M vs F) | 0.141 | 0.192 | 0.734 | 0.4632 | |
| | Age | 0.012 | 0.005 | 2.393 | 0.0167 | |
| | **Group × Sex (interaction)** | **-0.061** | **0.223** | **-0.272** | **0.7854** | 0.9603 |
| **Malonate** | Intercept | 4.976 | 0.306 | 16.272 | < 0.001 | |
| | Group (gPD vs HC) | -0.287 | 0.181 | -1.580 | 0.1141 | |
| | Sex (M vs F) | 0.256 | 0.177 | 1.449 | 0.1472 | |
| | Age | -0.005 | 0.004 | -1.132 | 0.2578 | |
| | **Group × Sex (interaction)** | **-0.404** | **0.210** | **-1.924** | **0.0543** | 0.3711 |
| **L-Serine** | Intercept | 1.672 | 0.403 | 4.144 | < 0.001 | |
| | Group (gPD vs HC) | 0.422 | 0.141 | 2.985 | 0.0028 | |

| Outcome | Term | β | SE | t | p (raw) | p (FDR) |
|---|---|---|---|---|---|---|
| | Sex (M vs F) | 0.502 | 0.206 | 2.431 | 0.0151 | |
| | Age | -0.001 | 0.006 | -0.166 | 0.8683 | |
| | **Group × Sex (interaction)** | **-0.734** | **0.238** | **-3.080** | **0.0021** | **0.0424** |
| **Valine** | Intercept | 4.511 | 0.526 | 8.580 | < 0.001 | |
| | Group (gPD vs HC) | 0.300 | 0.321 | 0.936 | 0.3495 | |
| | Sex (M vs F) | 0.394 | 0.407 | 0.967 | 0.3333 | |
| | Age | -0.006 | 0.009 | -0.648 | 0.5168 | |
| | **Group × Sex (interaction)** | **-0.290** | **0.435** | **-0.667** | **0.5049** | 0.9603 |
| **L-Glutamine** | Intercept | 2.941 | 0.822 | 3.576 | < 0.001 | |
| | Group (gPD vs HC) | 0.393 | 0.313 | 1.253 | 0.2101 | |
| | Sex (M vs F) | 0.567 | 0.314 | 1.804 | 0.0712 | |
| | Age | 0.005 | 0.009 | 0.514 | 0.6073 | |
| | **Group × Sex (interaction)** | **-0.739** | **0.354** | **-2.090** | **0.0367** | 0.3006 |
| **Acetone** | Intercept | 4.955 | 0.276 | 17.936 | < 0.001 | |
| | Group (gPD vs HC) | -0.038 | 0.104 | -0.371 | 0.7108 | |
| | Sex (M vs F) | 0.053 | 0.139 | 0.381 | 0.7033 | |
| | Age | -0.005 | 0.004 | -1.250 | 0.2113 | |
| | **Group × Sex (interaction)** | **-0.013** | **0.158** | **-0.079** | **0.9369** | 0.9603 |

| Outcome | Term | β | SE | t | p (raw) | p (FDR) |
|---|---|---|---|---|---|---|
| **Isobutyric acid** | Intercept | 5.216 | 1.280 | 4.074 | < 0.001 | |
| | Group (gPD vs HC) | 1.075 | 0.619 | 1.737 | 0.0823 | |
| | Sex (M vs F) | -0.068 | 0.828 | -0.083 | 0.9341 | |
| | Age | -0.057 | 0.019 | -2.976 | 0.0029 | |
| | **Group × Sex (interaction)** | **-0.095** | **0.918** | **-0.103** | **0.9180** | 0.9603 |

Results from ordinary least squares (OLS) linear models fitted separately for each of the 41 amino acid outcomes. Model formula: log(amino acid) ~ group + sex + age + group:sex, where group = HC (0) vs gPD (1) and sex = female (0) vs male (1). Coefficients (β), robust standard errors (SE), t-statistics, and raw p-values are reported for all model terms. The p (FDR) column reports Benjamini-Hochberg false discovery rate-corrected p-values for the interaction term only, derived from correction across the 41 interaction tests. Highlighted cells in the p (FDR) column indicate significance at $p < 0.05$ after FDR correction. All inference is based on heteroscedasticity-consistent (HC3) robust standard errors. n = 166.

*Model: log(amino acid) ~ group + sex + age + group:sex. group: 0 = HC, 1 = gPD; sex: 0 = female, 1 = male. Inference based on heteroscedasticity-consistent robust standard errors (HC3). p (FDR): Benjamini-Hochberg correction applied to the 41 interaction p-values. Highlighted cells indicate significance at p < 0.05 after FDR correction.*

**Checklist**

**1. Reporting Guidelines**
-STROBE Compliance: This study was designed and reported following the Strengthening the Reporting of Observational Studies in Epidemiology (STROBE) guidelines for case-control studies. **Pag. 26 lines 18-19**

-Metabolomics Reporting: Metabolomic data acquisition and processing comply with the Metabolomics Standards Initiative (MSI) guidelines. **Pag. 26 lines 18-19**

**2. Ethical Compliance & Registration**
**-**Institutional Review Board (IRB): Approved by the IRB of IRCCS Neuromed, Italy (Protocols: N°9/2015, N°19/2020, N°4/2023). **Pag. 26 lines 10-17**

-ClinicalTrials.gov Registration: The study is registered under identifiers NCT02403765, NCT04620980, and NCT05721911. **Pag. 26 lines 12-13**

-Declaration of Helsinki: All clinical investigations were conducted according to the principles of the Declaration of Helsinki. **Pag. 26 lines 14-17**

-Informed Consent: Written informed consent was obtained from all study participants. **Pag. 26 line 16**

**3. Data Stratification & Quality Control**
- Genetic Stratification: Patients were stratified into two distinct groups:
iPD: Idiopathic (no variants in PD genes).
gPD: Genetic (pathogenic mutations in LRRK2, GBA1, TMEM175, PARK2, PINK1, PARK7). **Pag. 5 lines 3-5**

- Sex-Matching: Healthy controls (HC) were sex-matched to the PD cohort to minimize confounding bias in metabolomic analysis. **Pag. 5 line 4 and pag. 27 Lines 3-5**

- Confounder Adjustment: Statistical analyses were adjusted for age, disease duration, and L-DOPA Equivalent Daily Dose (LEDD). **Pag. 29 lines 16-18**

**4. Technical Validation**
- NMR Quantitation: Metabolite identification and quantification were performed using Chenomx NMR Suite and confirmed with Bayesil software. **Pag. 29 lines 3-9**

**5. Data Availability & Reproducibility**
- Software used: MetaboAnalyst 6.0 (Metabolomics) **Pag.30 lines 7-30;** Chenomx (NMR) **Pag. 29 lines 3-9,** Small Molecules Pathways Database (SMPDB) **Pag. 31 Lines 1-4**; Permutational Multivariate Analysis of Variance (PERMANOVA) test **Pag. 30 Lines 4-5.**

- Public Databases: NA

**1. Study Design & Setting (STROBE Items 4 & 5)**
- Design Type: Defined as a case-control observational study. **Pag.26 line 1**

- Study Periods: Two recruitment windows specified (June 2015–Dec 2017 and June 2021–Dec 2023) **Pag. 25 lines 8-10.**

- Location: Parkinson Centre of the IRCCS INM Neuromed, Italy **Pag 25. Lines 4-5.**

**2. Participant Selection & Eligibility (STROBE Item 6)**
-Case Definition: PD diagnosis based on ≥2 cardinal motor signs (tremor, bradykinesia, rigidity) and positive response to L-DOPA. **Pag. 25 lines 14-17**

-Control Definition: Healthy subjects (HC) negative for PD gene mutations, matched for sex with the PD cohort. **Pag.26 lines 22-24 and Pag.27 lines 1-2.**

-Age Threshold: Inclusion limited to individuals aged ≥40 years to maintain cohort relevance. **Pag.26 line 24**

- Exclusion Criteria: Explicitly listed (pre-existing psychiatric conditions, other neurodegenerative diseases like MS or ALS, dementia, depression, and use of specific psychotropic medications). **Pag. 25 lines 17-21**

**3. Data Sources & Clinical Assessment (STROBE Item 8)**
-Clinical Scale: MDS-UPDRS Part III used for motor symptom severity (assessed during the "ON" period). **Pag. 25 lines 22-25.**

-Ancestry: Confirmed European ancestry for all participants. **Pag. 25 lines 8-9**

-Biobank Origin: Subjects selected from the IRCCS Neuromed/IGB-CNR biobank. **Pag.25 lines 6-10**

**4. Genetic Stratification Framework**
- Sequencing Method: Whole Exome Sequencing (WES) data analyzed for the presence of mutations/variants in PD genes. **Pag.27 lines 8-20.**

- Group Classification:
iPD (Idiopathic): No mutations in PD genes. **Pag. 27 lines 17-18.**
gPD (Genetic): Carrying known pathogenic mutations (e.g., LRRK2 G2019S, GBA1). **Pag. 27 lines 11-16.**

**5. Laboratory & Analytical Protocols**
- Serum Handling: Standardized 6-hour fasting collection, 30-min clotting, and -80°C storage. **Pag. 28 lines 1-6.**

- NMR Parameters: 600 MHz Bruker spectrometer **Pag.28 lines 16-19**, CPMG pulse sequence **Pag.28 lines 22-24**,

- Standardization: Use of anonymized codes and internal reference signals (TSP) **Pag. 28 lines 14-16**.

**6. Statistical & Confounding Control (STROBE Items 10 & 12)**
- Sample Size: Acknowledged as determined by biobank availability (no formal a priori calculation). **Pag. 26 lines 20-21**

- Normality Testing: Metabolite concentrations were log-transformed prior to analysis to improve normality assumptions; robust (HC3) standard errors were also applied to account for deviations from normality and heteroscedasticity. **Pag. 31 Lines 17-18**

- Confounder Adjustment: Models were adjusted for relevant covariates, including age, sex, disease duration, and LEDD, with sex also included as an interaction term where appropriate. **Pag. 29 lines 19-20**

- Multivariate Analysis: Multivariate analyses included PCA (validated by PERMANOVA), PLS-DA with cross-validation (R2, Q2), and Random Forest classification with OOB error estimation. **Pag. 30 lines 4-9.**

- Multiple Testing: Multiple comparisons were controlled using the Benjamini–Hochberg FDR correction across metabolite-level and enrichment analyses, with FDR < 0.05 considered significant. **Pag. 31 lines 9-12 and 20-21.**
